# Global seasonal activities of respiratory syncytial virus before the COVID-19 pandemic: a systematic review

**DOI:** 10.1101/2023.06.12.23291266

**Authors:** Songwei Shan, Weixin Zhang, Huizhi Gao, Pei-Yu Huang, Zhanwei Du, Yuan Bai, Yiu-Chung Lau, Dongxuan Chen, Eric HY Lau, Joshua Nealon, Peng Wu

**Affiliations:** World Health Organization Collaborating Centre for Infectious Disease Epidemiology and Control, School of Public Health, Li Ka Shing Faculty of Medicine, The University of Hong Kong, Hong Kong SAR, China; Laboratory of Data Discovery for Health, Hong Kong Science and Technology Park, Hong Kong SAR, China

**Author notes:** Corresponding author: Peng Wu, School of Public Health, The University of Hong Kong.

**Keywords:** activity, respiratory syncytial virus, seasonality, global

## Abstract

**Background:** Varied seasonal patterns of respiratory syncytial virus (RSV) have been reported worldwide. We aimed to review the patterns of RSV activity globally before the COVID-19 pandemic and to explore factors potentially associated with RSV seasonality.

**Methods:** We conducted a systematic review on articles identified in PubMed reporting RSV seasonality based on data collected before 1 January 2020. Information on the timing of the start, peak, and end of an RSV season, study location, study period, and details in study methods were extracted. RSV seasonal patterns were examined by geographic location, calendar month, analytic method and meteorological factors including temperature and absolute humidity. Correlation and regression analyses were conducted to explore the relationship between RSV seasonality and study methods and characteristics of study locations.

**Results:** RSV seasons were reported in 209 articles published in 1973-2023 for 317 locations in 77 countries. Variations were identified in types of data, data collection and analytical methods across the studies. Regular RSV seasons were similarly reported in countries in temperate regions, with highly variable seasons identified in subtropical and tropical countries. Durations of RSV seasons were relatively longer in subtropical and tropical regions than from temperate regions. Longer durations of RSV seasons were associated with a higher daily average mean temperature and daily average mean absolute humidity.

**Conclusions:** The global seasonal patterns of RSV provided important information for optimizing interventions against RSV infection. Heterogeneity in study methods highlighted the importance of developing and applying standardized approaches in RSV surveillance and data reporting.

## INTRODUCTION

Respiratory syncytial virus (RSV) is a leading cause of hospitalization for acute lower respiratory tract infection (LRTI) in young children less than 5 years old ^1–3^, as well as an increasingly recognized important pathogen in older adults (aged ≥ 65) ^4, 5^. Globally, RSV was estimated to cause nearly 25 million LRTI episodes and more than 70,000 deaths every year ^6^. In addition to Palivizumab, a monoclonal antibody recommended for high-risk infants ^7, 8^ providing a short-term protection against RSV-associated severe infections ^9, 10^, a new monoclonal antibody nirsevimab with a prolonged protection ^11, 12^ has been approved by the EU and the UK ^13^. Recently the first RSV vaccine (Arexvy, RSVPreF3 from GSK) approved for the use in older adults and several other vaccine candidates in their final stage of testing demonstrated promising efficacy in preventing RSV-associated LRTI and severe infections^14, 15^.

Activities of RSV varied temporally and geographically. In temperate regions, the highest viral activities have been observed during winter months ^16, 17^ with similar intensities across different years. Two-year cycles of RSV activity have been reported for some countries in Northern Europe, with a major epidemic wave in winter followed by a minor one in spring next year ^18^. In subtropical and tropical regions, seasonal patterns of RSV were more diverse and less clear than those reported for temperate countries ^19^. Knowledge about the seasonality of RSV, can guide the formulation of prevention strategies and the planning of public health resources. In particular, accurate estimates of the RSV season start and its duration can help to determine the optimal timing of initial doses and optimal dosing interval for prophylactic monoclonal antibodies and vaccination schedules.

We conducted a systematic review on RSV seasonality to characterize the global patterns of RSV activity by summarizing the reported start, peak and end of RSV seasons worldwide and describe approaches applied in analyzing the seasonality. We aimed to provide a better understanding of the seasonal patterns of RSV circulation to inform policy and decisions in prevention, diagnosis, and treatment of RSV infections.

## METHODS

### Literature search strategy and selection criteria

This systematic review was conducted following the Preferred Reporting Items for Systematic Reviews and Meta-Analyses (PRISMA) guidelines ^20^. We focused on studies in which seasonal patterns of RSV were characterized by providing the time of the start, peak and/or end of a season based on laboratory-confirmed RSV infection data. We searched published articles from PubMed on January 17, 2023 using the combination of following search items (*1 and 2*) in “Title/Abstract” (Figure 1):

1. “RSV” or “Respiratory Syncytial Virus”
2. “seasonality” or “season*”

**Figure 1.**
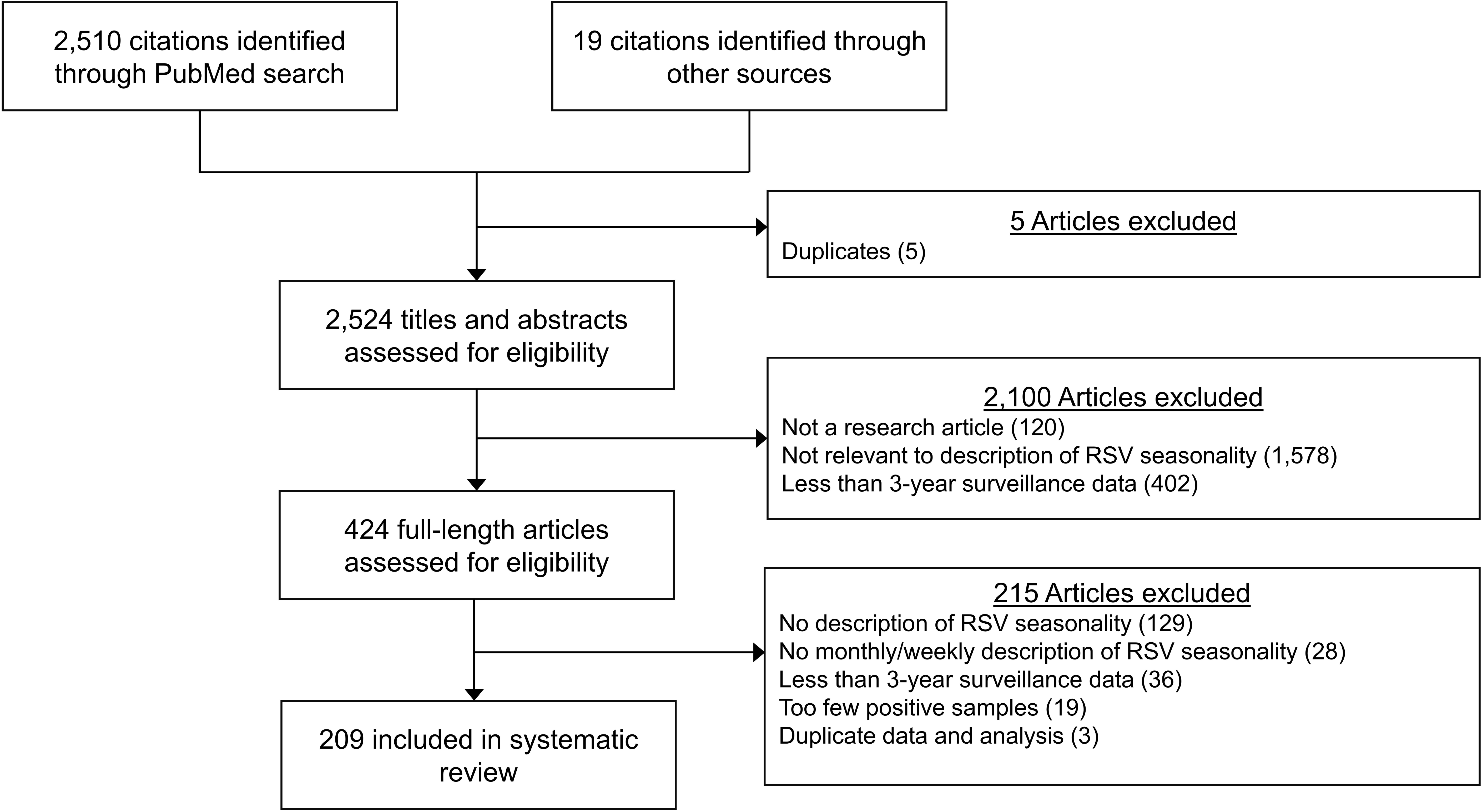
Flowchart of study identification and selection for the systematic review.

Two authors (S.S. and W.Z.) performed the literature search (#1 and #2) and screened retrieved articles independently. Full-text screening was conducted after title/abstract screening. Two authors (S.S. and W.Z.) independently identified articles for inclusion. Any disagreement in article selection and data retrieval was resolved by a third author (P.W.).

Eligible articles were studies reporting the seasonality of RSV based on monthly or weekly laboratory-confirmed RSV infections collected for a specified place, i.e., a city, a province or a country, for three years or longer. We excluded articles if: 1) the full text was not available or not in English, 2) the analysis was based on less than three years’ data, 3) the average number of reported RSV infections per year in a study was less than 50, 4) the time of the start, peak and/or end of RSV seasons was not specified in a calendar month or week. If an investigation on RSV seasonality covered the time period beyond 2019, the study period before the COVID-19 pandemic ≥ 3 years would be included in our analysis, and the RSV data collected during the pandemic would be excluded. Articles without original data analysis on RSV activities, such as reviews, commentaries, letters, were excluded, but we reviewed the references of these articles for additional studies to include for screening.

### Data extraction and analysis

Two authors (S.S. and W.Z.) independently extracted data into a standardized data extraction form. Data fields included the timing (start, peak, and end) of an RSV season, study location (city, region or country), study period, information on study characteristics (such as the source of cases, case definitions, laboratory testing methods and analytic methods applied to determine RSV seasons). We classified the sources of study cases into outpatient, inpatient, outpatient and inpatient, case definitions into influenza-like-illness (ILI) or acute respiratory infection (ARI), severe acute respiratory infection (SARI) and acute LRTI and grouped laboratory testing methods for RSV confirmation into virus detection, antibody detection, antigen detection, and nucleic acid detection.

One publication might contain several different investigations on seasonal patterns of RSV, either based on different data sources, or based on the same dataset but with different analytic methods. If more than one estimate on timing of RSV season was reported in a single publication, using different data sources or methods, all were extracted and included in our analyses. It was common that one investigation on RSV seasonality was conducted based on more than one type of case source, case definition and/or testing method, and all the methodological details were extracted and documented for analyses. Analytic methods used by different studies to determine the RSV seasons were classified as qualitative methods and quantitative methods based on whether the analysis on RSV seasonality was carried out using a statistical or mathematical approach, and the quantitative methods were further divided into three groups, threshold-based, coverage-based and model-based (***Supplementary Materials***).

In addition to the study-specific information, we also documented the latitude, daily average mean temperature and daily average mean relative humidity for all study locations reported in each investigation using Google Geocoding API ^21^ and R package GSOR ^22^(Supplementary materials). The extracted daily average mean temperature and relative humidity data were used to derive the daily average mean absolute humidity for each location. We also extracted the geoclimatic information of each study site, and classified the investigations of RSV seasonality into three groups based on the latitude of the study sites, temperate zone (latitude > 35° or latitude < -35°), subtropical zone (23·5° < latitude ≤ 35° or -35° ≤ latitude < -23·5°) and tropical zone (−23·5° ≤ latitude ≤ 23·5°).

We described the reported RSV seasons as a function of geographical location, meteorological factors and characteristics of study design, and explored potential associations using Pearson correlation analyses and Kendall’s rank correlation between the duration of the season and the latitude, daily average mean temperature and daily average mean absolute humidity of study sites as well as the analytic method and climate zone. Finally, linear regression models were applied to explore the independent associations between the duration of RSV seasons and the abovementioned factors as covariates. The statistical analyses were conducted using R software Version 4.0.3 (R Foundation for Statistical Computing, Vienna, Austria).

## RESULTS

We identified 2,510 citations from the PubMed through the keyword search and additional 19 citations from other sources. In total, 209 publications were included after screening based on the inclusion and exclusion criteria (Figure 1) and 552 investigations of RSV seasonality were identified and included in analyses.

The identified studies were published between 1973 and 2023, with the majority (147/209, 70%) from 2010 onwards (Supplementary Figure 1). RSV seasonality was reported for 317 distinct study sites in 77 countries from Asia, Europe, North America, South America, Oceania and Africa (Figure 2). Most investigations (342/552, 62%) were conducted with a study period of 3-6 years (Supplementary Figure 1), with the longest one from Malaysia using surveillance data over 27 years ^23^. Among the identified publications, more than half (124/209, 59%) were from temperate regions (latitude > 35° or latitude < -35°), followed by 31% (65/209) and 22% (47/209) in subtropical (23·5° < latitude ≤ 35° or -35° ≤ latitude < - 23·5°) and tropical (−23·5° ≤ latitude ≤ 23·5°) regions, respectively (Table 1, Supplementary Table 1).

**Figure 2.**
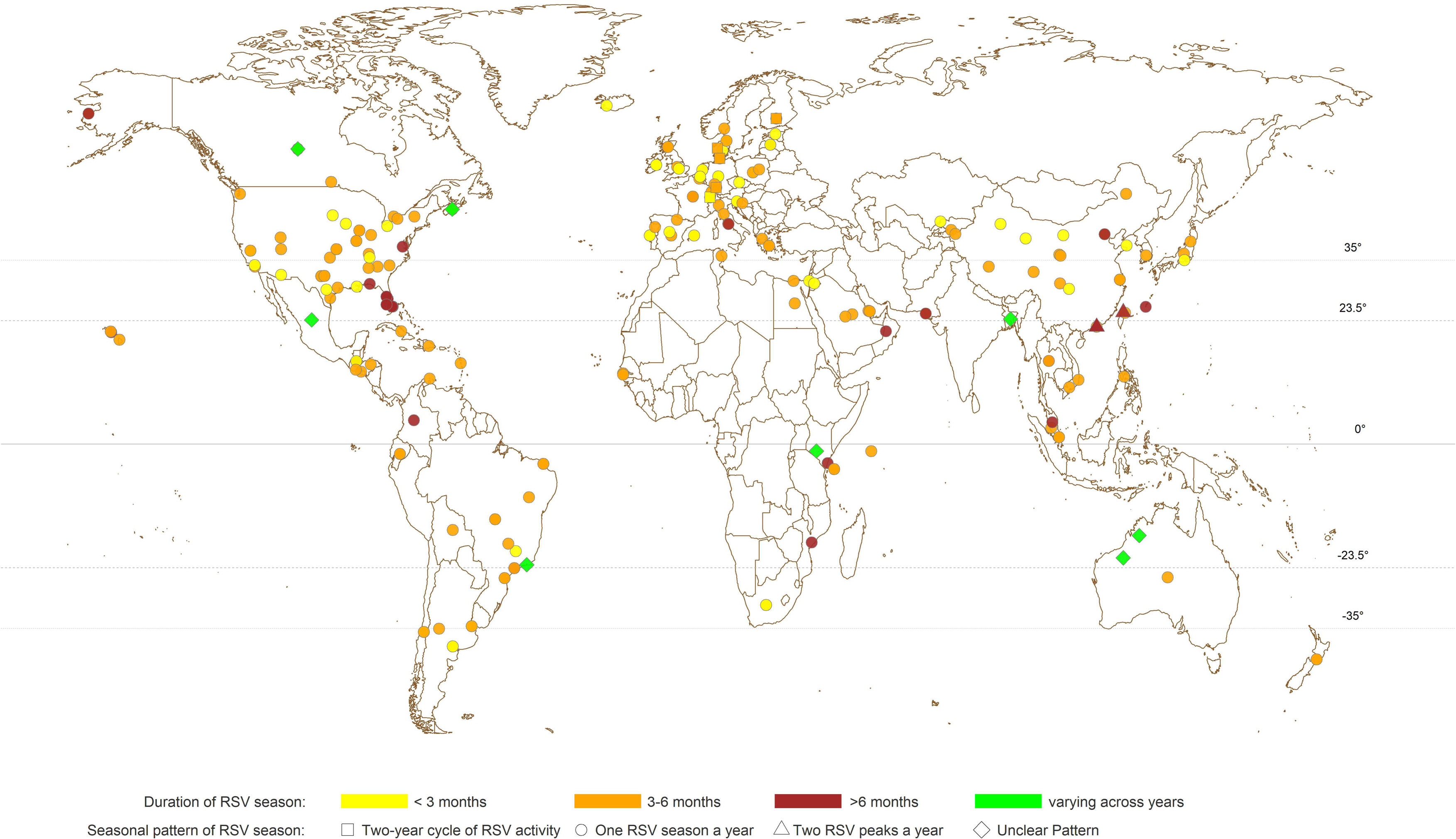
Geographic distribution of study locations and the reported RSV seasonal patterns from all included investigations using either quantitative or qualitative approaches.

**Table 1.** Characteristics of publications and investigations included in the systematic review.

| Characteristics | No. of Publications<br>(N=209)<sup>a</sup><br>n (%) | No. of Investigations<br>(N = 552)<br>n (%) |
| --- | --- | --- |
| **Climatic zone** |  |  |
| Temperate (>35°) | 123 (58.9) | 335 (60.7) |
| Subtropical (23.5° - 35°) | 65 (31.1) | 134 (24.3) |
| Tropical (equator - 23.5°) | 49 (23.4) | 83 (15.0) |
| **Continent** |  |  |
| Asia | 64 (30.6) | 140 (25.4) |
| Europe | 68 (32.5) | 155 (28.1) |
| North America | 38 (18.2) | 164 (29.7) |
| South America | 14 (6.7) | 49 (8.9) |
| Africa | 12 (5.7) | 24 (4.3) |
| Oceania | 5 (2.4) | 19 (3.4) |
| Others<sup>b</sup> | 8 (3.8) | 1 (0.2) |
| **Study period<sup>c</sup>** |  |  |
| Before Palivizumab approved | 88 (42.1) | 147 (26.6) |
| After Palivizumab approved | 75 (35.9) | 320 (58.0) |
| Others<sup>d</sup> | 46 (22.0) | 85 (15.4) |
| **Length of study period** |  |  |
| 3-6 years | 127 (60.8) | 342 (62.0) |
| 7-10 years | 53 (25.4) | 123 (22.3) |
| > 10 years | 37 (17.7) | 87 (15.8) |
| **Type of RSV data** |  |  |
| Weekly data | 57 (27.3) | 294 (53.3) |
| Monthly data | 155 (74.2) | 258 (46.7) |
| **Analytic method** |  |  |
| Qualitative | 146 (69.9) | 179 (32.4) |
| Quantitative | 66 (31.6) | 373 (67.6) |
| Threshold-based | 54 (25·9) | 205 (37·1) |
| Coverage-based | 8 (3·8) | 142 (25·7) |
| Model-based | 6 (2·9) | 26 (4·7) |
| **RSV case source** |  |  |
| Inpatient | 94 (45·0) | 152 (27·5) |
| Outpatient | 14 (6·7) | 25 (4·5) |
| Both | 53 (25·4) | 115 (20·8) |
| Unknown | 48 (23·0) | 260 (47·1) |
| **RSV case definition <sup>e</sup>** |  |  |
| ARI or ILI | 82 (39·2) | 150 (27·2) |
| SARI | 23 (11·0) | 45 (8·2) |
| ALRI | 48 (23·0) | 59 (10·7) |
| Clinical judgement | 9 (4·3) | 30 (5·4) |
| Unknown | 70 (33·5) | 310 (56·2) |
| **Testing method <sup>f</sup>** |  |  |
| Virus detection | 45 (21·5) | 168 (30·4) |
| Antigen detection | 107 (51·2) | 303 (54·9) |
| Antibody detection | 8 (3·8) | 42 (7·6) |
| Nucleic acid detection | 110 (52·6) | 323 (58·5) |
| Unknown | 33 (15·8) | 127 (23·0) |

Note: ARI, acute respiratory infection; ILI, influenza-like illness; SARI, severe acute respiratory infection; ALRI, acute lower respiratory infection
<sup>a</sup>An investigation was identified as an analysis with a specified method on the reported RSV data for the seasonality of RSV activities. Analyses on the same data with different analytic methods were recognized as different investigations. A publication might include several investigations describing the seasonality of RSV activities by analysing different sets of data, or applying different analytic methods, etc. Therefore, the sum of investigations might be greater than the total number of publications.
<sup>b</sup>Refers to the publications/investigations with study sites involving more than one continent.
<sup>c</sup>For different locations, the time for palivizumab to be licensed for prophylactic use are different. For the countries where palivizumab has not been licensed, such as China, all the publications/investigations of this country would be recognized as “before palivizumab approved”.
<sup>d</sup>Refers to the publications/investigations in which the study period contains the periods both before and after the year when Palivizumab was approved in the study locations.
<sup>e</sup>A publication/investigation on RSV seasonality might be based on multiple sources of cases, e.g., ARI, ILI and SARI perhaps were used together or in different combinations to identify patients for RSV testing. Some investigations/studies might be counted multiple times, and the sum of the proportions of publications/investigations from different case definition categories may exceed 100%.
<sup>f</sup>A publication/investigation on RSV seasonality might adopt multiple testing methods for RSV detection. Some investigations/studies might be counted multiple times, and the sum of the proportions of publications/investigations from different testing method categories may exceed 100%.

The information on the case source, case definition, and laboratory testing methods was missing in some investigations (case source: 260/552, 47%; case definition: 310/552, 56%; testing method: 127/552, 23%). Among those with information of case source and case definition provided, the majority of investigations (152/292, 52%) identified RSV cases from inpatients, and ARI or ILI (150/242, 62%) was the most frequently used syndromic definitions to screen patients for RSV testing, and antigen (325/552, 59%) and nucleic acid detections (323/552, 59%) were the most commonly used laboratory testing methods in more recent publications (Table 1, Supplementary Table 1, Supplementary Figures 2-3). The majority of investigations (373/552, 68%) applied quantitative methods to determine the time of the start, peak and/or end of seasons, and among these studies, over half of the investigations used the threshold-based method (205/373, 55%) (Supplementary Tables 2-3).

**Figure 3.**
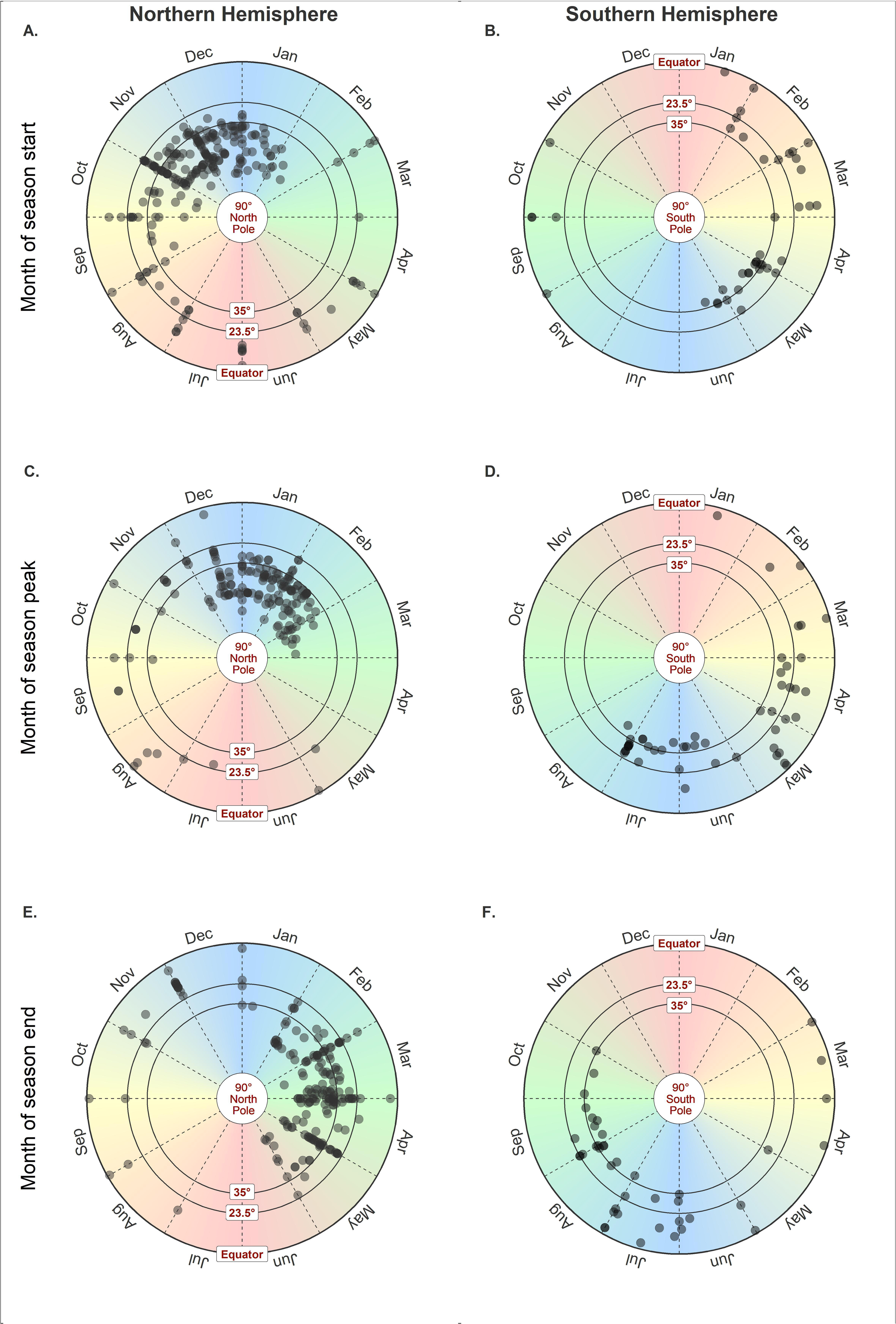
The timing of one-peak RSV seasons reported in investigations using either quantitative or qualitative approaches against the latitude of the study site located in the Northern Hemisphere (NH, centred at the north pole) in panels A, C and E, and Southern Hemisphere (SH, centred at the south pole) in panels B, D and F. The start (A), peak (C) and end (E) of RSV seasons in NH. The start (B), peak (D) and end (F) of RSV seasons in SH. The centre of the radius circles represents the north pole or the south pole (latitude = 90°) and the external border represents the equator (latitude = 0°). The circles were divided by the dashed radiuses into 12 sections to represent 12 months of a year. Two radius circles between the north/south pole and the external border indicating the latitudes at 23·5° and 35° are used to define the temperate region (35°-90°), subtropical region (23·5°-35°) and tropical region (equator-23·5°).

The majority of the investigations were conducted using data collected from locations in the Northern Hemisphere (NH) and indicated one RSV season every year (Figure 2). A small number of investigations (11/552, 2%) reported two RSV peaks identified in one year (Supplementary Table 4) while 18 (3%) investigations indicated unique RSV seasonal patterns repeated every 2 years (Supplementary Table 5). Among study sites reporting one RSV season annually, locations in temperate countries were fairly consistent in the time of the start, peak and end of RSV seasons in either the NH or Southern Hemisphere (SH) (Figure 3). In temperate countries, RSV activities generally started from late autumn or early winter, i.e., October-December in NH, May-June in SH, peaked in winter (December-February in NH, and June to July in SH), and ended in late winter or spring (February-May in NH, and July-September in SH). (Figure 3, Supplementary Figure 4). Larger variations were observed in the start, peak and end month of RSV seasons in subtropical and tropical countries at a lower latitude, with high RSV activities observed in July/August-November in tropical locations (Supplementary Figures 5-6). The durations of RSV seasons were relatively shorter in study sites in the temperate region (median: 3·97 months, interquartile range (IQR): 3·00-5·00) compared with the estimates from subtropical (4·00, 3·52-6·14) and tropical countries (5·00, 4·00-6·00) (Figure 2, Supplementary Figure 6).

The Pearson correlation analysis indicated an overall negative association between the duration of the RSV season (in months) and the latitude both in the NH (p<0·001) and the SH (p=0·002) (Supplementary Table 7), and the statistical significance in the association was only indicated with the estimates derived from quantitative methods (p<0·001). Positive associations were indicated in the correlation analyses between the daily average mean temperature (correlation coefficient: 0·33; 95% CI: 0·22, 0·44) and absolute humidity (0·34; 0·22, 0·45) of the study locations and the durations of RSV season estimated by the quantitative methods (p<0·001) but not for those estimated by the qualitative methods (Supplementary Table 7). Durations of RSV seasons appeared to be longer in tropical/subtropical regions than the estimates from temperate regions, and the estimated RSV seasons were longer from the threshold-based methods, but shorter from the coverage-based method (Table 2, Supplementary Table 8).

**Table 2.** Associations identified in the linear regression analysis between durations of RSV seasons estimated from quantitative approaches and characteristics of the studies.

| Variable | Change in duration of RSV season (months) | 95% confidence interval | p-value |
| --- | --- | --- | --- |
| **Climatic zone** |  |  |  |
| Temperate region (<-35° or >35°) | 0 | Referent |  |
| Subtropical region (-35° - -23.5° or 23.5° - 35°) | 1.06 | 0.45, 1.66 | <0.001 |
| Tropical region (-23.5° – 23.5°) | 1.65 | 0.77, 2.54 | <0.001 |
| **Analysis method** |  |  |  |
| Coverage-based method | 0 | Referent |  |
| Threshold-based method | 1.43 | 0.96, 1.90 | <0.001 |
| Model-based method | 1.98 | -0.99, 4.95 | 0.19 |
| **Meteorological factor** |  |  |  |
| Daily average mean absolute humidity | 0.013 | -0.0049, 0.030 | 0.15 |

In the main analysis with the linear regression model, we identified that climatic zone appeared to be independently associated with the estimated duration of RSV seasons, and analytic method used in estimating RSV seasons, annual daily average mean absolute humidity at the study location could also explain some variations (Adjusted R^2^=0·33 for estimates from quantitative approaches, 0·25 for all estimates) in the estimated durations of RSV seasons (Table 2, Supplementary Table 8). Considering the close intercorrelations between climatic zone, latitude and temperature, we included the climatic zone (Table 2, Supplementary Table 8), latitude (Supplementary Tables 9-10) and daily average mean temperature (Supplementary Tables 11-12) of study locations into different linear regression models separately. The duration of RSV season would decrease by 0·041 (95% CI: 0·017, 0·064) months with every increase of 1 degree in latitude, and increase by 0.058 (95%CI: 0.0077, 0.11) months with every increase of 1 degree in daily average mean temperature.

Durations of RSV seasons estimated for the subtropical and tropical locations were on average 1·06 (95% CI: 0·45, 1·66) and 1·65 (95% CI: 0·77, 2·54) months longer than for the study sites in the temperate region. Compared to the coverage-based methods, the threshold-based methods tended to produce longer estimates of the duration of RSV season by 1·43 (95% CI: 0·96, 1·90) months. Overall, the duration of RSV season prolonged by 0·013 (95% CI: - 0·0048, 0·030) months with every 1 g/m^3^ increase in absolute humidity at the study location (Table 2).

## DISCUSSION

RSV seasonal patterns have been studied widely in particular since the last decade, with most of the data reported from temperate countries in the Northern Hemisphere. Our review indicated that largely similar seasonal patterns of RSV were observed in locations from the same climatic region either in the NH or the SH. In temperate regions, RSV activities primarily showed regular annual patterns starting in winter seasons, and the estimated durations of RSV season were relatively shorter than the reported from tropical and subtropical regions where prolonged RSV circulations were often more than half a year with greatly varied times of season start and/or peak.

Human infections of RSV are likely driven by two major antigenic groups, A and B ^24^ between which cross-immunity has been suggested ^25, 26^. However, RSV typing is often not included in most of the established surveillance systems ^27^. The increasing susceptibility in infants due to the loss of protection from waning maternal antibodies leads to a higher risk of infection particularly in young children with high-risk conditions ^1, 28, 29^, potentially driving the transmission of RSV in the population ^30^ although the virus can infect people at any age including older adults ^31, 32^. RSV shows comparable environmental survivability and transmissibility to influenza viruses with a similar estimate of the basic reproduction number (R_0_) and household second attack rate ^33^. Increasing RSV activities were commonly reported in cold seasons starting from late autumn through to early spring in temperate regions, and in rainy seasons in tropical/subtropical areas, and the associations between RSV activities and environmental factors including temperature, relative/absolute humidity, rainfall were inconsistently reported from studies conducted in tropical areas ^34–36^.

RSV seasonality characterised in different studies appeared to be associated with the approaches used in data collection and analysis ^19^. Previous research demonstrated that the timing of RSV seasons determined with laboratory surveillance data might be different from the results based on hospitalized patient data although the two estimates were highly correlated ^37^. The improvement of laboratory testing methods could also affect the estimates of RSV seasons ^38, 39^. Our review showed that estimates of the timing of RSV seasons were correlated with the analytic approaches used in the studies, with shorter durations of RSV seasons estimated from “coverage-based method” (defined as the shortest time period when the majority of confirmed (severe) RSV infections occurred) than the estimates from the threshold-based methods which although were highly dependent on the often arbitrarily selected threshold of varied measures for RSV activities, such as proportion of virus detections or patients presenting ILI/ARI symptoms ^40^. Therefore, prudent comparison and interpretation of RSV seasons estimated from different studies or with different approaches is necessary.

RSV surveillance is often established based on the existing routine surveillance on influenza, more commonly seen in developed economies including the US ^41^, Canada ^42^ and some European countries ^43^. The WHO initiated a pilot RSV surveillance scheme by leveraging the current Global Influenza Surveillance and Response System covering over 100 countries to explore possible surveillance strategies including case definitions, laboratory methods and target population, etc. ^44^. Current RSV surveillance largely applied similar syndromic case definitions as those used for influenza surveillance, i.e., sampling patients showing influenza-like-illness and testing for the virus, while the validity of using such case definitions for RSV surveillance has been questioned ^45^. In some countries/areas RSV surveillance was not conducted throughout a year but only during restricted period(s) following the influenza surveillance protocol given the regular annual influenza seasons in those locations ^17, 46^. However, this might lead to an underreporting of RSV infections during the non-surveillance periods ^47^.

During the COVID-19 pandemic, circulation of respiratory viruses including RSV changed considerably worldwide largely due to implementation of non-pharmaceutical interventions and changes in human behaviours in response to the pandemic ^48, 49^. RSV remained at a low level in many countries in 2020 when public health and social measure were widely adopted globally to contain the spread of SARS-CoV-2, but caused surges of infections with a large number of severe cases admitted into hospital in some countries when some interventions were relaxed from 2021 onwards ^50, 51^, including the US during 2021-2023 ^52^, some geographically separated locations in Australia in 2020-21 ^53^ and the UK in 2021-22 ^54^. The inter-seasonal viral activities were perhaps related to the increased susceptibility in the population due to the lack of exposure to RSV during the previous year in response to COVID-19 ^55^.

Monoclonal antibodies approved for prophylactic therapy provide immediate protection of high-risk individuals particularly young children against RSV-associated severe infections ^7, 13^. Five monthly injections of palivizumab and a single dose of nirsevimab can substantially reduce the risk of hospitalizations due to RSV in individuals receiving the therapy throughout RSV seasons especially in temperate regions where RSV seasons would normally not be longer than 5 months. The duration of protection provided by RSV vaccines has not been fully determined although promising efficacy for short-term prevention from infections was demonstrated in clinical trials ^56–58^. The effectiveness of the abovementioned pharmaceutical interventions for high-risk population is determined by the duration of protection of the monoclonal antibodies and the vaccines, the timing of administration and RSV activities in specific locations. Ideally these preventive measures should be administered before an RSV season starts, and the protection can maintain throughout the season, likely during November - April, and May - October in temperate regions of the NH and the SH, respectively. However great challenges in implementing these efficacious interventions would be seen in locations where there were irregular seasons particularly with prolonged viral circulations ^59–61^. Based on the reported seasonal patterns in locations from northern temperate regions in this review, the Studies incorporating epidemiologic analysis on RSV seasonality into the effectiveness and cost-effectiveness of these RSV preventive strategies are urgently needed ^62^.

There are several limitations in this study. First, data of RSV seasonality that were published and included in the review were skewed to high-income countries with well-established RSV surveillance systems. These countries are largely in temperate regions where RSV circulation might be different from the underrepresented middle- and low-income countries due to varied population structure, contact patterns and geoclimatic features ^63^. Second, RSV antigenic information could not be incorporated into the current analysis of seasonality due to the lack of data. Expanding RSV surveillance by collecting antigenic and genetic data is urgently needed to improve our understanding of the transmission dynamics of RSV locally and globally ^64^. Third, except for the analytic method, we could not examine potential associations between RSV seasonality and other study characteristics such as case sources, case definitions, and laboratory methods due to the lack of information and non-standardized laboratory protocols used for RSV surveillance, or explore the association between RSV seasonality and population structure particularly birth rate given the lack of data although a high disease burden of RSV identified in young children ^65–67^ implied a possible driving force from the young population on RSV activities. Lastly, the reported data of seasonality in this review did not allow us to investigate the RSV-associated disease burden occurred during the characterised RSV seasons which is a critical metric for evaluation of the health impact related to RSV circulation, particularly when there were large uncertainties in the start and the end of RSV seasons in a specific location and the infection risk is less likely to be homogeneous in a population over the season.

## CONCLUSIONS

This study provided a global overview of RSV seasonality before the COVID-19 pandemic. The seasonal patterns illustrated in this review may inform optimal use of preventive and therapeutic interventions against RSV infection particularly in temperate regions of the NH and SH. Considerable variations in methods used by different studies highlighted the importance of developing and applying standardized approaches in RSV surveillance and data reporting. Further research is needed to understand the underlying mechanisms driving RSV seasonal patterns locally and globally.

## Data Availability

All data produced in the present study are available upon reasonable request to the authors

## Acknowledgements

ACKNOWLEDGEMENTS

The authors thank Prof. Julian W Tang, and Dr. Eeva Broberg for kindly sharing the data of RSV seasonality from their publications. The authors also thank Stella SW Lai for technical assistance.

## AUTHOR CONTRIBUTIONS

All authors meet the International Committee of Medical Journal Editors criteria for authorship. SS, PW, BY, DZ conceived and designed the study; SS, WZ, HG, PH collected and extracted data; SS, PW conducted the data analysis and wrote the first draft of the manuscript. All authors contributed to the interpretation of the results and revision of the manuscript, and have approved the final version of the manuscript.

## DECLATATION OF INTERESTS

JN was previously employed by and owns shares in Sanofi. The authors report no other potential conflicts of interest.

