## Supporting Information for "Global seasonal activities of respiratory syncytial virus before the COVID-19 pandemic: a systematic review"

**Literature search and selection**

We performed a literature search on PubMed on 17 January 2023 with search terms pertaining to respiratory syncytial virus (RSV) (#1 “RSV OR respiratory syncytial virus”), and seasonlity (#2 “seasonality OR season*”) in the “Title/Abstract” field, without restrictions on the publication time and language. The search results were shown below.

|  | **Search terms** | **Number of publications identified** |
| --- | --- | --- |
| RSV disease | #1: (RSV OR respiratory syncytial virus) | 23,005 |
| Seasonality | #2: (Seasonality OR season*) | 201,170 |
| RSV seasonality | #3: #1 AND #2. | 2,510 |

After removing the duplicates and articles not meeting the inclusion criteria (See Methods), our review focused on publications estimating RSV seasons with a study period of three years or longer. If a publication included data from a time period beyond the year 2020, only the data before January 1, 2020 would be included into the analysis. Besides, the remaining study period after excluding the data from 2020 onwards should be at least three years. For example, if a publication applied the data collected from 2016 through to 2021 to estimate RSV seasonality, we would only include RSV seasonal patterns indicated by the data collected during 2016-2019 into our analysis.

**Latitude coordinates of the studied sites**

We extracted the latitude and longitude coordinates for each study site with RSV seasonal patterns reported using Google Geocoding API [[1]](https://paperpile.com/c/zPIKrB/pdyI). For multicenter studies we used the coordinates of the centroid of multiple centers. For the 10 Health and Human Services (HHS) [213] regions of the United States, we used coordinates of the corresponding reference cities following the study by Staadegaard et al. [[2]](https://paperpile.com/c/zPIKrB/aIPfL).

**Temperature and absolute humidity of studied sites**

We used the latitudes inferred by the above-mentioned method to represent the study sites and collected the daily average temperature, average relative humidity from the nearest weather station providing the data within the study period using R package GSODR. The daily average absolute humidity was calculated from the temperature and relative humidity. Each season estimate was associated with a value of dialy average mean temperature and daily average mean absolute humidity by calculating the mean of the daily average temperature and daily average absolute humidity within the study period for further analysis.

**Categorization of RSV seasonal patterns**

We classified the RSV seasonal patterns into four different types based on the reported season timing (the start, peak and end) during the study period.

1. One RSV season a year. RSV activity presented one season every year, and the time of RSV season start, peak and end were similar during the study period (differences of season starts, peaks, and ends across different years were within 3 months). There was only one peak of RSV activity reported in a year during the study period.
2. Two-year cycle. The seasonality of RSV presented a two-year cycle in terms of the timing of season start or peak, or the magnitude of peak in the study period, i.e. RSV activity patterns repeatedly occur every 2 years. Under this pattern, RSV activity may exhibit an alternating pattern with an early season followed by a late season, and/or a mild outbreak followed by a markedly intense one.
3. Two RSV peaks a year. Two peaks of RSV activities occurred in one year during the study period.
4. Unclear pattern. If the original study reported that RSV seasons were not able to be clearly identified, or the timing of RSV seasons (the start, peak, and end) changed substantially across years (differences were larger than 3 months), we put it into the category of “unclear pattern”.

### **Categorization of analysis methods**

We coarsely categorized the methods to determine the RSV seasons as qualitative methods and quantitative methods. If the analysis of RSV seasonality in an investigation was carried out using a statistical or mathematical approach, it would be classified into quantitative methods. Otherwise, if the seasonality of RSV was only reported by descriptive texts and/or graphic illustrations, it would be of qualitative methods.

We further classified the quantitative methods into three types based on the approaches used to determine the RSV seasons: 1) threshold-based methods, which means that if the indicator of RSV activity is beyond a predefined threshold RSV season begins. The indicators used in the methods were different, and the mostly used indicators were positive percentage (what percentage of samples were tested positive), the number of RSV cases, and RSV hospitalization. The thresholds could be defined as fixed values, or dynamically calculated from historical data. Possibly there were additional requirements for the season start other than beyond the threshold values, such as meeting threshold values for a minimum number of consecutive weeks/months, or the minimum requirement of testing/positive samples (Supplementary Table 6). 2) coverage-based methods, which basically defines RSV seasons as a probably shortest consecutive temporal period or intermittent periods covering at least a certain percentage of RSV cases (for example, 75% in [[3]](https://paperpile.com/c/zPIKrB/BQ74y)). 3) model-based methods, which apply certain statistical models (e.g. Poisson regression model [[4]](https://paperpile.com/c/zPIKrB/JH4bW), wavelet model [[5]](https://paperpile.com/c/zPIKrB/o6J01) etc.) to the RSV surveillance data and the seasons were determined based on the fitting results.

**Categorization of case definitions**

Case definition refers to the syndromic definition of the patients selected for RSV testing. We categorized the case definition into four groups, ARI or ILI, SARI, ALRI and clinical judgment. For the studies that the case definition was not clearly described, we categorized them as “Unknown”. If the case definition was described as either ARI (acute respiratory infection) or ILI (influenza-like illness), or as “respiratory disease”, “respiratory symptoms”, the case definition of this study would be classified as “ARI or ILI”. The type “SARI” included the studies in which the criteria were described as “SARI” or hospitalized for respiratory illness. “ALRI” included the inclusion criteria of lower respiratory tract infection including pneumonia and bronchitis. If the inclusion criteria was described as clinical requirement or clinician's discretion, the study would be categorized as “clinical judgment”.

**Categorization of testing methods**

We categorized the testing methods adopted in the included studies into four categories by the substance they detect to confirm RSV infection: virus detection, antibody detection, antigen detection, and nucleic acid detection. Testing methods like cell culture and virus isolation were classified into the group of virus detection, serology and HAI were classified as antibody detection, IF, DFA, EFA, ELISA were generally classified as antigen detection if they test the existence of antigen, and PCR and molecular method were categorized as nucleic acid detection.

**The timing of RSV seasons**

The timing of RSV season (the start, peak, and end) were extracted from the included studies. For the studies using qualitative methods, the estiamtes about the season start, peak and end were extracted from the descriptive text of RSV seasons. For example, from description of “ RSV seasons occur during the winter months from November to March”, season start as November and season end as March were extracted.

All the extracted estimates of the season start, end and peak were transformed to numerical values between 0 to 12, 0-1 representing January and 1-2 representing February, and so on. We assume that there is 52 weeks in a year (no week 53), and weeks 1-52 were evenly transformed to the scope of 0-12.

The timing of RSV seasons (the start, peak, and end) were reported in different formats. The estimates might be a specific month or week, such as “November” or “Week 42”, as a range like “from November to December”, or both with a specific time as the median or mean of multi-year timing and a period representing the range, 95% confidence interval or interquartile range such as “Week 42 (41-45)”.

In Supplementary Figure 4, we tried to provide as much information in the studies as possible, so we plotted the period (range, 95% CI, or IQR) of RSV season start, end and peak when they were available, and plotted the specific week/month otherwise. While in the correlation and regression analysis, the duration of RSV season was calculated using the specific estimate when available, or using the mid-time point of a reported duration when the estimate of a specific time point was not available.

**Linear regression model of durations of RSV seasons with the latitude, climatic zone, absolute humidity and analytic method**

The linear regression model below was used to investigate the relationship between duration of RSV seasons and the latitude, climatic zone, absolute humidity and analytic method. The model was separately applied to quantitative estimates (Table 2) and all estimates (Supplementary Table 8) of RSV seasons for a period not less than 3 years from the included studies.

$$y_{i}=\beta_{0}+\beta_{1}x_{i1}+\beta_{2}x_{i2}+\beta_{3}x_{i3}$$

Where

$i=n$ observed RSV duations

$y_{i}=RSV duration in months$

$$\beta_{0}=intercept (constant term)$$

$$x_{i1}=climatic zone$$

$$x_{i2}=daily average mean absolute humidity$$

$$x_{i3}=analytic method$$

$\beta_{1},\beta_{2}, \beta_{3}$are the corresponding coefficients for $x_{i1},x_{i2},$ and $x_{i3}$

We also applied alternate regression models using the absolute value of latitude and daily average mean temperature to relplace the climatic zone (let $x_{i1}$be the daily average mean temperature and absolute value of latitude in the above regression equation, respectively) to investigate the association between study characteristics and estimated durations of RSV seasons from quantitative methods (Supplementary Table 9 and Supplementary Table 11) and all estimates (Supplementary Table 10 and Supplementary Table 12).

**Possible strategies in prophylactic use of monoclonal antibodies**

Based on the RSV seasonality identified in this review, we intended to examine possible strategies in applying currently available preventive pharmaceutical interventions against RSV epidemics. As indicated in previous efficacy studies and recommendations from the professional organizations, the two approved monoclonal antibodies, palivizumab and nirsevimab, presumably could provide 5-month protection (one dose of nirsevimab or 5 doses of palivizumab) for the individual treated.

Substantial uncertainties of RSV seasonal patterns resulted from the fact that the start or the end the RSV season was reported as a time period covering multiple months, which surely would bring challenges in determining the optimal time for the use of prophylactic therapies in the target population. We investigated the coverage of durations of RSV seasons of 123 distinct estimates extracted from 76 different locations in northern temperate regions where less heterogeneous seasonal patterns were indicated if administering the monoclonal antibodies to provide 5-month protection. If using the month of the mid-point of the reported season start period as the possible time for initiation of the prophylactic therapy for the target population, and the month of the mid-point of the reported season end period as the end of the season, the seasons of the locations in temperate countries were shown to start from autumn to winter months (median: December, range: September-February), and end in winter to spring (median: March, range: December-May), leading to the durations of the RSV seasons being 2 to 8 months (median: 4 months). Assuming that all the 123 locations in northern temperate regions start to administer the monoclonal antibody for the target population in November, 355/563 (60·1%) RSV season months would be covered in these locations, administration in December leading to treated individuals being protected in 480/563 (85·3%) RSV season months, and the initiation in January covering 406/563 (72·1%) season months. If the prophylactic therapy can be initiated exactly on the reported month of season start in each location, the targe population can only be protected in 521/563 (92.5%) RSV season months. However, we were aware that data of seasonality from some locations might be over represented in this analysis.

Alternatively, if using the earliest month of the reported season start period as the possible time for initiation of the prophylactic therapy, and the latest month of the reported season end period as the end of a season, the median months of season start and end were November (range: July-February) and April (range: December – June), respectively, with the median duration of RSV seasons being 5 months (range: 2-9). Assuming that all the 123 locations in northern temperate regions initiated the therapy from November, it would cover 350/563 (62·2%) season months, 516/633 (81·5%) season months if starting from December, 445/633 (70·3%) months if from January. Even if the prophylactic therapy can be initiated exactly on the reported month of season start in each location, the targe population can only be protected in 553/633 (87·4%) RSV season months.

**SUPPLEMENTARY FIGURES**

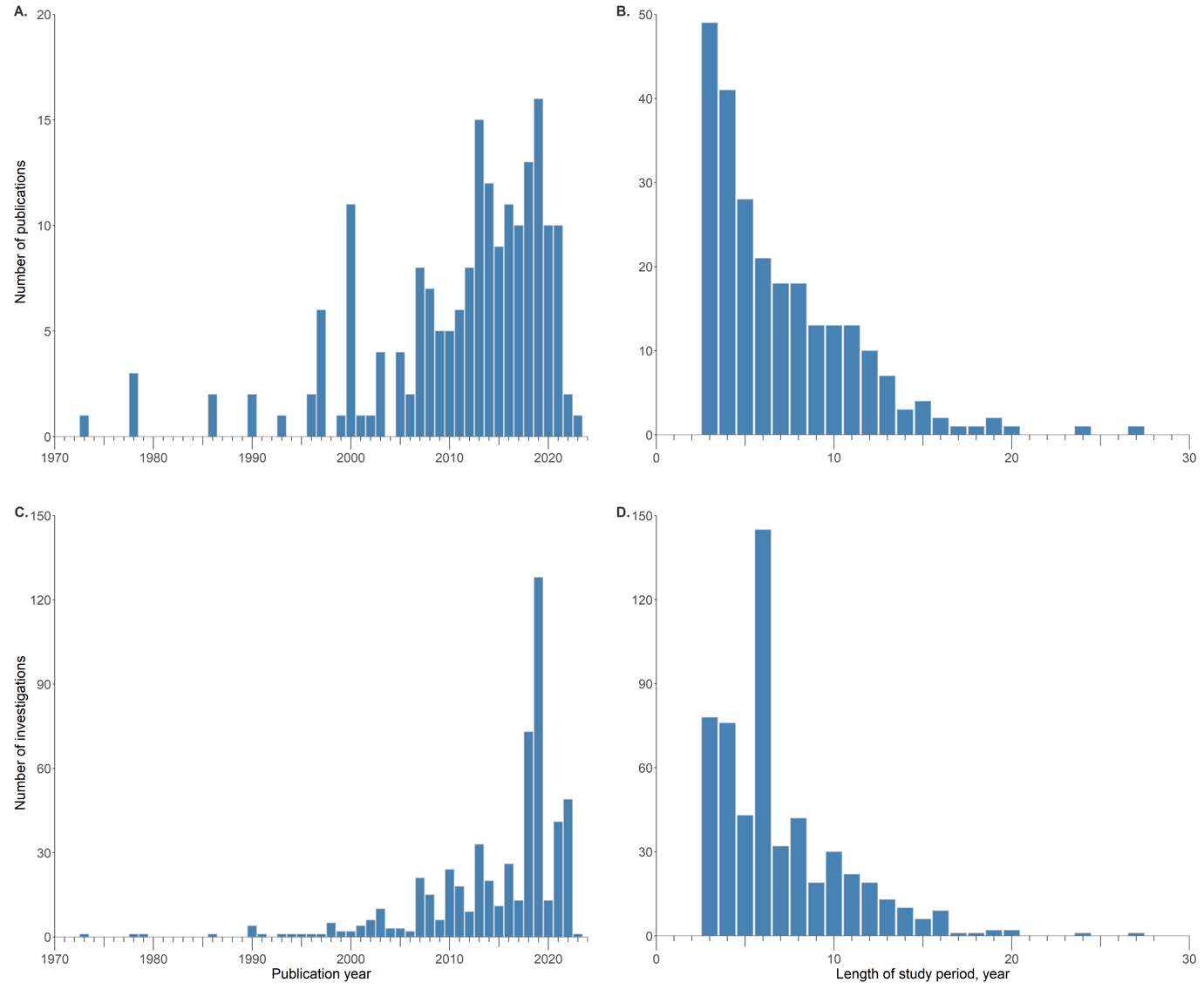

**Supplementary Figure 1** Annual numbers of publications and the investigations on RSV seasonality as of January 17, 2023 and the distribution of length of study period. (A) Annual number of included publications. (B) The distribution of length of study period for the included publications. (C) Annual number of investigations in the selected publications. (D) The distribution of length of study periods for the investigations in the selected publications.

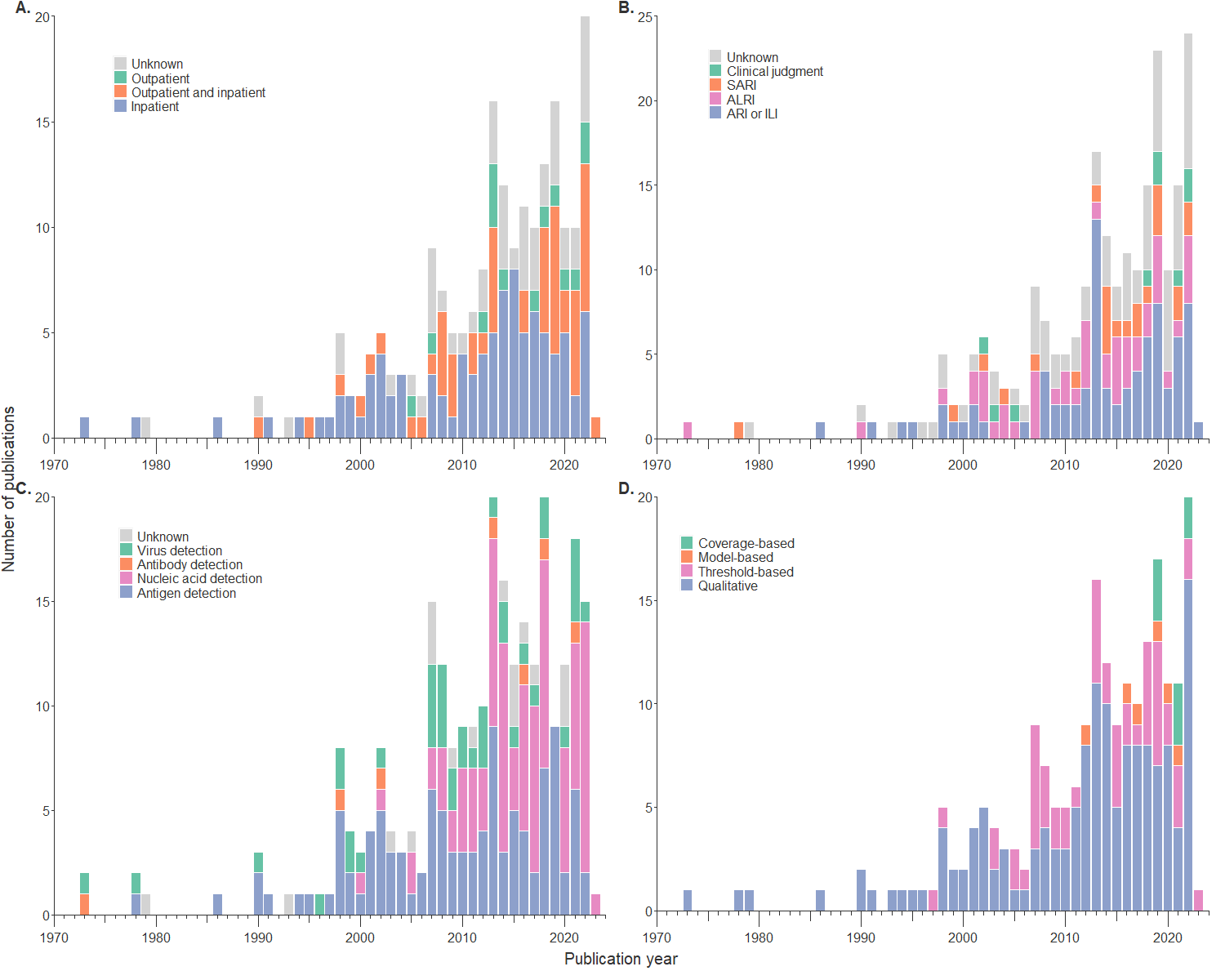

**Supplementary Figure 2.** The distribution of case sources (A), case definitions (B), testing methods (C) and analytic methods (D) adopted in the included publications by publication year.

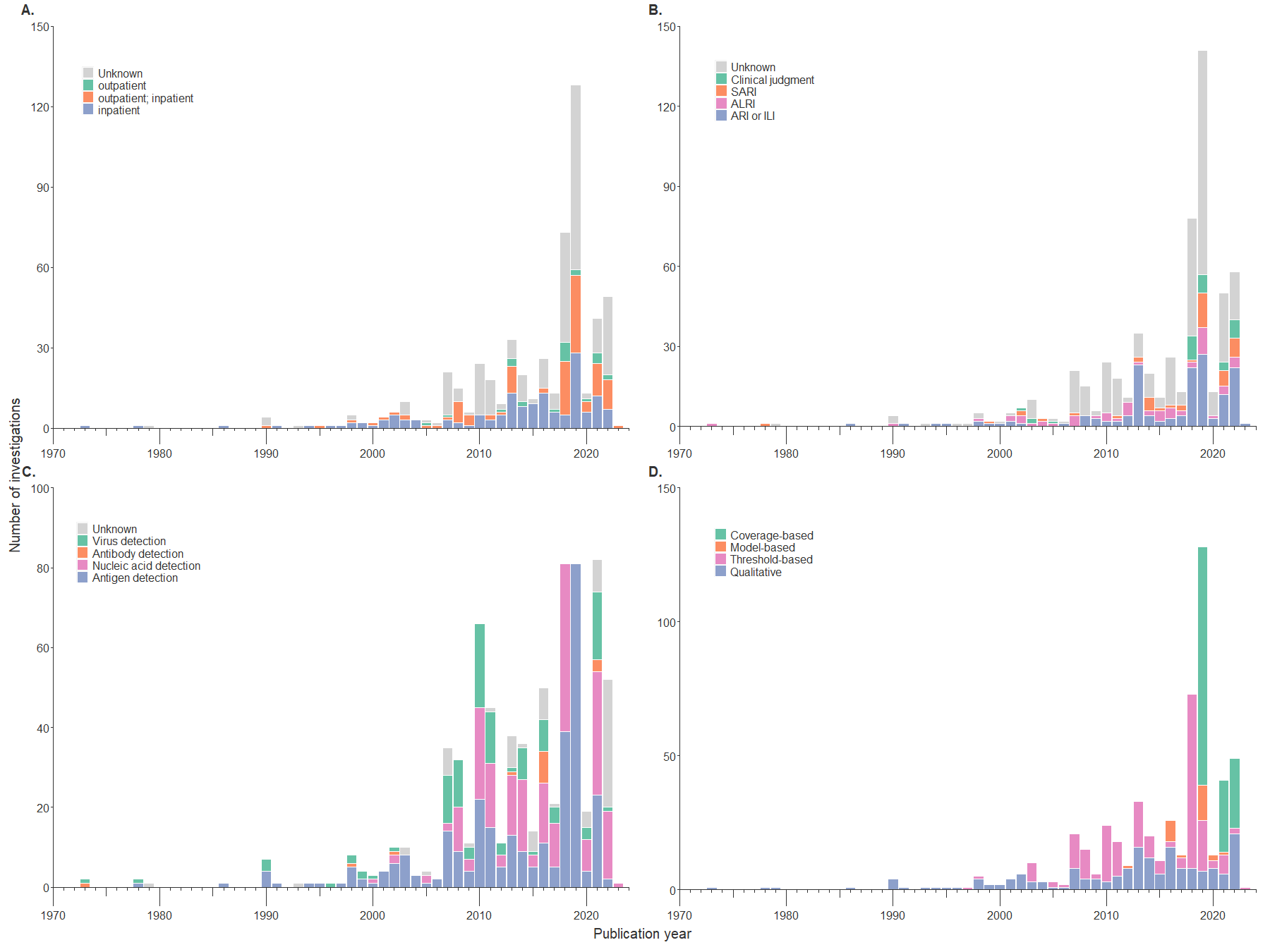

**Supplementary Figure 3.** The distribution of case sources, case definitions, testing methods and analysis methods adopted by the investigations in the selected publications by the publication year. (A) The distribution of case sources. (B) The distribution of case definitions.(C) The distribution of testing methods. (D) The distribution of analysis methods.

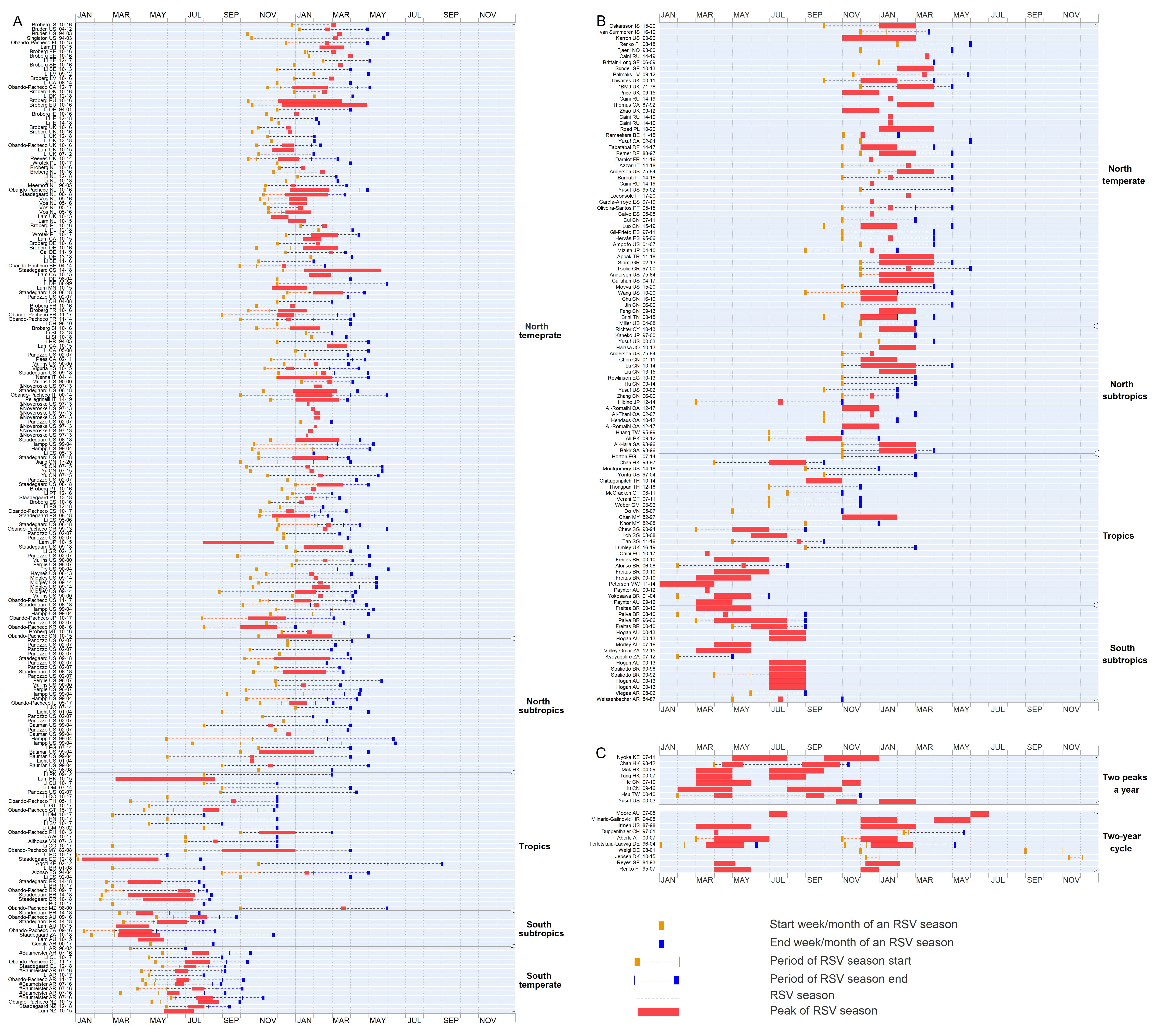

***** The title of this publication was not available, so we used the publisher instead.

& The 95% confidence interval of estimates of season timing were plotted.

\# The IQR (interquatile range) of estimates of RSV season timing were plotted.

$ The estimate of RSV season was associated with multiple countries, only the first country was shown.

**Supplementary Figure 4.** The reported time of the start, peak and/or end of RSV seasons from the investigations with the study period for 3 years or longer. Black lines denote the duration of the RSV season, red bars represent the peak period of reported RSV seasons. Orange segments indicate the reported week/month of the RSV season start/end, with the extended dashed line indicating the season start (to the right) or end (to the left) reported as a time period. The studies were ordered according to the centre latitude of the study site from the north (top) to the south (bottom). Estimated timing of RSV seasons for investigations classified as “One RSV peak a year” by quantitative methods (A) or by qualitative methods (B), and investigations classified as “Two-year cycle” and “Two RSV peaks a year” by either quantitative or qualitative methods (C).

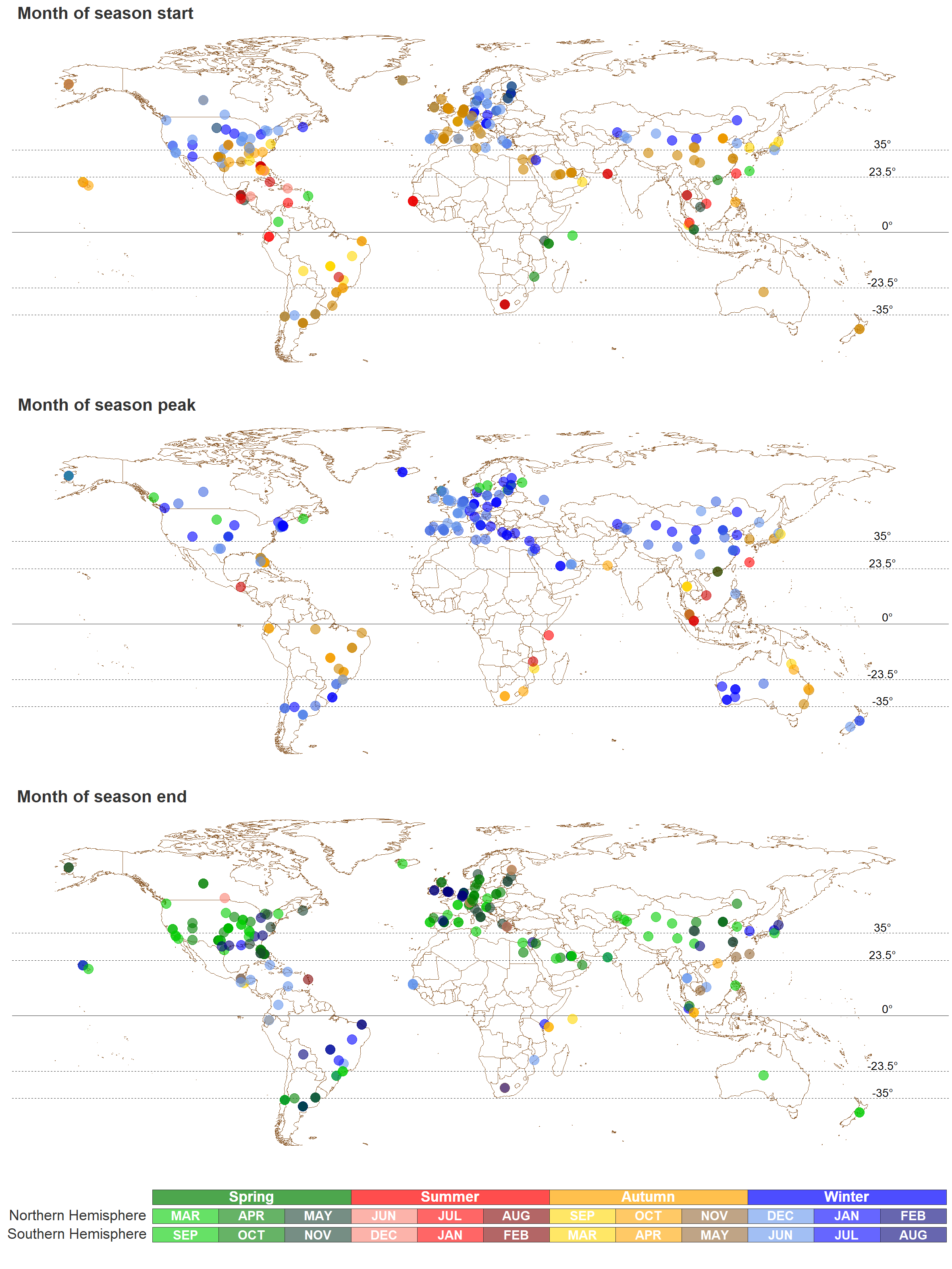
**Supplementary Figure 5.** Global map of the estimates on the start, peak, and end of RSV season from the included publications. (A) The start of RSV seasons. (B) The peak of RSV seasons. (C) The end of RSV seasons.

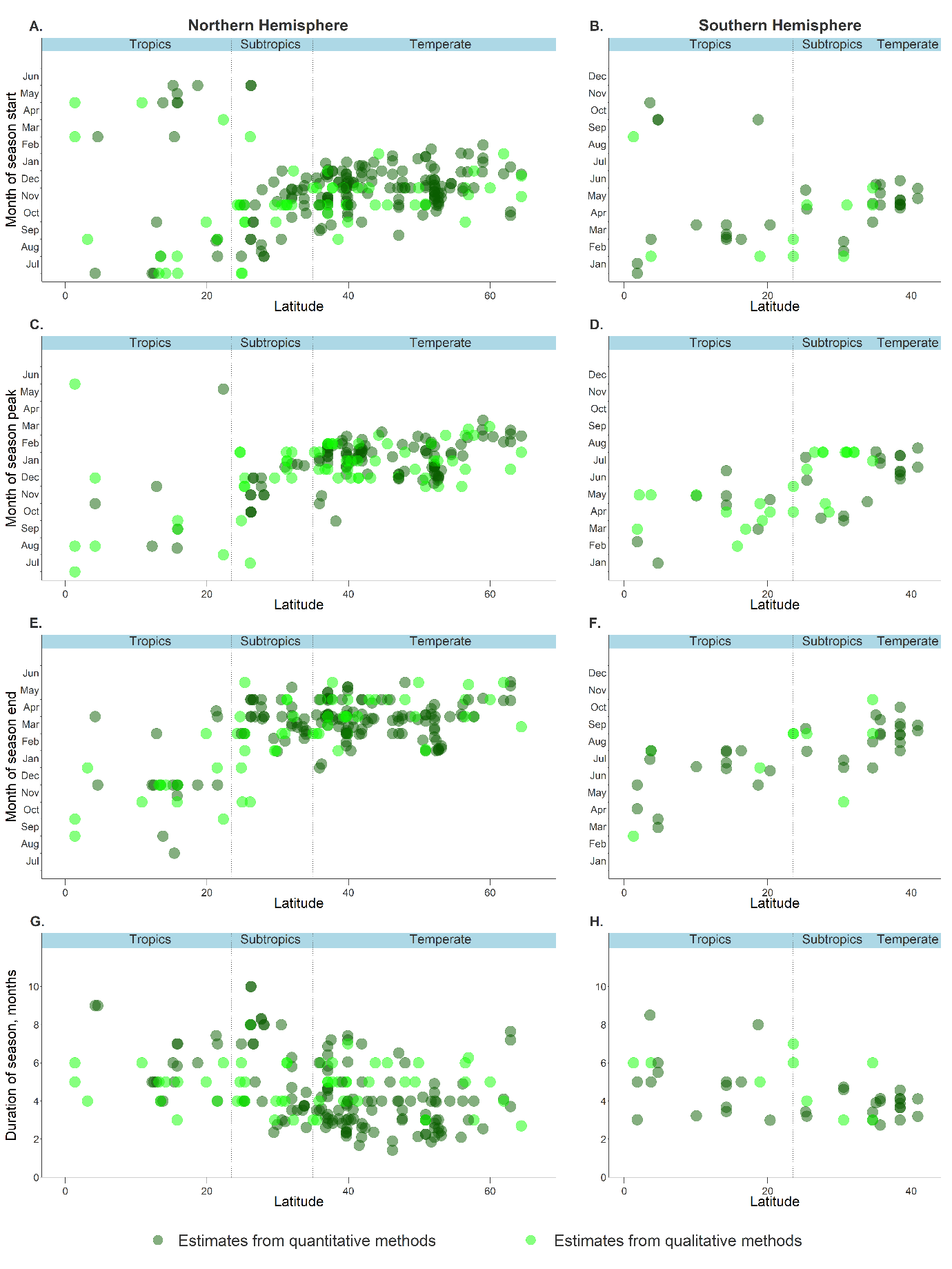
**Supplementary Figure 6.** The timing (the start, peak, and end) and duration of RSV seasons extracted from the included publications with the latitudes of the study sites. The start of RSV seasons with the latitudes in the Northern Hemisphere (A) and Southern Hemisphere (B). The peak of RSV seasons with the latitudes in the Northern Hemisphere (C) and Southern Hemisphere (D). The end of RSV seasons against the latitudes in the Northern Hemisphere (E) and Southern Hemisphere (F). The duration of RSV seasons against the latitudes in the Northern Hemisphere (G) and Southern Hemisphere (H). The dark green points represent the estimates using quantitative methods, and the light green points are for the estimates from qualitative approaches.

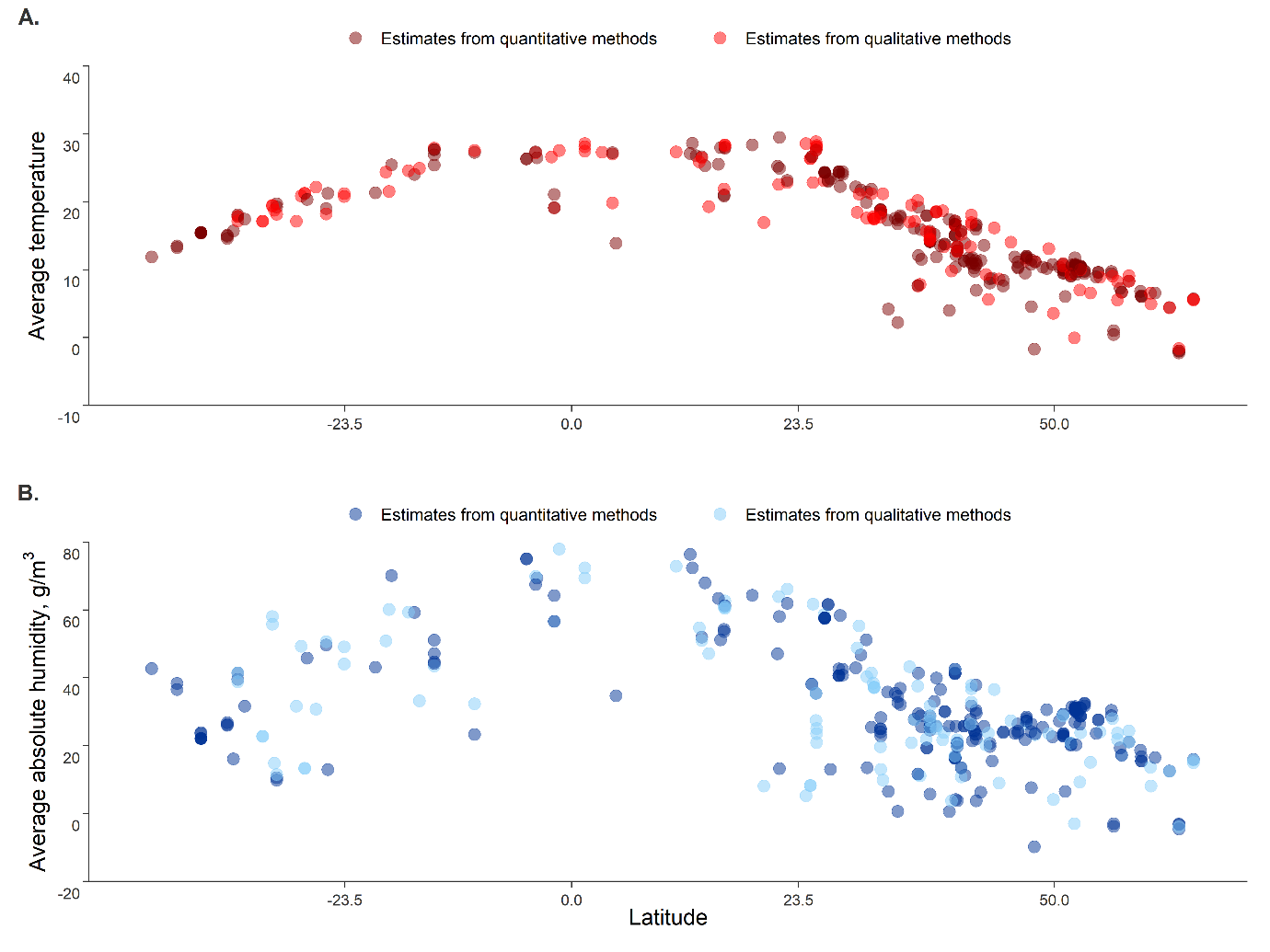
**Supplementary Figure 7.** The daily average mean temperature and daily average mean absolute humidity of the study sites during the study period by the latitudes of the study sites.

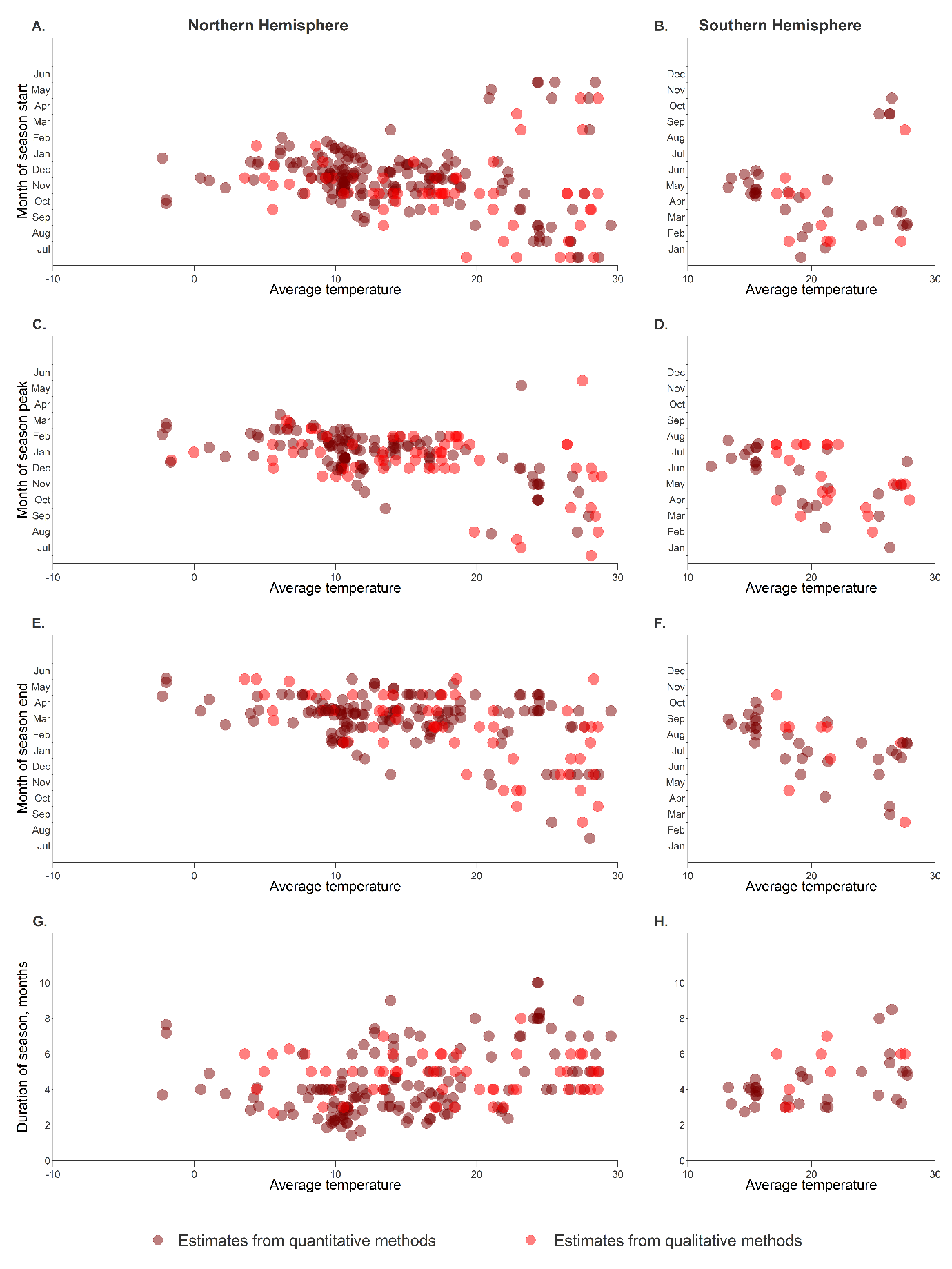

**Supplementary Figure 8.** The timing (the start, peak, and end) and duration of RSV seasons extracted from the included publications agaist the daily average mean temperature of the study sites during the study period. The start month of RSV seasons agaist the daily average mean temperature in the Northern Hemisphere (A) and Southern Hemisphere (B). The peak month of RSV seasons agaist the daily average mean temperature in the Northern Hemisphere (C) and Southern Hemisphere (D). The end month of RSV seasons agaist the daily average mean temperature in the Northern Hemisphere (E) and Southern Hemisphere (F). The duration of RSV seasons in months agaist the daily average mean temperature in the Northern Hemisphere (G) and Southern Hemisphere (H). The dark red points represent the estimates using quantitative methods, and the light red points are for the estimates from qualitative approaches.

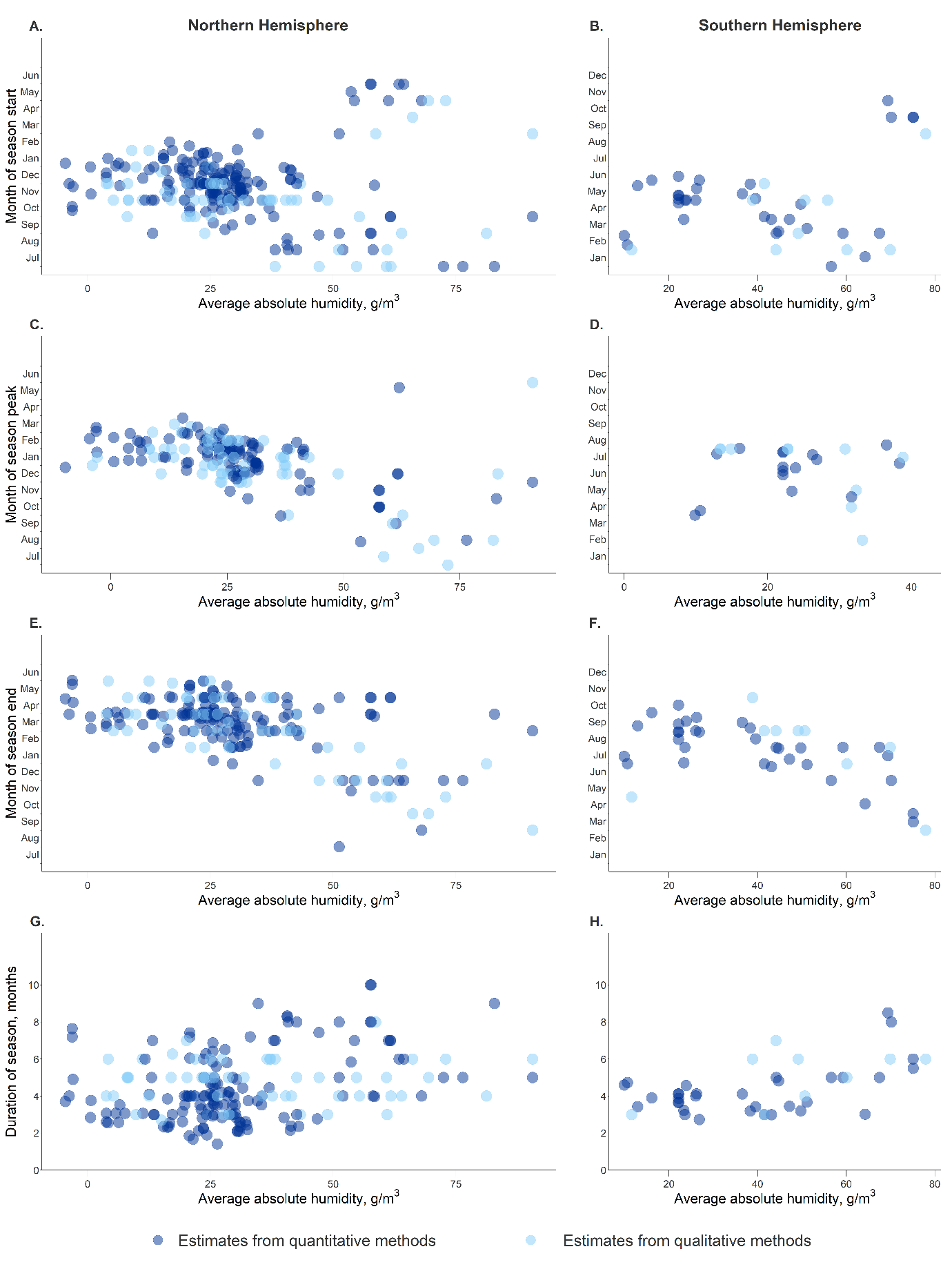
**Supplementary Figure 9.** The timing (the start, peak, and end) and duration of RSV seasons extracted from the included publications against the daily average mean absolute humidity of the study sites during the study period. The start of RSV seasons agaist the average absolute humidity in the Northern Hemisphere (A) and Southern Hemisphere (B). The peak of RSV seasons agaist the daily average mean absolute humidity in the Northern Hemisphere (C) and Southern Hemisphere (D). The end of RSV seasons agaist the daily average mean absolute humidity in the Northern Hemisphere (E). and Southern Hemisphere (F). The duration of RSV seasons against the daily average mean absolute humidity in the Northern Hemisphere (G) and Southern Hemisphere (H). The dark blue points represent the estimates using quantitative methods , and the light blue points are for the estimates from qualitative approaches.

**SUPPLEMENTARY TABLES**

**Supplementary Table 1**. Description of the sites and study characteristics of the associated investigations included in the systematic review.

| **Publication** | **Country** | **Location** | **Longitude and latitude** | **Study period** | **Case source** | **Case definition** | **Testing method** | **Season method** | **Season scale** |
| --- | --- | --- | --- | --- | --- | --- | --- | --- | --- |
| Aberle, 2008 [[6]](https://paperpile.com/c/zPIKrB/xZQ74) | Austria | Vienna | 16·4, 48·2 | 2000 - 2007 | inpatient | ARI or ILI | Nucleic acid detection | Qualitative | Month |
| Agoti, 2015 [[7]](https://paperpile.com/c/zPIKrB/HZLo) | Kenya | Kilifi | 39·9, -3·6 | 2002 - 2012 | inpatient | ALRI | Antigen detection; Nucleic acid detection | Threshold-based | Month |
| AI-Assam, 2009 [[8]](https://paperpile.com/c/zPIKrB/tfn1D) | Canada | nationwide | -106·3, 56·1 | 2005 - 2008 | outpatient; inpatient | ARI or ILI | Virus detection; Antigen detection; Nucleic acid detection | Qualitative | Month |
| AI-Assam, 2009 [[8]](https://paperpile.com/c/zPIKrB/tfn1D) | Canada | Nova Scotia | -63·7, 44·7 | 2005 - 2008 | outpatient; inpatient | ARI or ILI | Virus detection; Antigen detection; Nucleic acid detection | Qualitative | Month |
| AI-Romaihi, 2019 [[9]](https://paperpile.com/c/zPIKrB/rqUb) | Qatar | Doha | 51·5, 25·3 | 2012 - 2017 | outpatient | ARI or ILI | Nucleic acid detection | Qualitative | Month |
| AI-Romaihi, 2020 [[10]](https://paperpile.com/c/zPIKrB/ohzb) | Qatar | nationwide | 51·2, 25·4 | 2012 - 2017 | outpatient | ARI or ILI | Nucleic acid detection | Qualitative | Month |
| AI-Thani, 2008 [[11]](https://paperpile.com/c/zPIKrB/x39d) | Qatar | nationwide | 51·2, 25·4 | 2002 - 2007 | outpatient; inpatient | ARI or ILI | Antigen detection | Qualitative | Month |
| Al-Hajja, 1998 [[12]](https://paperpile.com/c/zPIKrB/LxVi) | Saudi Arabia | Riyadh | 46·7, 24·7 | 1993 - 1996 | Unknown | ARI or ILI | Antigen detection | Qualitative | Month |
| Ali, 2017 [[13]](https://paperpile.com/c/zPIKrB/QU8n) | Pakistan | Karachi | 67, 24·9 | 2009 - 2012 | inpatient | ARI or ILI | Nucleic acid detection | Qualitative | Month |
| Alonso, 2007 [[14]](https://paperpile.com/c/zPIKrB/9fNn0) | Spain | Sacyl | 41·7, -4·7 | 1992 - 2004 | inpatient | ALRI | Antigen detection | Threshold-based | Month |
| Alonso, 2012 [[15]](https://paperpile.com/c/zPIKrB/YD3c) | Brazil | Fortaleza | -38·5, -3·7 | 2006 - 2008 | Unknown | ARI or ILI | Antigen detection | Qualitative | Month |
| Althouse, 2018 [[16]](https://paperpile.com/c/zPIKrB/tl6A9) | Vietnam | Nha Trang | 109·2, 12·3 | 2007 - 2012 | inpatient | ARI or ILI | Nucleic acid detection | Threshold-based | Month |
| Ambrose, 2019 [[17]](https://paperpile.com/c/zPIKrB/gWGAj) | USA | nationwide | -95·7, 37·1 | 2011 - 2016 | Unknown | Unknown | Virus detection; Antigen detection; Nucleic acid detection | Threshold-based | Week |
| Ampofo, 2008 [[18]](https://paperpile.com/c/zPIKrB/jlUM) | USA | Utah | -111·1, 39·3 | 2001 - 2007 | outpatient; inpatient | ARI or ILI | Virus detection; Antigen detection | Qualitative | Month |
| Anderson, 1990 [[19]](https://paperpile.com/c/zPIKrB/eub9) | USA | Colorado, Pennsylvania, and Washington | -100·7, 44·3 | 1975 - 1984 | Unknown | Unknown | Virus detection; Antigen detection | Qualitative | Month |
| Anderson, 1990 [[19]](https://paperpile.com/c/zPIKrB/eub9) | USA | nationwide | -95·7, 37·1 | 1975 - 1984 | Unknown | Unknown | Virus detection; Antigen detection | Qualitative | Month |
| Anderson, 1990 [[19]](https://paperpile.com/c/zPIKrB/eub9) | USA | Texas state | -99·9, 32 | 1975 - 1984 | Unknown | Unknown | Virus detection; Antigen detection | Qualitative | Month |
| Appak, 2019 [[20]](https://paperpile.com/c/zPIKrB/iCtM) | Turkey | Izmir | 27·1, 38·4 | 2011 - 2018 | outpatient; inpatient | ARI or ILI | Nucleic acid detection | Qualitative | Month |
| Arnott, 2011 [[21]](https://paperpile.com/c/zPIKrB/NNGa) | Cambodia | nationwide | 105, 12·6 | 2005 - 2009 | inpatient | ALRI | Nucleic acid detection | Qualitative | Month |
| Assink, 2009 [[22]](https://paperpile.com/c/zPIKrB/Z8Prv) | The Netherlands | nationwide | 5·3, 52·1 | 1998 - 2006 | Unknown | Unknown | Unknown | Threshold-based | Week |
| Azzari, 2021 [[23]](https://paperpile.com/c/zPIKrB/Ioh4) | Italy | Lombardy | 9·8, 45·5 | 2014 - 2018 | outpatient | ARI or ILI | Unknown | Qualitative | Month |
| Bakir, 1998 [[24]](https://paperpile.com/c/zPIKrB/PBuo) | Saudi Arabia | Riyadh | 46·7, 24·7 | 1993 - 1996 | inpatient | ARI or ILI | Virus detection; Antigen detection | Qualitative | Month |
| Balmaks, 2014 [[25]](https://paperpile.com/c/zPIKrB/Xgw6) | Latvia | Riga | 24·1, 56·9 | 2009 - 2012 | inpatient | ALRI | Nucleic acid detection | Qualitative | Week |
| Bandeira, 2022 [[26]](https://paperpile.com/c/zPIKrB/avkSI) | Portugal | nationwide | -9·2, 39·7 | 2015 - 2018 | inpatient | Unknown | Unknown | Qualitative | Week |
| Barbati, 2020 [[27]](https://paperpile.com/c/zPIKrB/uycl) | Italy | Tuscany | 11·2, 43·8 | 2014 - 2019 | inpatient | Unknown | Nucleic acid detection | Qualitative | Month |
| Bauman, 2007 [[28]](https://paperpile.com/c/zPIKrB/T5MU6) | USA | Central, Florida state | -81·9, 28·1 | 1999 - 2004 | Unknown | Unknown | Unknown | Threshold-based | Month |
| Bauman, 2007 [[28]](https://paperpile.com/c/zPIKrB/T5MU6) | USA | Florida state | -81·5, 27·7 | 1999 - 2004 | Unknown | Unknown | Unknown | Threshold-based | Month |
| Bauman, 2007 [[28]](https://paperpile.com/c/zPIKrB/T5MU6) | USA | North, Florida state | -80·2, 26·2 | 1999 - 2004 | Unknown | Unknown | Unknown | Threshold-based | Month |
| Bauman, 2007 [[28]](https://paperpile.com/c/zPIKrB/T5MU6) | USA | Southeast, Florida state | -80·4, 26·2 | 1999 - 2004 | Unknown | Unknown | Unknown | Threshold-based | Month |
| Bauman, 2007 [[28]](https://paperpile.com/c/zPIKrB/T5MU6) | USA | Southwest, Florida state | -81·9, 26·6 | 1999 - 2004 | Unknown | Unknown | Unknown | Threshold-based | Month |
| Baumeister, 2019 [[29]](https://paperpile.com/c/zPIKrB/StFDy) | Argentina | Central | -63·6, -38·4 | 2007 - 2016 | outpatient; inpatient | ARI or ILI; SARI; Clinical judgment | Antigen detection; Nucleic acid detection | Threshold-based | Week |
| Baumeister, 2019 [[29]](https://paperpile.com/c/zPIKrB/StFDy) | Argentina | Cuyo | -67·4, -35·1 | 2007 - 2016 | outpatient; inpatient | ARI or ILI; SARI; Clinical judgment | Antigen detection; Nucleic acid detection | Threshold-based | Week |
| Baumeister, 2019 [[29]](https://paperpile.com/c/zPIKrB/StFDy) | Argentina | nationwide | -63·6, -38·4 | 2007 - 2016 | outpatient; inpatient | ARI or ILI; SARI; Clinical judgment | Antigen detection; Nucleic acid detection | Threshold-based | Week |
| Baumeister, 2019 [[29]](https://paperpile.com/c/zPIKrB/StFDy) | Argentina | Northeast | -63·6, -38·4 | 2007 - 2016 | outpatient; inpatient | ARI or ILI; SARI; Clinical judgment | Antigen detection; Nucleic acid detection | Threshold-based | Week |
| Baumeister, 2019 [[29]](https://paperpile.com/c/zPIKrB/StFDy) | Argentina | Northwest | -63·6, -38·4 | 2007 - 2016 | outpatient; inpatient | ARI or ILI; SARI; Clinical judgment | Antigen detection; Nucleic acid detection | Threshold-based | Week |
| Baumeister, 2019 [[29]](https://paperpile.com/c/zPIKrB/StFDy) | Argentina | South | -63·6, -38·4 | 2007 - 2016 | outpatient; inpatient | ARI or ILI; SARI; Clinical judgment | Antigen detection; Nucleic acid detection | Threshold-based | Week |
| Berner, 2001 [[30]](https://paperpile.com/c/zPIKrB/EIBi) | Germany | Freiburg | 7·8, 48 | 1988 - 1998 | inpatient | ARI or ILI | Antigen detection | Qualitative | Month |
| Billard, 2022 [[31]](https://paperpile.com/c/zPIKrB/aUflC) | Brazil | nationwide | -51·9, -14·2 | 2017 - 2019 | Unknown | Unknown | Unknown | Coverage-based | Week |
| Billard, 2022 [[31]](https://paperpile.com/c/zPIKrB/aUflC) | Canada | nationwide | -106·3, 56·1 | 2017 - 2019 | Unknown | Unknown | Unknown | Coverage-based | Week |
| Billard, 2022 [[31]](https://paperpile.com/c/zPIKrB/aUflC) | Chile | nationwide | -71·5, -35·7 | 2017 - 2019 | Unknown | Unknown | Unknown | Coverage-based | Week |
| Billard, 2022 [[31]](https://paperpile.com/c/zPIKrB/aUflC) | France | nationwide | 2·7, 47·1 | 2017 - 2019 | Unknown | Unknown | Unknown | Coverage-based | Week |
| Billard, 2022 [[31]](https://paperpile.com/c/zPIKrB/aUflC) | Israel | nationwide | 34·9, 31 | 2017 - 2019 | Unknown | Unknown | Unknown | Coverage-based | Week |
| Billard, 2022 [[31]](https://paperpile.com/c/zPIKrB/aUflC) | Japan | nationwide | 138·3, 36·2 | 2017 - 2019 | Unknown | Unknown | Unknown | Coverage-based | Week |
| Billard, 2022 [[31]](https://paperpile.com/c/zPIKrB/aUflC) | South Africa | nationwide | 22·9, -30·6 | 2017 - 2019 | Unknown | Unknown | Unknown | Coverage-based | Week |
| Billard, 2022 [[31]](https://paperpile.com/c/zPIKrB/aUflC) | South Korea | nationwide | 127·8, 35·9 | 2017 - 2019 | Unknown | Unknown | Unknown | Coverage-based | Week |
| Billard, 2022 [[31]](https://paperpile.com/c/zPIKrB/aUflC) | Taiwan | nationwide | 121, 23·7 | 2017 - 2019 | Unknown | Unknown | Unknown | Coverage-based | Week |
| Billard, 2022 [[31]](https://paperpile.com/c/zPIKrB/aUflC) | The Netherlands | nationwide | 5·3, 52·1 | 2017 - 2019 | Unknown | Unknown | Unknown | Coverage-based | Week |
| Billard, 2022 [[31]](https://paperpile.com/c/zPIKrB/aUflC) | USA | nationwide | -95·7, 37·1 | 2017 - 2019 | Unknown | Unknown | Unknown | Coverage-based | Week |
| BMJ 1979* [[32]](https://paperpile.com/c/zPIKrB/qwlJ) | United Kingdom | Scotland | -4·2, 56·5 | 1971 - 1978 | Unknown | Unknown | Unknown | Qualitative | Month |
| Boron, 2008 [[33]](https://paperpile.com/c/zPIKrB/80EEw) | USA | Midwest | -93·1, 41·9 | 2004 - 2007 | outpatient; inpatient | Unknown | Virus detection; Antigen detection; Nucleic acid detection | Threshold-based | Week |
| Boron, 2008 [[33]](https://paperpile.com/c/zPIKrB/80EEw) | USA | nationwide | -95·7, 37·1 | 2004 - 2007 | outpatient; inpatient | Unknown | Virus detection; Antigen detection; Nucleic acid detection | Threshold-based | Week |
| Boron, 2008 [[33]](https://paperpile.com/c/zPIKrB/80EEw) | USA | Northeast | -74·2, 43·3 | 2004 - 2007 | outpatient; inpatient | Unknown | Virus detection; Antigen detection; Nucleic acid detection | Threshold-based | Week |
| Boron, 2008 [[33]](https://paperpile.com/c/zPIKrB/80EEw) | USA | South | -99, 32 | 2004 - 2007 | outpatient; inpatient | Unknown | Virus detection; Antigen detection; Nucleic acid detection | Threshold-based | Week |
| Boron, 2008 [[33]](https://paperpile.com/c/zPIKrB/80EEw) | USA | West | -111, 37·1 | 2004 - 2007 | outpatient; inpatient | Unknown | Virus detection; Antigen detection; Nucleic acid detection | Threshold-based | Week |
| Brini, 2020 [[34]](https://paperpile.com/c/zPIKrB/71Jr) | Tunisia | Sousse | 10·6, 35·8 | 2003 - 2015 | inpatient | ALRI | Antigen detection | Qualitative | Month |
| Brittain-Long, 2011 [[35]](https://paperpile.com/c/zPIKrB/jYh9) | Sweden | Gothenburg | 12, 57·7 | 2006 - 2009 | outpatient; inpatient | Unknown | Nucleic acid detection | Qualitative | Month |
| Broberg, 2018 [[36]](https://paperpile.com/c/zPIKrB/Alr49) | Denmark | nationwide | 10·9, 55·9 | 2010 - 2016 | outpatient; inpatient | ARI or ILI; Clinical judgment | Virus detection; Antibody detection; Antigen detection; Nucleic acid detection | Threshold-based | Week |
| Broberg, 2018 [[36]](https://paperpile.com/c/zPIKrB/Alr49) | Estonia | nationwide | 25·5, 59 | 2010 - 2016 | outpatient | ARI or ILI | Virus detection; Antibody detection; Antigen detection; Nucleic acid detection | Threshold-based | Week |
| Broberg, 2018 [[36]](https://paperpile.com/c/zPIKrB/Alr49) | Estonia | nationwide | 25·5, 59 | 2010 - 2016 | outpatient; inpatient | ARI or ILI | Virus detection; Antibody detection; Antigen detection; Nucleic acid detection | Threshold-based | Week |
| Broberg, 2018 [[36]](https://paperpile.com/c/zPIKrB/Alr49) | Europe | nationwide | 15·3, 54·5 | 2010 - 2016 | outpatient; inpatient | ARI or ILI; Clinical judgment | Virus detection; Antibody detection; Antigen detection; Nucleic acid detection | Threshold-based | Week |
| Broberg, 2018 [[36]](https://paperpile.com/c/zPIKrB/Alr49) | Europe | nationwide | 15·3, 54·5 | 2010 - 2016 | outpatient; inpatient | ARI or ILI; Clinical judgment | Virus detection; Antibody detection; Antigen detection; Nucleic acid detection | Threshold-based | Week |
| Broberg, 2018 [[36]](https://paperpile.com/c/zPIKrB/Alr49) | France | nationwide | 2·7, 47·1 | 2010 - 2016 | outpatient | ARI or ILI | Virus detection; Antibody detection; Antigen detection; Nucleic acid detection | Threshold-based | Week |
| Broberg, 2018 [[36]](https://paperpile.com/c/zPIKrB/Alr49) | France | nationwide | 2·7, 47·1 | 2010 - 2016 | outpatient; inpatient | ARI or ILI | Virus detection; Antibody detection; Antigen detection; Nucleic acid detection | Threshold-based | Week |
| Broberg, 2018 [[36]](https://paperpile.com/c/zPIKrB/Alr49) | Germany | nationwide | 9·7, 50·9 | 2010 - 2016 | outpatient | ARI or ILI | Virus detection; Antibody detection; Antigen detection; Nucleic acid detection | Threshold-based | Week |
| Broberg, 2018 [[36]](https://paperpile.com/c/zPIKrB/Alr49) | Germany | nationwide | 9·7, 50·9 | 2010 - 2016 | outpatient; inpatient | ARI or ILI | Virus detection; Antibody detection; Antigen detection; Nucleic acid detection | Threshold-based | Week |
| Broberg, 2018 [[36]](https://paperpile.com/c/zPIKrB/Alr49) | Iceland | nationwide | -21·1, 64·4 | 2010 - 2016 | outpatient; inpatient | Clinical judgment | Virus detection; Antibody detection; Antigen detection; Nucleic acid detection | Threshold-based | Week |
| Broberg, 2018 [[36]](https://paperpile.com/c/zPIKrB/Alr49) | Ireland | nationwide | -7·4, 53·1 | 2010 - 2016 | outpatient; inpatient | Clinical judgment | Virus detection; Antibody detection; Antigen detection; Nucleic acid detection | Threshold-based | Week |
| Broberg, 2018 [[36]](https://paperpile.com/c/zPIKrB/Alr49) | Latvia | nationwide | 24·4, 56·8 | 2010 - 2016 | outpatient; inpatient | Clinical judgment | Virus detection; Antibody detection; Antigen detection; Nucleic acid detection | Threshold-based | Week |
| Broberg, 2018 [[36]](https://paperpile.com/c/zPIKrB/Alr49) | Malta | nationwide | 14·5, 35·9 | 2010 - 2016 | outpatient; inpatient | ARI or ILI | Virus detection; Antibody detection; Antigen detection; Nucleic acid detection | Threshold-based | Week |
| Broberg, 2018 [[36]](https://paperpile.com/c/zPIKrB/Alr49) | Poland | nationwide | 19·3, 51·7 | 2010 - 2016 | outpatient; inpatient | ARI or ILI | Virus detection; Antibody detection; Antigen detection; Nucleic acid detection | Threshold-based | Week |
| Broberg, 2018 [[36]](https://paperpile.com/c/zPIKrB/Alr49) | Portugal | nationwide | -9·2, 39·7 | 2010 - 2016 | outpatient; inpatient | ARI or ILI | Virus detection; Antibody detection; Antigen detection; Nucleic acid detection | Threshold-based | Week |
| Broberg, 2018 [[36]](https://paperpile.com/c/zPIKrB/Alr49) | Slovenia | nationwide | 14·9, 46·2 | 2010 - 2016 | outpatient | ARI or ILI | Virus detection; Antibody detection; Antigen detection; Nucleic acid detection | Threshold-based | Week |
| Broberg, 2018 [[36]](https://paperpile.com/c/zPIKrB/Alr49) | Spain | nationwide | -3·3, 39·7 | 2010 - 2016 | outpatient; inpatient | Clinical judgment | Virus detection; Antibody detection; Antigen detection; Nucleic acid detection | Threshold-based | Week |
| Broberg, 2018 [[36]](https://paperpile.com/c/zPIKrB/Alr49) | Sweden | nationwide | 15·4, 58·9 | 2010 - 2016 | outpatient; inpatient | Clinical judgment | Virus detection; Antibody detection; Antigen detection; Nucleic acid detection | Threshold-based | Week |
| Broberg, 2018 [[36]](https://paperpile.com/c/zPIKrB/Alr49) | The Netherlands | nationwide | 5·3, 52·1 | 2010 - 2016 | outpatient | ARI or ILI | Virus detection; Antibody detection; Antigen detection; Nucleic acid detection | Threshold-based | Week |
| Broberg, 2018 [[36]](https://paperpile.com/c/zPIKrB/Alr49) | The Netherlands | nationwide | 5·3, 52·1 | 2010 - 2016 | outpatient; inpatient | ARI or ILI; Clinical judgment | Virus detection; Antibody detection; Antigen detection; Nucleic acid detection | Threshold-based | Week |
| Broberg, 2018 [[36]](https://paperpile.com/c/zPIKrB/Alr49) | United Kingdom | nationwide | -1·6, 52·7 | 2010 - 2016 | outpatient | ARI or ILI | Virus detection; Antibody detection; Antigen detection; Nucleic acid detection | Threshold-based | Week |
| Broberg, 2018 [[36]](https://paperpile.com/c/zPIKrB/Alr49) | United Kingdom | nationwide | -1·6, 52·7 | 2010 - 2016 | outpatient; inpatient | ARI or ILI | Virus detection; Antibody detection; Antigen detection; Nucleic acid detection | Threshold-based | Week |
| Bruden, 2015 [[37]](https://paperpile.com/c/zPIKrB/OpAug) | USA | YK Delta | -164·2, 62·9 | 1994 - 2012 | inpatient | ALRI | Virus detection; Antigen detection | Threshold-based | Week |
| Cai, 2022 [[38]](https://paperpile.com/c/zPIKrB/42Pvu) | Germany | nationwide | 9·7, 50·9 | 2011 - 2019 | outpatient | ARI or ILI | Nucleic acid detection | Threshold-based | Week |
| Caini, 2019 [[39]](https://paperpile.com/c/zPIKrB/WHKf) | Ecuador | nationwide | -78·2, -1·8 | 2010 - 2016 | outpatient; inpatient | ARI or ILI; SARI | Nucleic acid detection | Qualitative | Month |
| Caini, 2022 [[40]](https://paperpile.com/c/zPIKrB/DKVK) | Russia | Chita | 113·5, 52·1 | 2014 - 2019 | outpatient; inpatient | ARI or ILI; SARI | Nucleic acid detection | Qualitative | Month |
| Caini, 2022 [[40]](https://paperpile.com/c/zPIKrB/DKVK) | Russia | Kaliningrad | 20·5, 54·7 | 2014 - 2019 | outpatient; inpatient | ARI or ILI; SARI | Nucleic acid detection | Qualitative | Month |
| Caini, 2022 [[40]](https://paperpile.com/c/zPIKrB/DKVK) | Russia | Lipetsk | 39·6, 52·6 | 2014 - 2019 | outpatient; inpatient | ARI or ILI; SARI | Nucleic acid detection | Qualitative | Month |
| Caini, 2022 [[40]](https://paperpile.com/c/zPIKrB/DKVK) | Russia | St. Petersburg | 30·4, 59·9 | 2014 - 2019 | outpatient; inpatient | ARI or ILI; SARI | Nucleic acid detection | Qualitative | Month |
| Caini, 2022 [[40]](https://paperpile.com/c/zPIKrB/DKVK) | Russia | Vladivostok | 131·9, 43·1 | 2014 - 2019 | outpatient; inpatient | ARI or ILI; SARI | Nucleic acid detection | Qualitative | Month |
| Callahan, 2020 [[5]](https://paperpile.com/c/zPIKrB/o6J01) | USA | nationwide | -95·7, 37·1 | 2005 - 2018 | outpatient; inpatient | Unknown | Virus detection; Antigen detection; Nucleic acid detection | Model-based | Week |
| Callahan, 2020 [[5]](https://paperpile.com/c/zPIKrB/o6J01) | USA | nationwide | -95·7, 37·1 | 2005 - 2018 | outpatient; inpatient | Unknown | Virus detection; Antigen detection; Nucleic acid detection | Qualitative | Month |
| Callahan, 2020 [[5]](https://paperpile.com/c/zPIKrB/o6J01) | USA | Utah | -111·1, 39·3 | 2005 - 2018 | outpatient; inpatient | Unknown | Virus detection; Antigen detection; Nucleic acid detection | Model-based | Week |
| Calvo, 2010 [[41]](https://paperpile.com/c/zPIKrB/TCQq) | Spain | Leganés | -3·8, 40·3 | 2005 - 2008 | inpatient | ALRI | Nucleic acid detection | Qualitative | Month |
| Calvo, 2016 [[42]](https://paperpile.com/c/zPIKrB/PLZ0) | Spain | Leganés | -3·8, 40·3 | 2005 - 2013 | inpatient | ALRI | Nucleic acid detection | Qualitative | Month |
| Cattoir, 2018 [[43]](https://paperpile.com/c/zPIKrB/fNot) | Belgium | nationwide | 4·5, 50·5 | 2006 - 2016 | outpatient; inpatient | Unknown | Nucleic acid detection | Qualitative | Month |
| CDC, 2011 [[44]](https://paperpile.com/c/zPIKrB/u2aTv) | USA | Florida state | -81·5, 27·7 | 2007 - 2011 | Unknown | Unknown | Virus detection; Antigen detection; Nucleic acid detection | Threshold-based | Week |
| CDC, 2011 [[44]](https://paperpile.com/c/zPIKrB/u2aTv) | USA | HHS1 | 71·1, 42·4 | 2007 - 2011 | Unknown | Unknown | Virus detection; Antigen detection; Nucleic acid detection | Threshold-based | Week |
| CDC, 2011 [[44]](https://paperpile.com/c/zPIKrB/u2aTv) | USA | HHS10 | 122·3, 47·6 | 2007 - 2011 | Unknown | Unknown | Virus detection; Antigen detection; Nucleic acid detection | Threshold-based | Week |
| CDC, 2011 [[44]](https://paperpile.com/c/zPIKrB/u2aTv) | USA | HHS2 | 74, 40·7 | 2007 - 2011 | Unknown | Unknown | Virus detection; Antigen detection; Nucleic acid detection | Threshold-based | Week |
| CDC, 2011 [[44]](https://paperpile.com/c/zPIKrB/u2aTv) | USA | HHS3 | 75·2, 40 | 2007 - 2011 | Unknown | Unknown | Virus detection; Antigen detection; Nucleic acid detection | Threshold-based | Week |
| CDC, 2011 [[44]](https://paperpile.com/c/zPIKrB/u2aTv) | USA | HHS4 | 84·4, 33·8 | 2007 - 2011 | Unknown | Unknown | Virus detection; Antigen detection; Nucleic acid detection | Threshold-based | Week |
| CDC, 2011 [[44]](https://paperpile.com/c/zPIKrB/u2aTv) | USA | HHS5 | 87·6, 41·9 | 2007 - 2011 | Unknown | Unknown | Virus detection; Antigen detection; Nucleic acid detection | Threshold-based | Week |
| CDC, 2011 [[44]](https://paperpile.com/c/zPIKrB/u2aTv) | USA | HHS6 | 96·8, 32·8 | 2007 - 2011 | Unknown | Unknown | Virus detection; Antigen detection; Nucleic acid detection | Threshold-based | Week |
| CDC, 2011 [[44]](https://paperpile.com/c/zPIKrB/u2aTv) | USA | HHS7 | 94·6, 39·1 | 2007 - 2011 | Unknown | Unknown | Virus detection; Antigen detection; Nucleic acid detection | Threshold-based | Week |
| CDC, 2011 [[44]](https://paperpile.com/c/zPIKrB/u2aTv) | USA | HHS8 | 105, 39·7 | 2007 - 2011 | Unknown | Unknown | Virus detection; Antigen detection; Nucleic acid detection | Threshold-based | Week |
| CDC, 2011 [[44]](https://paperpile.com/c/zPIKrB/u2aTv) | USA | HHS9 | 122·4, 37·8 | 2007 - 2011 | Unknown | Unknown | Virus detection; Antigen detection; Nucleic acid detection | Threshold-based | Week |
| CDC, 2011 [[44]](https://paperpile.com/c/zPIKrB/u2aTv) | USA | nationwide | -95·7, 37·1 | 2007 - 2011 | Unknown | Unknown | Virus detection; Antigen detection; Nucleic acid detection | Threshold-based | Week |
| CDC, 2011 [[44]](https://paperpile.com/c/zPIKrB/u2aTv) | USA | nationwide | -95·7, 37·1 | 2007 - 2011 | Unknown | Unknown | Virus detection; Antigen detection; Nucleic acid detection | Threshold-based | Week |
| Chan, 1999 [[45]](https://paperpile.com/c/zPIKrB/o9nC) | Hong Kong | nationwide | 114·2, 22·3 | 1993 - 1997 | inpatient | SARI | Virus detection; Antigen detection | Qualitative | Month |
| Chan, 2002 [[46]](https://paperpile.com/c/zPIKrB/qofI) | Malaysia | nationwide | 102, 4·2 | 1982 - 1997 | inpatient | ALRI | Virus detection; Antigen detection | Qualitative | Month |
| Chan, 2015 [[47]](https://paperpile.com/c/zPIKrB/sdUN2) | Hong Kong | nationwide | 114·2, 22·3 | 1998 - 2012 | inpatient | ARI or ILI | Antigen detection | Threshold-based | Week |
| Chen, 2013 [[48]](https://paperpile.com/c/zPIKrB/A33W) | China | Suzhou | 120·6, 31·3 | 2001 - 2011 | inpatient | SARI | Nucleic acid detection | Qualitative | Month |
| Chew, 1998 [[49]](https://paperpile.com/c/zPIKrB/bzzz) | Singapore | nationwide | 103·8, 1·4 | 1990 - 1994 | outpatient; inpatient | Unknown | Virus detection; Antibody detection; Antigen detection | Qualitative | Month |
| Chi, 2011 [[50]](https://paperpile.com/c/zPIKrB/osdms) | Taiwan | nationwide | 121, 23·7 | 2004 - 2007 | inpatient | Unknown | Unknown | Qualitative | Month |
| Chittaganpitch, 2018 [[51]](https://paperpile.com/c/zPIKrB/RFay) | Thailand | nationwide | 101, 15·9 | 2010 - 2014 | outpatient; inpatient | ARI or ILI; SARI | Nucleic acid detection | Qualitative | Month |
| Choudhary, 2013 [[52]](https://paperpile.com/c/zPIKrB/H1R9) | India | nationwide | 79, 20·6 | 2009 - 2012 | outpatient; inpatient | ARI or ILI; SARI | Nucleic acid detection | Qualitative | Month |
| Choudhary, 2013 [[52]](https://paperpile.com/c/zPIKrB/H1R9) | India | Pune | 73·9, 18·5 | 2009 - 2012 | outpatient; inpatient | ARI or ILI; SARI | Nucleic acid detection | Qualitative | Month |
| Chu, 2022 [[53]](https://paperpile.com/c/zPIKrB/qC9rq) | China | Jinan | 117, 36·7 | 2016 - 2019 | inpatient | ALRI | Nucleic acid detection | Qualitative | Month |
| Cui, 2013 [[54]](https://paperpile.com/c/zPIKrB/IMnZ) | China | Beijing | 116·4, 39·9 | 2007 - 2012 | inpatient | ARI or ILI | Antigen detection | Qualitative | Month |
| Cui, 2016 [[55]](https://paperpile.com/c/zPIKrB/mcfA) | China | Eastern China | 104·2, 35·9 | 2009 - 2013 | inpatient | ALRI | Nucleic acid detection | Qualitative | Month |
| Cui, 2016 [[55]](https://paperpile.com/c/zPIKrB/mcfA) | China | nationwide | 104·2, 35·9 | 2009 - 2013 | inpatient | ALRI | Nucleic acid detection | Qualitative | Month |
| Darniot, 2018 [[56]](https://paperpile.com/c/zPIKrB/Jb43) | France | Burgundy | 4·4, 47·1 | 2011 - 2016 | inpatient | ARI or ILI | Antigen detection; Nucleic acid detection | Qualitative | Week |
| De Conto, 2019 [[57]](https://paperpile.com/c/zPIKrB/vNcVL) | Italy | Parma | 10·3, 44·8 | 2012 - 2015 | outpatient; inpatient | ARI or ILI | Antigen detection; Nucleic acid detection | Threshold-based | Month |
| De Silva, 1986 [[58]](https://paperpile.com/c/zPIKrB/zsbw) | Australia | Sydney | 151·2, -33·9 | 1979 - 1983 | inpatient | ARI or ILI | Antigen detection | Qualitative | Month |
| Dearden, 2018 [[59]](https://paperpile.com/c/zPIKrB/kuOZ) | South Africa | Pretoria | 28·2, -25·7 | 2013 - 2016 | outpatient; inpatient | ALRI | Antigen detection; Nucleic acid detection | Qualitative | Month |
| Do, 2011 [[60]](https://paperpile.com/c/zPIKrB/tFOs) | Vietnam | Ho Chi Minh | 106·6, 10·8 | 2004 - 2008 | inpatient | SARI | Nucleic acid detection | Qualitative | Month |
| Duppenthaler, 2003 [[61]](https://paperpile.com/c/zPIKrB/CpdqX) | Switzerland | Bern | 7·4, 46·9 | 1997 - 2001 | inpatient | Clinical judgment | Antigen detection | Qualitative | Week |
| Duppenthaler, 2003 [[61]](https://paperpile.com/c/zPIKrB/CpdqX) | Switzerland | Bern | 7·4, 46·9 | 1997 - 2001 | inpatient | Clinical judgment | Antigen detection | Qualitative | Week |
| Duppenthaler, 2003 [[61]](https://paperpile.com/c/zPIKrB/CpdqX) | Switzerland | nationwide | 8·2, 46·8 | 1988 - 1999 | outpatient; inpatient | Unknown | Unknown | Threshold-based | Week |
| Duppenthaler, 2003 [[61]](https://paperpile.com/c/zPIKrB/CpdqX) | Switzerland | nationwide | 8·2, 46·8 | 1988 - 1999 | outpatient; inpatient | Unknown | Unknown | Threshold-based | Week |
| Eidelman, 2009 [[62]](https://paperpile.com/c/zPIKrB/d1TO) | Israel | Jerusalem | 35·2, 31·8 | 2002 - 2007 | inpatient | ALRI | Antigen detection | Qualitative | Month |
| Eriksson, 2002 [[63]](https://paperpile.com/c/zPIKrB/j8Al7) | Sweden | Stockholm | 18·1, 59·3 | 1987 - 1998 | inpatient | Clinical judgment | Antigen detection | Qualitative | Month |
| Feng, 2014 [[64]](https://paperpile.com/c/zPIKrB/PY7d) | China | nationwide | 104·2, 35·9 | 2009 - 2013 | inpatient | ALRI | Nucleic acid detection | Qualitative | Month |
| Fergie, 2007 [[65]](https://paperpile.com/c/zPIKrB/o3kmc) | USA | nationwide | -95·7, 37·1 | 1996 - 2007 | Unknown | Unknown | Antigen detection;Virus detection | Threshold-based | Week |
| Fergie, 2007 [[65]](https://paperpile.com/c/zPIKrB/o3kmc) | USA | South | -99, 32 | 1996 - 2007 | Unknown | Unknown | Antigen detection;Virus detection | Threshold-based | Week |
| Fergie, 2007 [[65]](https://paperpile.com/c/zPIKrB/o3kmc) | USA | South Texas state | -99·9, 32 | 1996 - 2007 | Unknown | Unknown | Antigen detection;Virus detection | Threshold-based | Week |
| Fergie, 2007 [[65]](https://paperpile.com/c/zPIKrB/o3kmc) | USA | South Texas state | -99·9, 32 | 1996 - 2007 | Unknown | Unknown | Antigen detection;Virus detection | Threshold-based | Week |
| Ferrero, 2016 [[66]](https://paperpile.com/c/zPIKrB/YkTtU) | Argentina | Buenos Aires | -58·4, -34·6 | 1995 - 2014 | Unknown | Unknown | Unknown | Threshold-based | Week |
| Ferrero, 2016 [[66]](https://paperpile.com/c/zPIKrB/YkTtU) | Argentina | nationwide | -63·6, -38·4 | 1995 - 2014 | Unknown | Unknown | Unknown | Threshold-based | Week |
| Fjaerli, 2004 [[67]](https://paperpile.com/c/zPIKrB/NZeh) | Norway | Akershus | 11·4, 60 | 1993 - 2000 | inpatient | ALRI | Antigen detection | Qualitative | Month |
| Fleming, 1993 [[68]](https://paperpile.com/c/zPIKrB/4ENS) | United Kingdom | nationwide | -1·6, 52·7 | 1989 - 1992 | Unknown | Unknown | Unknown | Qualitative | Week |
| Fleming, 2005 [[69]](https://paperpile.com/c/zPIKrB/kKmQz) | United Kingdom | England | -1·2, 52·4 | 1989 - 2000 | Unknown | Unknown | Unknown | Threshold-based | Week |
| Freitas, 2013 [[70]](https://paperpile.com/c/zPIKrB/tQMa) | Brazil | Midwest | -51·9, -14·2 | 2000 - 2010 | Unknown | ARI or ILI | Antigen detection | Qualitative | Month |
| Freitas, 2013 [[70]](https://paperpile.com/c/zPIKrB/tQMa) | Brazil | North | -58·4, -2·1 | 2000 - 2010 | Unknown | ARI or ILI | Antigen detection | Qualitative | Month |
| Freitas, 2013 [[70]](https://paperpile.com/c/zPIKrB/tQMa) | Brazil | Northeast | -42·6, -10·1 | 2000 - 2010 | Unknown | ARI or ILI | Antigen detection | Qualitative | Month |
| Freitas, 2013 [[70]](https://paperpile.com/c/zPIKrB/tQMa) | Brazil | South | -49·3, -25·4 | 2000 - 2010 | Unknown | ARI or ILI | Antigen detection | Qualitative | Month |
| Freitas, 2013 [[70]](https://paperpile.com/c/zPIKrB/tQMa) | Brazil | Southeast | -46·2, -20·3 | 2000 - 2010 | Unknown | ARI or ILI | Antigen detection | Qualitative | Month |
| Fry, 2006 [[71]](https://paperpile.com/c/zPIKrB/rMNIq) | USA | nationwide | -95·7, 37·1 | 1990 - 2004 | Unknown | Unknown | Antigen detection | Threshold-based | Week |
| García-Arroyo, 2022 [[72]](https://paperpile.com/c/zPIKrB/WKCsT) | Spain | Barcelona | 2·2, 41·4 | 1997 - 2020 | Unknown | Unknown | Virus detection;Antigen detection;Nucleic acid detection | Qualitative | Month |
| Gentile, 2019 [[73]](https://paperpile.com/c/zPIKrB/23GWJ) | Argentina | Buenos Aires | -58·4, -34·6 | 2000 - 2017 | inpatient | ALRI | Antigen detection; Nucleic acid detection | Threshold-based | Week |
| Gil-Prieto, 2015 [[74]](https://paperpile.com/c/zPIKrB/LQaL) | Spain | nationwide | -3·3, 39·7 | 1997 - 2011 | inpatient | ALRI | Unknown | Qualitative | Month |
| Glatman-Freedman, 2020 [[75]](https://paperpile.com/c/zPIKrB/nwDvF) | Israel | nationwide | 34·9, 31 | 2000 - 2017 | inpatient | Unknown | Unknown | Threshold-based | Month |
| Goddard, 2007 [[76]](https://paperpile.com/c/zPIKrB/XvW5a) | United Kingdom | nationwide | -1·6, 52·7 | 1994 - 2004 | Unknown | Unknown | Antigen detection; Nucleic acid detection | Threshold-based | Week |
| Grilc, 2021 [[77]](https://paperpile.com/c/zPIKrB/1iYBm) | Slovenia | nationwide | 14·9, 46·2 | 2008 - 2018 | outpatient; inpatient | Unknown | Nucleic acid detection | Coverage-based | Week |
| Grilc, 2021 [[77]](https://paperpile.com/c/zPIKrB/1iYBm) | Slovenia | nationwide | 14·9, 46·2 | 2008 - 2018 | outpatient; inpatient | Unknown | Nucleic acid detection | Threshold-based | Week |
| Gunell, 2016 [[78]](https://paperpile.com/c/zPIKrB/1N0Q) | Finland | nationwide | 25·7, 61·9 | 2010 - 2014 | Unknown | ARI or ILI | Antigen detection | Qualitative | Week |
| Halasa, 2015 [[79]](https://paperpile.com/c/zPIKrB/JjTJ) | Hashemite Kingdom of Jordan | Amman | 35·9, 32 | 2010 - 2013 | inpatient | ARI or ILI | Nucleic acid detection | Qualitative | Month |
| Halstead, 1998 [[80]](https://paperpile.com/c/zPIKrB/Raleq) | USA | Florida state | -81·5, 27·7 | 1993 - 1996 | Unknown | Unknown | Antigen detection | Threshold-based | Month |
| Hampp, 2013 [[81]](https://paperpile.com/c/zPIKrB/7vWjg) | USA | California state | -119·4, 36·8 | 1999 - 2004 | inpatient | Unknown | Unknown | Threshold-based | Week |
| Hampp, 2013 [[81]](https://paperpile.com/c/zPIKrB/7vWjg) | USA | California state | -119·4, 36·8 | 1999 - 2004 | inpatient | Unknown | Unknown | Threshold-based | Week |
| Hampp, 2013 [[81]](https://paperpile.com/c/zPIKrB/7vWjg) | USA | Florida state | -81·5, 27·7 | 1999 - 2004 | inpatient | Unknown | Unknown | Threshold-based | Week |
| Hampp, 2013 [[81]](https://paperpile.com/c/zPIKrB/7vWjg) | USA | Florida state | -81·5, 27·7 | 1999 - 2004 | inpatient | Unknown | Unknown | Threshold-based | Week |
| Hampp, 2013 [[81]](https://paperpile.com/c/zPIKrB/7vWjg) | USA | Illinois state | -89·4, 40·6 | 1999 - 2004 | inpatient | Unknown | Unknown | Threshold-based | Week |
| Hampp, 2013 [[81]](https://paperpile.com/c/zPIKrB/7vWjg) | USA | Illinois state | -89·4, 40·6 | 1999 - 2004 | inpatient | Unknown | Unknown | Threshold-based | Week |
| Hampp, 2013 [[81]](https://paperpile.com/c/zPIKrB/7vWjg) | USA | Texas state | -99·9, 32 | 1999 - 2004 | inpatient | Unknown | Unknown | Threshold-based | Week |
| Hampp, 2013 [[81]](https://paperpile.com/c/zPIKrB/7vWjg) | USA | Texas state | -99·9, 32 | 1999 - 2004 | inpatient | Unknown | Unknown | Threshold-based | Week |
| Haynes, 2013 [[82]](https://paperpile.com/c/zPIKrB/GMmUc) | Bangladesh | Dhaka, Bogra, Barisal, Comilla and Kishoreganj | 90·4, 23·7 | 2004 - 2011 | outpatient; inpatient | ARI or ILI | Nucleic acid detection | Threshold-based | Month |
| Haynes, 2013 [[82]](https://paperpile.com/c/zPIKrB/GMmUc) | Egypt | Damanhour | 30·5, 31 | 2009 - 2012 | outpatient; inpatient | ARI or ILI | Nucleic acid detection | Threshold-based | Month |
| Haynes, 2013 [[82]](https://paperpile.com/c/zPIKrB/GMmUc) | Guatemala | Santa Rosa, Guateala and Quetzaltenango | -90·4, 14·2 | 2007 - 2011 | outpatient; inpatient | ARI or ILI | Nucleic acid detection | Threshold-based | Month |
| Haynes, 2013 [[82]](https://paperpile.com/c/zPIKrB/GMmUc) | Kenya | Lwak and Kibera | 36·8, -1·3 | 2007 - 2011 | outpatient; inpatient | ARI or ILI | Nucleic acid detection | Threshold-based | Month |
| Haynes, 2013 [[82]](https://paperpile.com/c/zPIKrB/GMmUc) | South Africa | Pretoria and Soweto | 27·9, -26·2 | 2006 - 2012 | inpatient | ARI or ILI | Nucleic acid detection | Threshold-based | Month |
| Haynes, 2013 [[82]](https://paperpile.com/c/zPIKrB/GMmUc) | Thailand | Nakhon Phanom and Sae Kaeo | 104·8, 17·4 | 2005 - 2011 | outpatient; inpatient | ARI or ILI | Nucleic acid detection | Threshold-based | Month |
| Haynes, 2016 [[83]](https://paperpile.com/c/zPIKrB/HrPoU) | USA | nationwide | -95·7, 37·1 | 2008 - 2014 | Unknown | Unknown | Antigen detection | Threshold-based | Week |
| He, 2014 [[84]](https://paperpile.com/c/zPIKrB/m7VoW) | China | Shenzhen | 114·1, 22·5 | 2007 - 2010 | inpatient | SARI | Nucleic acid detection | Qualitative | Month |
| Hendaus, 2018 [[85]](https://paperpile.com/c/zPIKrB/Lvqn) | Qatar | nationwide | 51·2, 25·4 | 2010 - 2012 | inpatient | ALRI | Nucleic acid detection | Qualitative | Month |
| Hervás, 2012 [[86]](https://paperpile.com/c/zPIKrB/e4Bx) | Spain | Mallorca | 3, 39·7 | 1995 - 2006 | inpatient | ALRI | Virus detection; Antigen detection | Qualitative | Month |
| Hibino, 2018 [[87]](https://paperpile.com/c/zPIKrB/Ii5N) | Japan | Okinawa | 127·7, 26·1 | 2012 - 2015 | outpatient | ARI or ILI | Antigen detection | Qualitative | Month |
| Hirsh, 2014 [[88]](https://paperpile.com/c/zPIKrB/CQ96) | Israel | nationwide | 34·9, 31 | 2005 - 2012 | inpatient | SARI | Antigen detection; Nucleic acid detection | Qualitative | Week |
| Hogan, 2016 [[89]](https://paperpile.com/c/zPIKrB/VsIIl) | Australia | Goldfields, Western Australia | 121·5, -30·8 | 2000 - 2013 | Unknown | Unknown | Virus detection; Antibody detection; Antigen detection; Nucleic acid detection | Qualitative | Month |
| Hogan, 2016 [[89]](https://paperpile.com/c/zPIKrB/VsIIl) | Australia | Great Southern, Western Australia | 121·6, -27·7 | 2000 - 2013 | Unknown | Unknown | Virus detection; Antibody detection; Antigen detection; Nucleic acid detection | Qualitative | Month |
| Hogan, 2016 [[89]](https://paperpile.com/c/zPIKrB/VsIIl) | Australia | Kimberley, Western Australia | 125·9, -17·3 | 2000 - 2013 | Unknown | Unknown | Virus detection; Antibody detection; Antigen detection; Nucleic acid detection | Qualitative | Month |
| Hogan, 2016 [[89]](https://paperpile.com/c/zPIKrB/VsIIl) | Australia | Metropoltan, Western Australia | 121·6, -27·7 | 2000 - 2013 | Unknown | Unknown | Virus detection; Antibody detection; Antigen detection; Nucleic acid detection | Qualitative | Month |
| Hogan, 2016 [[89]](https://paperpile.com/c/zPIKrB/VsIIl) | Australia | Midwest-Murchison, Western Australia | 116, -26·5 | 2000 - 2013 | Unknown | Unknown | Virus detection; Antibody detection; Antigen detection; Nucleic acid detection | Qualitative | Month |
| Hogan, 2016 [[89]](https://paperpile.com/c/zPIKrB/VsIIl) | Australia | Pilbara, Western Australia | 121·5, -21·6 | 2000 - 2013 | Unknown | Unknown | Virus detection; Antibody detection; Antigen detection; Nucleic acid detection | Qualitative | Month |
| Hogan, 2016 [[89]](https://paperpile.com/c/zPIKrB/VsIIl) | Australia | South West, Western Australia | 118, -32 | 2000 - 2013 | Unknown | Unknown | Virus detection; Antibody detection; Antigen detection; Nucleic acid detection | Qualitative | Month |
| Hogan, 2016 [[89]](https://paperpile.com/c/zPIKrB/VsIIl) | Australia | Wheatbelt, Western Australia | 118·1, -32 | 2000 - 2013 | Unknown | Unknown | Virus detection; Antibody detection; Antigen detection; Nucleic acid detection | Qualitative | Month |
| [[90]](https://paperpile.com/c/zPIKrB/ktSc), 2019 [[90]](https://paperpile.com/c/zPIKrB/ktSc) | South Africa | nationwide | 22·9, -30·6 | 2006 - 2008 | inpatient | Unknown | Virus detection; Antigen detection | Qualitative | Month |
| Horton, 2017 [[91]](https://paperpile.com/c/zPIKrB/T8rb) | Egypt, Jordan, Oman, Qatar and Yemen | nationwide | 44·8, 24·3 | 2007 - 2014 | inpatient | SARI | Nucleic acid detection | Qualitative | Month |
| Houspie, 2013 [[92]](https://paperpile.com/c/zPIKrB/hReV) | Belgium | nationwide | 4·5, 50·5 | 2006 - 2010 | Unknown | ARI or ILI | Antigen detection | Qualitative | Month |
| Hsu, 2014 [[93]](https://paperpile.com/c/zPIKrB/I3rwW) | Taiwan | greater Taipei metropolitan area | 121·5, 25·1 | 2000 - 2010 | inpatient | ALRI | Virus detection; Antigen detection | Threshold-based | Month |
| Hu, 2017 [[94]](https://paperpile.com/c/zPIKrB/8SKn) | China | Chengdu | 104·1, 30·6 | 2009 - 2014 | inpatient | ARI or ILI | Nucleic acid detection | Qualitative | Month |
| Huang, 2001 [[95]](https://paperpile.com/c/zPIKrB/3Gde) | Taiwan | Northern Taiwan | 122, 25 | 1995 - 1999 | inpatient | ALRI | Antigen detection | Qualitative | Month |
| Huang, 2020 [[96]](https://paperpile.com/c/zPIKrB/5Q3h) | China | Guangzhou | 113·3, 23·1 | 2009 - 2018 | outpatient; inpatient | ARI or ILI | Nucleic acid detection | Qualitative | Month |
| Irmen, 2000 [[97]](https://paperpile.com/c/zPIKrB/xcSo5) | USA | Bismarck | -101, 46·8 | 1987 - 1998 | outpatient; inpatient | Unknown | Virus detection; Antigen detection | Qualitative | Month |
| Jepsen, 2018 [[98]](https://paperpile.com/c/zPIKrB/EaElK) | Denmark | nationwide | 10·9, 55·9 | 2010 - 2015 | inpatient | Unknown | Unknown | Qualitative | Month |
| Jiang, 2023 [[99]](https://paperpile.com/c/zPIKrB/UJyei) | China | Beijing | 116·4, 39·9 | 4291 - 2021 | outpatient; inpatient | ARI or ILI | Nucleic acid detection | Threshold-based | Month |
| Jin, 2012 [[100]](https://paperpile.com/c/zPIKrB/9VLa) | China | Lanzhou | 103·8, 36·1 | 2006 - 2009 | inpatient | ALRI | Nucleic acid detection | Qualitative | Month |
| Kaneko, 2002 [[101]](https://paperpile.com/c/zPIKrB/KPEg) | Japan | Shizuoka | 138·4, 35 | 1997 - 2000 | inpatient | ALRI | Antibody detection; Antigen detection | Qualitative | Month |
| Karron, 1999 [[102]](https://paperpile.com/c/zPIKrB/YOac) | USA | YK Delta | -164·2, 62·9 | 1993 - 1996 | inpatient | ARI or ILI | Virus detection; Antigen detection | Qualitative | Month |
| Khor, 2012 [[103]](https://paperpile.com/c/zPIKrB/IIXB) | Malaysia | Kuala Lumpur | 101·7, 3·1 | 1982 - 2008 | inpatient | ARI or ILI | Virus detection; Antigen detection | Qualitative | Month |
| Korsun, 2019 [[104]](https://paperpile.com/c/zPIKrB/zVld) | Bulgaria | nationwide | 25·5, 42·7 | 2015 - 2018 | outpatient; inpatient | ALRI | Nucleic acid detection | Qualitative | Month |
| Kyeyagalire, 2014 [[105]](https://paperpile.com/c/zPIKrB/H2UB) | South Africa | nationwide | 22·9, -30·6 | 2007 - 2012 | Unknown | Unknown | Unknown | Qualitative | Month |
| Lagacé-Wiens, 2021 [[106]](https://paperpile.com/c/zPIKrB/vDVJf) | Canada | nationwide | -106·3, 56·1 | 2010 - 2020 | outpatient; inpatient | ARI or ILI | Virus detection; Antigen detection; Nucleic acid detection | Threshold-based | Week |
| Lam, 2019 [[107]](https://paperpile.com/c/zPIKrB/WnPPa) | Australia | Brisbane | 153, -27·5 | 2010 - 2015 | outpatient; inpatient | Unknown | Nucleic acid detection | Model-based | Month |
| Lam, 2019 [[107]](https://paperpile.com/c/zPIKrB/WnPPa) | Australia | Sydney | 151·2, -33·9 | 2010 - 2015 | outpatient; inpatient | Unknown | Nucleic acid detection | Model-based | Month |
| Lam, 2019 [[107]](https://paperpile.com/c/zPIKrB/WnPPa) | Canada | Edmonton | -113, 53·5 | 2010 - 2015 | inpatient | Unknown | Nucleic acid detection | Model-based | Month |
| Lam, 2019 [[107]](https://paperpile.com/c/zPIKrB/WnPPa) | Canada | Halifax | -63·1, 44·9 | 2010 - 2015 | outpatient; inpatient | Unknown | Nucleic acid detection | Model-based | Month |
| Lam, 2019 [[107]](https://paperpile.com/c/zPIKrB/WnPPa) | Canada | Vancouver | -123, 49·3 | 2010 - 2015 | outpatient; inpatient | Unknown | Nucleic acid detection | Model-based | Month |
| Lam, 2019 [[107]](https://paperpile.com/c/zPIKrB/WnPPa) | Finland | Turku | 22·3, 60·5 | 2010 - 2015 | outpatient; inpatient | Unknown | Nucleic acid detection | Model-based | Month |
| Lam, 2019 [[107]](https://paperpile.com/c/zPIKrB/WnPPa) | Hong Kong | nationwide | 114·2, 22·3 | 2010 - 2015 | outpatient; inpatient | Unknown | Antigen detection | Model-based | Month |
| Lam, 2019 [[107]](https://paperpile.com/c/zPIKrB/WnPPa) | Japan | Sendai | 141, 38·3 | 2010 - 2015 | outpatient | Unknown | Virus detection; Antigen detection | Model-based | Month |
| Lam, 2019 [[107]](https://paperpile.com/c/zPIKrB/WnPPa) | Mongolia | Ulaanbaatar | 107, 47·9 | 2010 - 2015 | outpatient; inpatient | Unknown | Nucleic acid detection | Model-based | Month |
| Lam, 2019 [[107]](https://paperpile.com/c/zPIKrB/WnPPa) | New Zealand | Canterbury | 171, -43·8 | 2010 - 2015 | outpatient; inpatient | Unknown | Nucleic acid detection | Model-based | Month |
| Lam, 2019 [[107]](https://paperpile.com/c/zPIKrB/WnPPa) | The Netherlands | Rotterdam | 4·5, 51·9 | 2010 - 2015 | outpatient; inpatient | Unknown | Nucleic acid detection | Model-based | Month |
| Lam, 2019 [[107]](https://paperpile.com/c/zPIKrB/WnPPa) | United Kingdom | Cambridge | 0·1, 52·2 | 2010 - 2015 | inpatient | Unknown | Nucleic acid detection | Model-based | Month |
| Lam, 2019 [[107]](https://paperpile.com/c/zPIKrB/WnPPa) | United Kingdom | Leicester | -1·1, 52·6 | 2010 - 2015 | inpatient | Unknown | Nucleic acid detection | Model-based | Month |
| Leecaster, 2011 [[108]](https://paperpile.com/c/zPIKrB/nwSk) | USA | Salt Lake County | -112, 40·6 | 2001 - 2008 | outpatient; inpatient | ARI or ILI | Antigen detection; Nucleic acid detection | Qualitative | Month |
| Li, 2019 [[3]](https://paperpile.com/c/zPIKrB/BQ74y) | Argentina | Buenos Aires | -58·4, -34·6 | 1998 - 2002 | inpatient | ALRI | Antigen detection | Coverage-based | Month |
| Li, 2019 [[3]](https://paperpile.com/c/zPIKrB/BQ74y) | Argentina | nationwide | -63·6, -38·4 | 2010 - 2017 | Unknown | Unknown | Unknown | Coverage-based | Month |
| Li, 2019 [[3]](https://paperpile.com/c/zPIKrB/BQ74y) | Aruba | nationwide | -70, 12·5 | 2010 - 2017 | Unknown | Unknown | Unknown | Coverage-based | Month |
| Li, 2019 [[3]](https://paperpile.com/c/zPIKrB/BQ74y) | Austria | Vienna | 16·4, 48·2 | 2000 - 2007 | inpatient | ARI or ILI | Nucleic acid detection | Coverage-based | Month |
| Li, 2019 [[3]](https://paperpile.com/c/zPIKrB/BQ74y) | Bardados | nationwide | -59·5, 13·2 | 2010 - 2017 | Unknown | Unknown | Unknown | Coverage-based | Month |
| Li, 2019 [[3]](https://paperpile.com/c/zPIKrB/BQ74y) | Belgium | Leuven | 4·7, 50·9 | 2011 - 2016 | inpatient | ARI or ILI | Nucleic acid detection | Coverage-based | Month |
| Li, 2019 [[3]](https://paperpile.com/c/zPIKrB/BQ74y) | Bolivia | nationwide | -63·6, -16·3 | 2010 - 2017 | Unknown | Unknown | Unknown | Coverage-based | Month |
| Li, 2019 [[3]](https://paperpile.com/c/zPIKrB/BQ74y) | Brazil | Fortaleza | -38·5, -3·7 | 2006 - 2008 | Unknown | ARI or ILI | Antigen detection | Coverage-based | Month |
| Li, 2019 [[3]](https://paperpile.com/c/zPIKrB/BQ74y) | Brazil | nationwide | -51·9, -14·2 | 2010 - 2017 | Unknown | Unknown | Unknown | Coverage-based | Month |
| Li, 2019 [[3]](https://paperpile.com/c/zPIKrB/BQ74y) | Brazil | Salvador | -38·5, -13 | 4005 - 4154 | outpatient; inpatient | ARI or ILI | Nucleic acid detection | Coverage-based | Month |
| Li, 2019 [[3]](https://paperpile.com/c/zPIKrB/BQ74y) | Canada | nationwide | -106·3, 56·1 | 2008 - 2014 | Unknown | Unknown | Unknown | Coverage-based | Month |
| Li, 2019 [[3]](https://paperpile.com/c/zPIKrB/BQ74y) | Canada | Nova Scotia | -63·7, 44·7 | 2005 - 2008 | outpatient; inpatient | ARI or ILI | Virus detection; Antigen detection; Nucleic acid detection | Coverage-based | Month |
| Li, 2019 [[3]](https://paperpile.com/c/zPIKrB/BQ74y) | Chile | nationwide | -71·5, -35·7 | 2010 - 2017 | Unknown | Unknown | Unknown | Coverage-based | Month |
| Li, 2019 [[3]](https://paperpile.com/c/zPIKrB/BQ74y) | Colombia | nationwide | -74·3, 4·6 | 2010 - 2017 | Unknown | Unknown | Unknown | Coverage-based | Month |
| Li, 2019 [[3]](https://paperpile.com/c/zPIKrB/BQ74y) | Croatia | Zagreb County | 16·4, 45·9 | 1994 - 2005 | inpatient | ARI or ILI | Virus detection; Antigen detection | Coverage-based | Month |
| Li, 2019 [[3]](https://paperpile.com/c/zPIKrB/BQ74y) | Cuba | nationwide | -77·8, 21·5 | 2010 - 2017 | Unknown | Unknown | Unknown | Coverage-based | Month |
| Li, 2019 [[3]](https://paperpile.com/c/zPIKrB/BQ74y) | Dominica | nationwide | -61·4, 15·4 | 2010 - 2017 | Unknown | Unknown | Unknown | Coverage-based | Month |
| Li, 2019 [[3]](https://paperpile.com/c/zPIKrB/BQ74y) | Dominican Republic | nationwide | -70·2, 18·7 | 2010 - 2017 | Unknown | Unknown | Unknown | Coverage-based | Month |
| Li, 2019 [[3]](https://paperpile.com/c/zPIKrB/BQ74y) | Ecuador | nationwide | -78·2, -1·8 | 2010 - 2017 | Unknown | Unknown | Unknown | Coverage-based | Month |
| Li, 2019 [[3]](https://paperpile.com/c/zPIKrB/BQ74y) | Egypt | nationwide | 30·8, 26·8 | 2007 - 2014 | inpatient | SARI | Nucleic acid detection | Coverage-based | Month |
| Li, 2019 [[3]](https://paperpile.com/c/zPIKrB/BQ74y) | EI Salvador | nationwide | -88·9, 13·8 | 2010 - 2017 | Unknown | Unknown | Unknown | Coverage-based | Month |
| Li, 2019 [[3]](https://paperpile.com/c/zPIKrB/BQ74y) | Gambia | Banjul | -16·6, 13·5 | 1993 - 2002 | inpatient | SARI | Antigen detection | Coverage-based | Month |
| Li, 2019 [[3]](https://paperpile.com/c/zPIKrB/BQ74y) | Germany | Freiburg | 7·8, 48 | 1988 - 1999 | inpatient | ARI or ILI | Antigen detection | Coverage-based | Month |
| Li, 2019 [[3]](https://paperpile.com/c/zPIKrB/BQ74y) | Germany | Kiel | 10·1, 54·3 | 1994 - 2001 | inpatient | SARI | Antigen detection; Nucleic acid detection | Coverage-based | Month |
| Li, 2019 [[3]](https://paperpile.com/c/zPIKrB/BQ74y) | Germany | Stuttgart | 9·2, 48·8 | 1996 - 2004 | outpatient; inpatient | Clinical judgment | Antigen detection; Nucleic acid detection | Coverage-based | Month |
| Li, 2019 [[3]](https://paperpile.com/c/zPIKrB/BQ74y) | Greece | Athens | 23·7, 37·7 | 2002 - 2013 | inpatient | SARI | Antigen detection | Coverage-based | Month |
| Li, 2019 [[3]](https://paperpile.com/c/zPIKrB/BQ74y) | Guatemala | nationwide | -90·2, 15·8 | 2010 - 2017 | Unknown | Unknown | Unknown | Coverage-based | Month |
| Li, 2019 [[3]](https://paperpile.com/c/zPIKrB/BQ74y) | Hashemite Kingdom of Jordan | nationwide | 36·2, 30·6 | 2007 - 2014 | inpatient | SARI | Nucleic acid detection | Coverage-based | Month |
| Li, 2019 [[3]](https://paperpile.com/c/zPIKrB/BQ74y) | Honduras | nationwide | -86·2, 15·2 | 2010 - 2017 | Unknown | Unknown | Unknown | Coverage-based | Month |
| Li, 2019 [[3]](https://paperpile.com/c/zPIKrB/BQ74y) | Latvia | Riga | 24·1, 56·9 | 2009 - 2012 | inpatient | ALRI | Nucleic acid detection | Coverage-based | Month |
| Li, 2019 [[3]](https://paperpile.com/c/zPIKrB/BQ74y) | Oman | nationwide | 56, 21·5 | 2007 - 2014 | inpatient | SARI | Nucleic acid detection | Coverage-based | Month |
| Li, 2019 [[3]](https://paperpile.com/c/zPIKrB/BQ74y) | Pakistan | Karachi | 67, 24·9 | 2009 - 2012 | inpatient | ARI or ILI | Nucleic acid detection | Coverage-based | Month |
| Li, 2019 [[3]](https://paperpile.com/c/zPIKrB/BQ74y) | Qatar | Doha | 51·5, 25·3 | 1996 - 1998 | inpatient | Unknown | Antigen detection | Coverage-based | Month |
| Li, 2019 [[3]](https://paperpile.com/c/zPIKrB/BQ74y) | Spain | Leganés | -3·8, 40·3 | 2005 - 2013 | inpatient | ALRI | Nucleic acid detection | Coverage-based | Month |
| Li, 2019 [[3]](https://paperpile.com/c/zPIKrB/BQ74y) | Spain | Mallorca | 3, 39·7 | 1995 - 2006 | inpatient | ALRI | Virus detection; Antigen detection | Coverage-based | Month |
| Li, 2019 [[3]](https://paperpile.com/c/zPIKrB/BQ74y) | Spain | Sacyl | 41·7, -4·7 | 1992 - 2004 | inpatient | ALRI | Antigen detection | Coverage-based | Month |
| Li, 2019 [[3]](https://paperpile.com/c/zPIKrB/BQ74y) | Sweden | Gothenburg | 12, 57·7 | 2010 - 2013 | outpatient; inpatient | ARI or ILI | Nucleic acid detection | Coverage-based | Month |
| Li, 2019 [[3]](https://paperpile.com/c/zPIKrB/BQ74y) | Switzerland | Basel | 7·6, 47·6 | 2004 - 2008 | outpatient; inpatient | ARI or ILI | Nucleic acid detection | Coverage-based | Month |
| Li, 2019 [[3]](https://paperpile.com/c/zPIKrB/BQ74y) | Switzerland | Bern | 7·4, 46·9 | 1998 - 2010 | inpatient | ARI or ILI | Antigen detection | Coverage-based | Month |
| Li, 2019 [[3]](https://paperpile.com/c/zPIKrB/BQ74y) | United Kingdom | England | -1·2, 52·4 | 2007 - 2012 | inpatient | Unknown | Unknown | Coverage-based | Month |
| Li, 2022 [[109]](https://paperpile.com/c/zPIKrB/68Em7) | Denmark | nationwide | 10·9, 55·9 | 2012 - 2019 | Unknown | ARI or ILI; Clinical judgment | Unknown | Coverage-based | Week |
| Li, 2022 [[109]](https://paperpile.com/c/zPIKrB/68Em7) | Estonia | nationwide | 25·5, 59 | 2012 - 2018 | Unknown | ARI or ILI | Unknown | Coverage-based | Week |
| Li, 2022 [[109]](https://paperpile.com/c/zPIKrB/68Em7) | Germany | nationwide | 9·7, 50·9 | 2013 - 2019 | Unknown | ARI or ILI | Unknown | Coverage-based | Week |
| Li, 2022 [[109]](https://paperpile.com/c/zPIKrB/68Em7) | Germany | nationwide | 9·7, 50·9 | 2013 - 2019 | Unknown | ARI or ILI | Unknown | Coverage-based | Week |
| Li, 2022 [[109]](https://paperpile.com/c/zPIKrB/68Em7) | Ireland | nationwide | -7·4, 53·1 | 2012 - 2019 | Unknown | Clinical judgment | Unknown | Coverage-based | Week |
| Li, 2022 [[109]](https://paperpile.com/c/zPIKrB/68Em7) | Ireland | nationwide | -7·4, 53·1 | 2014 - 2019 | Unknown | ARI or ILI | Unknown | Coverage-based | Week |
| Li, 2022 [[109]](https://paperpile.com/c/zPIKrB/68Em7) | Poland | nationwide | 19·3, 51·7 | 2012 - 2019 | Unknown | ARI or ILI | Unknown | Coverage-based | Week |
| Li, 2022 [[109]](https://paperpile.com/c/zPIKrB/68Em7) | Portugal | nationwide | -9·2, 39·7 | 2012 - 2017 | Unknown | ARI or ILI; Clinical judgment | Unknown | Coverage-based | Week |
| Li, 2022 [[109]](https://paperpile.com/c/zPIKrB/68Em7) | Slovenia | nationwide | 14·9, 46·2 | 2010 - 2019 | Unknown | Clinical judgment | Unknown | Coverage-based | Week |
| Li, 2022 [[109]](https://paperpile.com/c/zPIKrB/68Em7) | Slovenia | nationwide | 14·9, 46·2 | 2012 - 2019 | Unknown | ARI or ILI | Unknown | Coverage-based | Week |
| Li, 2022 [[109]](https://paperpile.com/c/zPIKrB/68Em7) | Spain | nationwide | -3·3, 39·7 | 2012 - 2019 | Unknown | Clinical judgment | Unknown | Coverage-based | Week |
| Li, 2022 [[109]](https://paperpile.com/c/zPIKrB/68Em7) | The Netherlands | nationwide | 5·3, 52·1 | 2010 - 2019 | Unknown | Clinical judgment | Unknown | Coverage-based | Week |
| Li, 2022 [[109]](https://paperpile.com/c/zPIKrB/68Em7) | The Netherlands | nationwide | 5·3, 52·1 | 2012 - 2019 | Unknown | ARI or ILI | Unknown | Coverage-based | Week |
| Li, 2022 [[109]](https://paperpile.com/c/zPIKrB/68Em7) | United Kingdom | nationwide | -1·6, 52·7 | 2012 - 2019 | Unknown | ARI or ILI | Unknown | Coverage-based | Week |
| Li, 2022 [[109]](https://paperpile.com/c/zPIKrB/68Em7) | United Kingdom | nationwide | -1·6, 52·7 | 2012 - 2019 | Unknown | ARI or ILI | Unknown | Coverage-based | Week |
| Light, 2007 [[110]](https://paperpile.com/c/zPIKrB/VCs9C) | USA | Southeast, Florida state | -80·4, 26·2 | 2003 - 2006 | outpatient | ALRI | Unknown | Threshold-based | Month |
| Light, 2008 [[111]](https://paperpile.com/c/zPIKrB/XkHEI) | USA | Central, Florida state | -81·9, 28·1 | 2001 - 2004 | Unknown | Unknown | Virus detection; Nucleic acid detection | Threshold-based | Month |
| Light, 2008 [[111]](https://paperpile.com/c/zPIKrB/XkHEI) | USA | North, Florida state | -80·2, 26·2 | 2001 - 2004 | Unknown | Unknown | Virus detection; Nucleic acid detection | Threshold-based | Month |
| Light, 2008 [[111]](https://paperpile.com/c/zPIKrB/XkHEI) | USA | Northwest, Florida state | -86·5, 30·5 | 2001 - 2004 | Unknown | Unknown | Virus detection; Nucleic acid detection | Threshold-based | Month |
| Light, 2008 [[111]](https://paperpile.com/c/zPIKrB/XkHEI) | USA | Southeast, Florida state | -80·4, 26·2 | 2001 - 2004 | Unknown | Unknown | Virus detection; Nucleic acid detection | Threshold-based | Month |
| Light, 2008 [[111]](https://paperpile.com/c/zPIKrB/XkHEI) | USA | Southwest, Florida state | -81·9, 26·6 | 2001 - 2004 | Unknown | Unknown | Virus detection; Nucleic acid detection | Threshold-based | Month |
| Liu, 2014 [[112]](https://paperpile.com/c/zPIKrB/CEqg) | China | nationwide | 104·2, 35·9 | 2009 - 2012 | outpatient | ARI or ILI | Nucleic acid detection | Qualitative | Month |
| Liu, 2014 [[112]](https://paperpile.com/c/zPIKrB/CEqg) | China | Shanghai | 121·5, 31·2 | 2009 - 2012 | outpatient | ARI or ILI | Nucleic acid detection | Qualitative | Month |
| Liu, 2018 [[113]](https://paperpile.com/c/zPIKrB/Sq1i) | China | Shanghai | 121·5, 31·2 | 2013 - 2015 | inpatient | ALRI | Antigen detection | Qualitative | Month |
| Liu, 2019 [[114]](https://paperpile.com/c/zPIKrB/rm9oD) | China | Guangzhou | 113·3, 23·1 | 2009 - 2016 | inpatient | ARI or ILI | Nucleic acid detection | Qualitative | Month |
| Loconsole, 2022 [[115]](https://paperpile.com/c/zPIKrB/1S5Pg) | Italy | Bari | 16·9, 41·4 | 2017 - 2020 | inpatient | Unknown | Nucleic acid detection | Qualitative | Month |
| Loh, 2011 [[116]](https://paperpile.com/c/zPIKrB/yLS8) | Singapore | nationwide | 103·8, 1·4 | 2003 - 2008 | outpatient; inpatient | ARI or ILI | Antigen detection | Qualitative | Month |
| Low, 2022 [[117]](https://paperpile.com/c/zPIKrB/8OXnW) | Malaysia | nationwide | 102, 4·2 | 2015 - 2019 | Unknown | ARI or ILI | Nucleic acid detection | Qualitative | Month |
| Lu, 2015 [[118]](https://paperpile.com/c/zPIKrB/UFDf) | China | Suzhou | 120·6, 31·3 | 2010 - 2014 | inpatient | ALRI | Antigen detection; Nucleic acid detection | Qualitative | Month |
| Lumley, 2022 [[119]](https://paperpile.com/c/zPIKrB/L8WR) | United Kingdom | Oxford and Banbury | -1·3, 52·1 | 2016 - 2019 | outpatient; inpatient | ARI or ILI; Clinical judgment | Nucleic acid detection | Qualitative | Month |
| Luo, 2022 [[120]](https://paperpile.com/c/zPIKrB/RCZsO) | China | Beijing | 116·4, 39·9 | 2015 - 2019 | outpatient; inpatient | ALRI;ARI or ILI | Nucleic acid detection | Qualitative | Month |
| Mak, 2012 [[121]](https://paperpile.com/c/zPIKrB/J6R1R) | Hong Kong | nationwide | 114·2, 22·3 | 2004 - 2011 | Unknown | Unknown | Virus detection | Qualitative | Month |
| Martin, 1978 [[122]](https://paperpile.com/c/zPIKrB/Fz7X) | United Kingdom | Newcastle | -1·6, 55 | 1971 - 1977 | inpatient | SARI | Virus detection; Antigen detection | Qualitative | Month |
| McCracken, 2014 [[123]](https://paperpile.com/c/zPIKrB/RKhU) | Guatemala | nationwide | -90·2, 15·8 | 2007 - 2012 | inpatient | ARI or ILI | Nucleic acid detection | Qualitative | Month |
| McGuiness, 2014 [[124]](https://paperpile.com/c/zPIKrB/aBsPn) | USA | Florida state | -81·5, 27·7 | 2007 - 2012 | Unknown | Unknown | Virus detection; Antigen detection; Nucleic acid detection | Threshold-based | Week |
| McGuiness, 2014 [[124]](https://paperpile.com/c/zPIKrB/aBsPn) | USA | Midwest | -93·1, 41·9 | 2007 - 2012 | Unknown | Unknown | Virus detection; Antigen detection; Nucleic acid detection | Threshold-based | Week |
| McGuiness, 2014 [[124]](https://paperpile.com/c/zPIKrB/aBsPn) | USA | nationwide | -95·7, 37·1 | 2007 - 2012 | Unknown | Unknown | Virus detection; Antigen detection; Nucleic acid detection | Threshold-based | Week |
| McGuiness, 2014 [[124]](https://paperpile.com/c/zPIKrB/aBsPn) | USA | nationwide | -95·7, 37·1 | 2007 - 2012 | Unknown | Unknown | Virus detection; Antigen detection; Nucleic acid detection | Threshold-based | Week |
| McGuiness, 2014 [[124]](https://paperpile.com/c/zPIKrB/aBsPn) | USA | Northeast | -74·2, 43·3 | 2007 - 2012 | Unknown | Unknown | Virus detection; Antigen detection; Nucleic acid detection | Threshold-based | Week |
| McGuiness, 2014 [[124]](https://paperpile.com/c/zPIKrB/aBsPn) | USA | South | -99, 32 | 2007 - 2012 | Unknown | Unknown | Virus detection; Antigen detection; Nucleic acid detection | Threshold-based | Week |
| McGuiness, 2014 [[124]](https://paperpile.com/c/zPIKrB/aBsPn) | USA | West | -111, 37·1 | 2007 - 2012 | Unknown | Unknown | Virus detection; Antigen detection; Nucleic acid detection | Threshold-based | Week |
| Meerhoff, 2009 [[125]](https://paperpile.com/c/zPIKrB/br0sv) | The Netherlands | nationwide | 5·3, 52·1 | 1998 - 2005 | inpatient | ALRI | Virus detection; Antigen detection; Nucleic acid detection | Threshold-based | Week |
| Meerhoff, 2009 [[125]](https://paperpile.com/c/zPIKrB/br0sv) | The Netherlands | nationwide | 5·3, 52·1 | 1998 - 2005 | inpatient | ALRI | Virus detection; Antigen detection; Nucleic acid detection | Threshold-based | Week |
| Meningher, 2014 [[126]](https://paperpile.com/c/zPIKrB/LCRZ) | Israel | nationwide | 34·9, 31 | 2007 - 2012 | Unknown | ARI or ILI | Nucleic acid detection | Qualitative | Week |
| Midgley, 2017 [[127]](https://paperpile.com/c/zPIKrB/LX59k) | USA | nationwide | -95·7, 37·1 | 2005 - 2015 | Unknown | Unknown | Virus detection; Antigen detection; Nucleic acid detection | Threshold-based | Week |
| Miller, 2013 [[128]](https://paperpile.com/c/zPIKrB/7hOJ) | USA | Tennessee | -86·6, 35·5 | 2004 - 2008 | outpatient; inpatient | ARI or ILI | Nucleic acid detection | Qualitative | Month |
| Miyama, 2021 [[129]](https://paperpile.com/c/zPIKrB/hD5ZO) | Japan | nationwide | 138·3, 36·2 | 2012 - 2019 | Unknown | Unknown | Antigen detection; Nucleic acid detection | Model-based | Week |
| Mizuta, 2013 [[130]](https://paperpile.com/c/zPIKrB/UQ2i) | Japan | Yamagata prefecture | 140·1, 38·5 | 2004 - 2011 | outpatient | ARI or ILI | Antibody detection; Antigen detection;Virus detection; Nucleic acid detection | Qualitative | Month |
| Mlinaric-Galinovic, 2008[[131]](https://paperpile.com/c/zPIKrB/otzRz) | Croatia | Zagreb County | 16·4, 45·9 | 1994 - 2005 | inpatient | ARI or ILI | Virus detection; Antigen detection | Qualitative | Month |
| Montgomery, 2021 [[132]](https://paperpile.com/c/zPIKrB/3KyN) | USA | Oahu | -158, 21·4 | 2014 - 2018 | outpatient; inpatient | Unknown | Virus detection; Antigen detection; Nucleic acid detection | Qualitative | Month |
| Moore, 2009 [[133]](https://paperpile.com/c/zPIKrB/mAtYr) | Australia | Perth | 115·9, -32 | 1997 - 2005 | outpatient; inpatient | Unknown | Virus detection; Antigen detection | Qualitative | Month |
| Morley, 2018 [[134]](https://paperpile.com/c/zPIKrB/5eVn) | Australia | Gold Coast region of South East Queensland | 153·4, -28 | 2007 - 2016 | outpatient; inpatient | ARI or ILI | Antigen detection; Nucleic acid detection | Qualitative | Month |
| Moura, 2013 [[135]](https://paperpile.com/c/zPIKrB/yqMda) | Brazil | Fortaleza | -38·5, -3·7 | 2004 - 2008 | Unknown | ARI or ILI | Antigen detection | Threshold-based | Week |
| Movva, 2022 [[136]](https://paperpile.com/c/zPIKrB/ddPlR) | USA | nationwide | -95·7, 37·1 | 2015 - 2020 | outpatient; inpatient | Unknown | Unknown | Qualitative | NA |
| Mufson, 1973 [[137]](https://paperpile.com/c/zPIKrB/ACqt) | USA | Chicago | -87·6, 41·9 | 1967 - 1971 | inpatient | ALRI | Virus detection; Antibody detection | Qualitative | Month |
| Mullins, 2003 [[138]](https://paperpile.com/c/zPIKrB/QzfFW) | USA | Midwest | -93·1, 41·9 | 1990 - 2000 | Unknown | Unknown | Antigen detection | Threshold-based | Week |
| Mullins, 2003 [[138]](https://paperpile.com/c/zPIKrB/QzfFW) | USA | nationwide | -95·7, 37·1 | 1990 - 2000 | Unknown | Unknown | Antigen detection | Threshold-based | Week |
| Mullins, 2003 [[138]](https://paperpile.com/c/zPIKrB/QzfFW) | USA | Northeast | -74·2, 43·3 | 1990 - 2000 | Unknown | Unknown | Antigen detection | Threshold-based | Week |
| Mullins, 2003 [[138]](https://paperpile.com/c/zPIKrB/QzfFW) | USA | South | -99, 32 | 1990 - 2000 | Unknown | Unknown | Antigen detection | Threshold-based | Week |
| Mullins, 2003 [[138]](https://paperpile.com/c/zPIKrB/QzfFW) | USA | West | -111, 37·1 | 1990 - 2000 | Unknown | Unknown | Antigen detection | Threshold-based | Week |
| Nenna, 2017 [[4]](https://paperpile.com/c/zPIKrB/JH4bW) | Italy | Rome | 12·5, 41·9 | 2004 - 2014 | inpatient | ALRI | Nucleic acid detection | Model-based | Month |
| Noveroske, 2016 [[139]](https://paperpile.com/c/zPIKrB/JiaW8) | USA | Fairfield, Connecticut state | -73·3, 41·1 | 1997 - 2013 | inpatient | Unknown | Unknown | Model-based | Week |
| Noveroske, 2016 [[139]](https://paperpile.com/c/zPIKrB/JiaW8) | USA | Hartford, Connecticut state | -72·7, 41·8 | 1997 - 2013 | inpatient | Unknown | Unknown | Model-based | Week |
| Noveroske, 2016 [[139]](https://paperpile.com/c/zPIKrB/JiaW8) | USA | Litchfield, Connecticut state | -73·2, 41·7 | 1997 - 2013 | inpatient | Unknown | Unknown | Model-based | Week |
| Noveroske, 2016 [[139]](https://paperpile.com/c/zPIKrB/JiaW8) | USA | Middlesex, Connecticut state | -72·5, 41·5 | 1997 - 2013 | inpatient | Unknown | Unknown | Model-based | Week |
| Noveroske, 2016 [[139]](https://paperpile.com/c/zPIKrB/JiaW8) | USA | New Haven, Connecticut state | -72·9, 41·3 | 1997 - 2013 | inpatient | Unknown | Unknown | Model-based | Week |
| Noveroske, 2016 [[139]](https://paperpile.com/c/zPIKrB/JiaW8) | USA | New London, Connecticut state | -72·1, 41·4 | 1997 - 2013 | inpatient | Unknown | Unknown | Model-based | Week |
| Noveroske, 2016 [[139]](https://paperpile.com/c/zPIKrB/JiaW8) | USA | Tolland, Connecticut state | -72·4, 41·9 | 1997 - 2013 | inpatient | Unknown | Unknown | Model-based | Week |
| Noveroske, 2016 [[139]](https://paperpile.com/c/zPIKrB/JiaW8) | USA | Windham, Connecticut state | -72·2, 41·7 | 1997 - 2013 | inpatient | Unknown | Unknown | Model-based | Week |
| Nyoka, 2017 [[140]](https://paperpile.com/c/zPIKrB/hdTg7) | Kenya | Dadaab | 40·3, 0·1 | 2007 - 2011 | outpatient | ARI or ILI | Nucleic acid detection | Qualitative | Month |
| O'Kelly, 1991 [[141]](https://paperpile.com/c/zPIKrB/o3vf) | Republic of Ireland | Dublin | -6·3, 53·3 | 1987 - 1990 | inpatient | ARI or ILI | Antigen detection | Qualitative | Month |
| Obando-Pacheco, 2018 [[142]](https://paperpile.com/c/zPIKrB/h4sVb) | Argentina | nationwide | -63·6, -38·4 | 2011 - 2017 | Unknown | Unknown | Unknown | Threshold-based | Week |
| Obando-Pacheco, 2018 [[142]](https://paperpile.com/c/zPIKrB/h4sVb) | Australia | nationwide | 133·8, -25·3 | 2009 - 2016 | Unknown | Unknown | Unknown | Threshold-based | Week |
| Obando-Pacheco, 2018 [[142]](https://paperpile.com/c/zPIKrB/h4sVb) | Belgium | nationwide | 4·5, 50·5 | 2004 - 2014 | Unknown | Unknown | Unknown | Threshold-based | Week |
| Obando-Pacheco, 2018 [[142]](https://paperpile.com/c/zPIKrB/h4sVb) | Brazil | nationwide | -51·9, -14·2 | 2009 - 2017 | Unknown | Unknown | Unknown | Threshold-based | Week |
| Obando-Pacheco, 2018 [[142]](https://paperpile.com/c/zPIKrB/h4sVb) | Canada | nationwide | -106·3, 56·1 | 2012 - 2017 | Unknown | Unknown | Unknown | Threshold-based | Week |
| Obando-Pacheco, 2018 [[142]](https://paperpile.com/c/zPIKrB/h4sVb) | Chile | nationwide | -71·5, -35·7 | 2011 - 2017 | Unknown | Unknown | Unknown | Threshold-based | Week |
| Obando-Pacheco, 2018 [[142]](https://paperpile.com/c/zPIKrB/h4sVb) | China | nationwide | 104·2, 35·9 | 2010 - 2015 | Unknown | Unknown | Unknown | Threshold-based | Month |
| Obando-Pacheco, 2018 [[142]](https://paperpile.com/c/zPIKrB/h4sVb) | Finland | nationwide | 25·7, 61·9 | 2010 - 2015 | Unknown | Unknown | Unknown | Threshold-based | Week |
| Obando-Pacheco, 2018 [[142]](https://paperpile.com/c/zPIKrB/h4sVb) | France | nationwide | 2·7, 47·1 | 2011 - 2017 | Unknown | Unknown | Unknown | Threshold-based | Week |
| Obando-Pacheco, 2018 [[142]](https://paperpile.com/c/zPIKrB/h4sVb) | France | nationwide | 2·7, 47·1 | 2011 - 2017 | Unknown | Unknown | Unknown | Threshold-based | Month |
| Obando-Pacheco, 2018 [[142]](https://paperpile.com/c/zPIKrB/h4sVb) | Germany | nationwide | 9·7, 50·9 | 2010 - 2017 | Unknown | Unknown | Unknown | Threshold-based | Week |
| Obando-Pacheco, 2018 [[142]](https://paperpile.com/c/zPIKrB/h4sVb) | Germany | nationwide | 9·7, 50·9 | 2010 - 2017 | Unknown | Unknown | Unknown | Threshold-based | Week |
| Obando-Pacheco, 2018 [[142]](https://paperpile.com/c/zPIKrB/h4sVb) | Greece | nationwide | 21·8, 39·1 | 1999 - 2013 | Unknown | Unknown | Unknown | Threshold-based | Month |
| Obando-Pacheco, 2018 [[142]](https://paperpile.com/c/zPIKrB/h4sVb) | Guatemala | nationwide | -90·2, 15·8 | 2015 - 2017 | Unknown | Unknown | Unknown | Threshold-based | Week |
| Obando-Pacheco, 2018 [[142]](https://paperpile.com/c/zPIKrB/h4sVb) | Israel | nationwide | 34·9, 31 | 2005 - 2017 | Unknown | Unknown | Unknown | Threshold-based | Week |
| Obando-Pacheco, 2018 [[142]](https://paperpile.com/c/zPIKrB/h4sVb) | Italy | nationwide | 12·6, 41·9 | 2000 - 2014 | Unknown | Unknown | Unknown | Threshold-based | Month |
| Obando-Pacheco, 2018 [[142]](https://paperpile.com/c/zPIKrB/h4sVb) | Japan | nationwide | 138·3, 36·2 | 2010 - 2017 | Unknown | Unknown | Unknown | Threshold-based | Week |
| Obando-Pacheco, 2018 [[142]](https://paperpile.com/c/zPIKrB/h4sVb) | Malaysia | nationwide | 102, 4·2 | 1982 - 2008 | Unknown | Unknown | Unknown | Threshold-based | Month |
| Obando-Pacheco, 2018 [[142]](https://paperpile.com/c/zPIKrB/h4sVb) | Mexico | nationwide | -102·6, 23·6 | 2012 - 2015 | Unknown | Unknown | Unknown | Threshold-based | Week |
| Obando-Pacheco, 2018 [[142]](https://paperpile.com/c/zPIKrB/h4sVb) | Mozambique | nationwide | 35·5, -18·7 | 1998 - 2000 | Unknown | Unknown | Unknown | Threshold-based | Month |
| Obando-Pacheco, 2018 [[142]](https://paperpile.com/c/zPIKrB/h4sVb) | New Zealand | nationwide | 174·9, -40·9 | 2010 - 2015 | Unknown | Unknown | Unknown | Threshold-based | Week |
| Obando-Pacheco, 2018 [[142]](https://paperpile.com/c/zPIKrB/h4sVb) | Philippines | nationwide | 121·8, 12·9 | 2010 - 2013 | Unknown | Unknown | Unknown | Threshold-based | Month |
| Obando-Pacheco, 2018 [[142]](https://paperpile.com/c/zPIKrB/h4sVb) | South Africa | nationwide | 22·9, -30·6 | 2009 - 2016 | Unknown | Unknown | Unknown | Threshold-based | Week |
| Obando-Pacheco, 2018 [[142]](https://paperpile.com/c/zPIKrB/h4sVb) | South Korea | nationwide | 127·8, 35·9 | 2008 - 2016 | Unknown | Unknown | Unknown | Threshold-based | Month |
| Obando-Pacheco, 2018 [[142]](https://paperpile.com/c/zPIKrB/h4sVb) | Spain | nationwide | -3·3, 39·7 | 2010 - 2017 | Unknown | Unknown | Unknown | Threshold-based | Week |
| Obando-Pacheco, 2018 [[142]](https://paperpile.com/c/zPIKrB/h4sVb) | Thailand | nationwide | 101, 15·9 | 2005 - 2013 | Unknown | Unknown | Unknown | Threshold-based | Month |
| Obando-Pacheco, 2018 [[142]](https://paperpile.com/c/zPIKrB/h4sVb) | The Netherlands | nationwide | 5·3, 52·1 | 2010 - 2016 | Unknown | Unknown | Unknown | Threshold-based | Week |
| Obando-Pacheco, 2018 [[142]](https://paperpile.com/c/zPIKrB/h4sVb) | United Kingdom | nationwide | -1·6, 52·7 | 2010 - 2016 | Unknown | Unknown | Unknown | Threshold-based | Week |
| Obando-Pacheco, 2018 [[142]](https://paperpile.com/c/zPIKrB/h4sVb) | USA | nationwide | -95·7, 37·1 | 2011 - 2017 | Unknown | Unknown | Unknown | Threshold-based | Week |
| Oliveira-Santos, 2016 [[143]](https://paperpile.com/c/zPIKrB/uLso) | Portugal | Vila Real district | -7·7, 41·3 | 2005 - 2015 | inpatient | ALRI | Antigen detection | Qualitative | Month |
| Oskarsson, 2022 [[144]](https://paperpile.com/c/zPIKrB/zY6Hw) | Iceland | nationwide | -21·1, 64·4 | 2015 - 2020 | outpatient; inpatient | Unknown | Nucleic acid detection | Qualitative | Month |
| Paes, 2013 [[145]](https://paperpile.com/c/zPIKrB/ChHdE) | Canada | Hamilton | -79·9, 43·3 | 2002 - 2011 | outpatient; inpatient | Unknown | Antigen detection; Nucleic acid detection | Threshold-based | Week |
| Paiva, 2012 [[146]](https://paperpile.com/c/zPIKrB/PfEIA) | Brazil | Sao Paulo | -46·6, -23·6 | 1996 - 2010 | inpatient | ARI or ILI; ALRI | Antigen detection | Model-based | Month |
| Paiva, 2012 [[146]](https://paperpile.com/c/zPIKrB/PfEIA) | Brazil | Sao Paulo | -46·6, -23·6 | 1996 - 2010 | inpatient | ARI or ILI; ALRI | Antigen detection | Qualitative | Month |
| Panozzo, 2010 [[147]](https://paperpile.com/c/zPIKrB/6twje) | USA | Atlanta | -84·4, 33·7 | 2002 - 2007 | Unknown | Unknown | Virus detection; Antigen detection; Nucleic acid detection | Threshold-based | Week |
| Panozzo, 2010 [[147]](https://paperpile.com/c/zPIKrB/6twje) | USA | Birmingham | -86·8, 33·5 | 2002 - 2007 | Unknown | Unknown | Virus detection; Antigen detection; Nucleic acid detection | Threshold-based | Week |
| Panozzo, 2010 [[147]](https://paperpile.com/c/zPIKrB/6twje) | USA | Cleveland | -81·7, 41·5 | 2002 - 2007 | Unknown | Unknown | Virus detection; Antigen detection; Nucleic acid detection | Threshold-based | Week |
| Panozzo, 2010 [[147]](https://paperpile.com/c/zPIKrB/6twje) | USA | Columbia | -81, 34 | 2002 - 2007 | Unknown | Unknown | Virus detection; Antigen detection; Nucleic acid detection | Threshold-based | Week |
| Panozzo, 2010 [[147]](https://paperpile.com/c/zPIKrB/6twje) | USA | Corpus Christi | -97·4, 27·8 | 2002 - 2007 | Unknown | Unknown | Virus detection; Antigen detection; Nucleic acid detection | Threshold-based | Week |
| Panozzo, 2010 [[147]](https://paperpile.com/c/zPIKrB/6twje) | USA | Honolulu | -157·9, 21·3 | 2002 - 2007 | Unknown | Unknown | Virus detection; Antigen detection; Nucleic acid detection | Threshold-based | Week |
| Panozzo, 2010 [[147]](https://paperpile.com/c/zPIKrB/6twje) | USA | Indianapolis | -86·2, 39·8 | 2002 - 2007 | Unknown | Unknown | Virus detection; Antigen detection; Nucleic acid detection | Threshold-based | Week |
| Panozzo, 2010 [[147]](https://paperpile.com/c/zPIKrB/6twje) | USA | Long Beach | -118·2, 33·8 | 2002 - 2007 | Unknown | Unknown | Virus detection; Antigen detection; Nucleic acid detection | Threshold-based | Week |
| Panozzo, 2010 [[147]](https://paperpile.com/c/zPIKrB/6twje) | USA | Los Angeles | -118·2, 34·1 | 2002 - 2007 | Unknown | Unknown | Virus detection; Antigen detection; Nucleic acid detection | Threshold-based | Week |
| Panozzo, 2010 [[147]](https://paperpile.com/c/zPIKrB/6twje) | USA | Nashville | -86·8, 36·2 | 2002 - 2007 | Unknown | Unknown | Virus detection; Antigen detection; Nucleic acid detection | Threshold-based | Week |
| Panozzo, 2010 [[147]](https://paperpile.com/c/zPIKrB/6twje) | USA | New Orleans | -90·1, 30 | 2002 - 2007 | Unknown | Unknown | Virus detection; Antigen detection; Nucleic acid detection | Threshold-based | Week |
| Panozzo, 2010 [[147]](https://paperpile.com/c/zPIKrB/6twje) | USA | Oklahoma City | -97·5, 35·5 | 2002 - 2007 | Unknown | Unknown | Virus detection; Antigen detection; Nucleic acid detection | Threshold-based | Week |
| Panozzo, 2010 [[147]](https://paperpile.com/c/zPIKrB/6twje) | USA | Richmond | -77·4, 37·5 | 2002 - 2007 | Unknown | Unknown | Virus detection; Antigen detection; Nucleic acid detection | Threshold-based | Week |
| Panozzo, 2010 [[147]](https://paperpile.com/c/zPIKrB/6twje) | USA | San Antonio | -98·5, 29·4 | 2002 - 2007 | Unknown | Unknown | Virus detection; Antigen detection; Nucleic acid detection | Threshold-based | Week |
| Panozzo, 2010 [[147]](https://paperpile.com/c/zPIKrB/6twje) | USA | San Diego | -117·2, 32·7 | 2002 - 2007 | Unknown | Unknown | Virus detection; Antigen detection; Nucleic acid detection | Threshold-based | Week |
| Panozzo, 2010 [[147]](https://paperpile.com/c/zPIKrB/6twje) | USA | Seattle | -122·3, 47·6 | 2002 - 2007 | Unknown | Unknown | Virus detection; Antigen detection; Nucleic acid detection | Threshold-based | Week |
| Panozzo, 2010 [[147]](https://paperpile.com/c/zPIKrB/6twje) | USA | Sioux Falls | -96·7, 43·5 | 2002 - 2007 | Unknown | Unknown | Virus detection; Antigen detection; Nucleic acid detection | Threshold-based | Week |
| Panozzo, 2010 [[147]](https://paperpile.com/c/zPIKrB/6twje) | USA | St Louis | -90·2, 38·6 | 2002 - 2007 | Unknown | Unknown | Virus detection; Antigen detection; Nucleic acid detection | Threshold-based | Week |
| Panozzo, 2010 [[147]](https://paperpile.com/c/zPIKrB/6twje) | USA | St Louis | -90·2, 38·6 | 2002 - 2007 | Unknown | Unknown | Virus detection; Antigen detection; Nucleic acid detection | Threshold-based | Week |
| Paynter, 2015 [[148]](https://paperpile.com/c/zPIKrB/2gJR) | Australia | Cairns | 145·8, -16·9 | 1999 - 2012 | inpatient | Unknown | Unknown | Qualitative | Month |
| Paynter, 2015 [[148]](https://paperpile.com/c/zPIKrB/2gJR) | Australia | Townsville | 146·8, -19·3 | 1999 - 2012 | inpatient | Unknown | Unknown | Qualitative | Month |
| Pellegrinelli, 2022 [[149]](https://paperpile.com/c/zPIKrB/Ox5th) | Italy | nationwide | 12·6, 41·9 | 2014 - 2019 | outpatient | ARI or ILI | Unknown | Threshold-based | Week |
| Peterson, 2016 [[150]](https://paperpile.com/c/zPIKrB/2IZU) | Malawi | Blantyre | 35, -15·8 | 2011 - 2014 | Unknown | SARI | Nucleic acid detection | Qualitative | Month |
| Pierangeli, 2014 [[151]](https://paperpile.com/c/zPIKrB/5UR0) | Italy | Ancona | 13·5, 43·6 | 2010 - 2013 | inpatient | SARI | Nucleic acid detection | Qualitative | Month |
| Pierangeli, 2014 [[151]](https://paperpile.com/c/zPIKrB/5UR0) | Italy | Rome | 12·5, 41·9 | 2010 - 2013 | inpatient | SARI | Nucleic acid detection | Qualitative | Month |
| Price, 2019 [[152]](https://paperpile.com/c/zPIKrB/b012) | United Kingdom | Edinburgh | -3·2, 56 | 2009 - 2015 | Unknown | Unknown | Nucleic acid detection | Qualitative | Month |
| Ramaekers, 2017 [[153]](https://paperpile.com/c/zPIKrB/eCEh) | Belgium | Leuven | 4·7, 50·9 | 2011 - 2016 | Unknown | ARI or ILI | Nucleic acid detection | Qualitative | Week |
| Reeves, 2016 [[154]](https://paperpile.com/c/zPIKrB/3NLPe) | United Kingdom | England | -1·2, 52·4 | 2011 - 2014 | outpatient; inpatient | Unknown | Nucleic acid detection | Threshold-based | Week |
| Reiche, 2009 [[155]](https://paperpile.com/c/zPIKrB/lp0rv) | Germany | nationwide | 9·7, 50·9 | 1998 - 2007 | outpatient; inpatient | ARI or ILI | Nucleic acid detection | Threshold-based | Week |
| Renko, 2019 [[156]](https://paperpile.com/c/zPIKrB/pBuC) | Finland | nationwide | 25·7, 61·9 | 1995 - 2018 | Unknown | Unknown | Nucleic acid detection | Qualitative | Month |
| Reyes, 1997 [[157]](https://paperpile.com/c/zPIKrB/uo77Z) | Sweden | Stockholm | 18·1, 59·3 | 1984 - 1993 | inpatient | Unknown | Antigen detection | Threshold-based | Week |
| Richter, 2016 [[158]](https://paperpile.com/c/zPIKrB/NkWB) | Cyprus | Nicosia | 33·4, 35·2 | 2010 - 2013 | inpatient | ARI or ILI | Nucleic acid detection | Qualitative | Month |
| Rose, 2018 [[159]](https://paperpile.com/c/zPIKrB/l3Uth) | USA | Atlanta | -84·4, 33·7 | 2014 - 2017 | Unknown | Unknown | Virus detection; Antigen detection; Nucleic acid detection | Threshold-based | Week |
| Rose, 2018 [[159]](https://paperpile.com/c/zPIKrB/l3Uth) | USA | Boston | -71·1, 42·4 | 2014 - 2017 | Unknown | Unknown | Virus detection; Antigen detection; Nucleic acid detection | Threshold-based | Week |
| Rose, 2018 [[159]](https://paperpile.com/c/zPIKrB/l3Uth) | USA | Chicago | -87·6, 41·9 | 2014 - 2017 | Unknown | Unknown | Virus detection; Antigen detection; Nucleic acid detection | Threshold-based | Week |
| Rose, 2018 [[159]](https://paperpile.com/c/zPIKrB/l3Uth) | USA | Dellas | -96·8, 32·8 | 2014 - 2017 | Unknown | Unknown | Virus detection; Antigen detection; Nucleic acid detection | Threshold-based | Week |
| Rose, 2018 [[159]](https://paperpile.com/c/zPIKrB/l3Uth) | USA | Denver | -105, 39·7 | 2014 - 2017 | Unknown | Unknown | Virus detection; Antigen detection; Nucleic acid detection | Threshold-based | Week |
| Rose, 2018 [[159]](https://paperpile.com/c/zPIKrB/l3Uth) | USA | Kansas City | -94·6, 39·1 | 2014 - 2017 | Unknown | Unknown | Virus detection; Antigen detection; Nucleic acid detection | Threshold-based | Week |
| Rose, 2018 [[159]](https://paperpile.com/c/zPIKrB/l3Uth) | USA | nationwide | -95·7, 37·1 | 2014 - 2017 | Unknown | Unknown | Virus detection; Antigen detection; Nucleic acid detection | Threshold-based | Week |
| Rose, 2018 [[159]](https://paperpile.com/c/zPIKrB/l3Uth) | USA | nationwide | -95·7, 37·1 | 2014 - 2017 | Unknown | Unknown | Virus detection; Antigen detection; Nucleic acid detection | Threshold-based | Week |
| Rose, 2018 [[159]](https://paperpile.com/c/zPIKrB/l3Uth) | USA | New York | -74, 40·7 | 2014 - 2017 | Unknown | Unknown | Virus detection; Antigen detection; Nucleic acid detection | Threshold-based | Week |
| Rose, 2018 [[159]](https://paperpile.com/c/zPIKrB/l3Uth) | USA | Philadelphia | -75·2, 40 | 2014 - 2017 | Unknown | Unknown | Virus detection; Antigen detection; Nucleic acid detection | Threshold-based | Week |
| Rose, 2018 [[159]](https://paperpile.com/c/zPIKrB/l3Uth) | USA | San Francisco | -122·4, 37·8 | 2014 - 2017 | Unknown | Unknown | Virus detection; Antigen detection; Nucleic acid detection | Threshold-based | Week |
| Rose, 2018 [[159]](https://paperpile.com/c/zPIKrB/l3Uth) | USA | Seattle | -122·3, 47·6 | 2014 - 2017 | Unknown | Unknown | Virus detection; Antigen detection; Nucleic acid detection | Threshold-based | Week |
| Rose, 2020 [[160]](https://paperpile.com/c/zPIKrB/i2Xbo) | Kenya | Kilifi | 39·9, -3·6 | 2006 - 2018 | outpatient; inpatient | ARI or ILI; SARI; ALRI | Nucleic acid detection | Threshold-based | Week |
| Rose, 2020 [[160]](https://paperpile.com/c/zPIKrB/i2Xbo) | Kenya | Nairobi | 36·8, -1·3 | 2006 - 2016 | outpatient; inpatient | ARI or ILI; SARI; ALRI | Nucleic acid detection | Threshold-based | Week |
| Rose, 2020 [[160]](https://paperpile.com/c/zPIKrB/i2Xbo) | Kenya | Siaya | 34·3, 0·1 | 2006 - 2018 | outpatient; inpatient | ARI or ILI; SARI; ALRI | Nucleic acid detection | Threshold-based | Week |
| Rowlinson, 2017 [[161]](https://paperpile.com/c/zPIKrB/99ho) | Egypt | Damanhour | 30·5, 31 | 2009 - 2013 | inpatient | SARI | Nucleic acid detection | Qualitative | Month |
| Rzad, 2022 [[162]](https://paperpile.com/c/zPIKrB/R9I6x) | Poland | nationwide | 19·3, 51·7 | 2010 - 2020 | inpatient | Unknown | Unknown | Qualitative | Month |
| Sato, 2005 [[163]](https://paperpile.com/c/zPIKrB/yDbN) | Japan | Niigata city | 139, 37·9 | 2001 - 2004 | outpatient | ALRI | Nucleic acid detection | Qualitative | Month |
| Shobugawa, 2017 [[164]](https://paperpile.com/c/zPIKrB/dlg8O) | Japan | nationwide | 138·3, 36·2 | 2007 - 2014 | Unknown | Unknown | Unknown | Qualitative | Month |
| Singleton, 2007 [[165]](https://paperpile.com/c/zPIKrB/VnTAx) | USA | YK Delta | -164·2, 62·9 | 1996 - 2004 | inpatient | ALRI | Virus detection; Antigen detection | Threshold-based | Month |
| Sirimi, 2016 [[166]](https://paperpile.com/c/zPIKrB/65sk) | Greece | Athens | 23·7, 37·7 | 2002 - 2013 | inpatient | SARI | Antigen detection | Qualitative | Month |
| Sitthikarnkha, 2022 [[167]](https://paperpile.com/c/zPIKrB/snhh4) | Thailand | nationwide | 101, 15·9 | 2015 - 2020 | outpatient; inpatient | ALRI | Unknown | Qualitative | Month |
| Staadegaard, 2021 [[2]](https://paperpile.com/c/zPIKrB/aIPfL) | Brazil | Midwest | -51·9, -14·2 | 2016 - 2018 | inpatient | Unknown | Unknown | Coverage-based | Week |
| Staadegaard, 2021 [[2]](https://paperpile.com/c/zPIKrB/aIPfL) | Brazil | nationwide | -51·9, -14·2 | 2014 - 2018 | inpatient | Unknown | Unknown | Coverage-based | Week |
| Staadegaard, 2021 [[2]](https://paperpile.com/c/zPIKrB/aIPfL) | Brazil | Northeast | -42·6, -10·1 | 2014 - 2018 | inpatient | Unknown | Unknown | Coverage-based | Week |
| Staadegaard, 2021 [[2]](https://paperpile.com/c/zPIKrB/aIPfL) | Brazil | South | -49·3, -25·4 | 2014 - 2018 | inpatient | Unknown | Unknown | Coverage-based | Week |
| Staadegaard, 2021 [[2]](https://paperpile.com/c/zPIKrB/aIPfL) | Brazil | Southeast | -46·2, -20·3 | 2014 - 2018 | inpatient | Unknown | Unknown | Coverage-based | Week |
| Staadegaard, 2021 [[2]](https://paperpile.com/c/zPIKrB/aIPfL) | Chile | nationwide | -71·5, -35·7 | 2012 - 2018 | inpatient | SARI | Nucleic acid detection; Antigen detection | Coverage-based | Week |
| Staadegaard, 2021 [[2]](https://paperpile.com/c/zPIKrB/aIPfL) | Czech Republic | nationwide | 15·5, 49·8 | 2014 - 2018 | inpatient | Unknown | Virus detection; Antigen detection; Nucleic acid detection | Coverage-based | Week |
| Staadegaard, 2021 [[2]](https://paperpile.com/c/zPIKrB/aIPfL) | Ecuador | nationwide | -78·2, -1·8 | 2012 - 2018 | inpatient | SARI | Antigen detection | Coverage-based | Week |
| Staadegaard, 2021 [[2]](https://paperpile.com/c/zPIKrB/aIPfL) | New Zealand | nationwide | 174·9, -40·9 | 2012 - 2018 | outpatient | ARI or ILI | Nucleic acid detection | Coverage-based | Week |
| Staadegaard, 2021 [[2]](https://paperpile.com/c/zPIKrB/aIPfL) | Portugal | nationwide | -9·2, 39·7 | 2013 - 2018 | outpatient | ARI or ILI | Antigen detection; Nucleic acid detection | Coverage-based | Week |
| Staadegaard, 2021 [[2]](https://paperpile.com/c/zPIKrB/aIPfL) | Singapore | nationwide | 103·8, 1·4 | 2011 - 2018 | inpatient | SARI | Antigen detection; Nucleic acid detection | Coverage-based | Week |
| Staadegaard, 2021 [[2]](https://paperpile.com/c/zPIKrB/aIPfL) | South Africa | nationwide | 22·9, -30·6 | 2010 - 2018 | outpatient | ARI or ILI | Nucleic acid detection | Coverage-based | Week |
| Staadegaard, 2021 [[2]](https://paperpile.com/c/zPIKrB/aIPfL) | Spain | nationwide | -3·3, 39·7 | 2006 - 2018 | inpatient | Unknown | Unknown | Coverage-based | Week |
| Staadegaard, 2021 [[2]](https://paperpile.com/c/zPIKrB/aIPfL) | The Netherlands | nationwide | 5·3, 52·1 | 2000 - 2018 | inpatient | Unknown | Nucleic acid detection | Coverage-based | Week |
| Staadegaard, 2021 [[2]](https://paperpile.com/c/zPIKrB/aIPfL) | USA | HHS1 | 71·1, 42·4 | 2009 - 2018 | Unknown | Unknown | Virus detection; Antigen detection; Nucleic acid detection | Coverage-based | Week |
| Staadegaard, 2021 [[2]](https://paperpile.com/c/zPIKrB/aIPfL) | USA | HHS10 | 122·3, 47·6 | 2008 - 2018 | Unknown | Unknown | Virus detection; Antigen detection; Nucleic acid detection | Coverage-based | Week |
| Staadegaard, 2021 [[2]](https://paperpile.com/c/zPIKrB/aIPfL) | USA | HHS2 | 74, 40·7 | 2008 - 2018 | Unknown | Unknown | Virus detection; Antigen detection; Nucleic acid detection | Coverage-based | Week |
| Staadegaard, 2021 [[2]](https://paperpile.com/c/zPIKrB/aIPfL) | USA | HHS3 | 75·2, 40 | 2007 - 2018 | Unknown | Unknown | Virus detection; Antigen detection; Nucleic acid detection | Coverage-based | Week |
| Staadegaard, 2021 [[2]](https://paperpile.com/c/zPIKrB/aIPfL) | USA | HHS4 | 84·4, 33·8 | 2009 - 2018 | Unknown | Unknown | Virus detection; Antigen detection; Nucleic acid detection | Coverage-based | Week |
| Staadegaard, 2021 [[2]](https://paperpile.com/c/zPIKrB/aIPfL) | USA | HHS5 | 87·6, 41·9 | 2006 - 2018 | Unknown | Unknown | Virus detection; Antigen detection; Nucleic acid detection | Coverage-based | Week |
| Staadegaard, 2021 [[2]](https://paperpile.com/c/zPIKrB/aIPfL) | USA | HHS6 | 96·8, 32·8 | 2008 - 2018 | Unknown | Unknown | Virus detection; Antigen detection; Nucleic acid detection | Coverage-based | Week |
| Staadegaard, 2021 [[2]](https://paperpile.com/c/zPIKrB/aIPfL) | USA | HHS7 | 94·6, 39·1 | 2008 - 2018 | Unknown | Unknown | Virus detection; Antigen detection; Nucleic acid detection | Coverage-based | Week |
| Staadegaard, 2021 [[2]](https://paperpile.com/c/zPIKrB/aIPfL) | USA | HHS8 | 105, 39·7 | 2008 - 2018 | Unknown | Unknown | Virus detection; Antigen detection; Nucleic acid detection | Coverage-based | Week |
| Staadegaard, 2021 [[2]](https://paperpile.com/c/zPIKrB/aIPfL) | USA | HHS9 | 122·4, 37·8 | 2009 - 2018 | Unknown | Unknown | Virus detection; Antigen detection; Nucleic acid detection | Coverage-based | Week |
| Staadegaard, 2021 [[2]](https://paperpile.com/c/zPIKrB/aIPfL) | USA | nationwide | -95·7, 37·1 | 2006 - 2018 | Unknown | Unknown | Virus detection; Antigen detection; Nucleic acid detection | Coverage-based | Week |
| Stockman, 2013 [[168]](https://paperpile.com/c/zPIKrB/9Yq61) | Bangladesh | Dhaka | 90·4, 23·8 | 2004 - 2008 | outpatient | ARI or ILI | Nucleic acid detection | Qualitative | Month |
| Straliotto, 2001 [[169]](https://paperpile.com/c/zPIKrB/KWRS) | Brazil | Porto Alegre | -51·1, -31 | 1990 - 1998 | outpatient; inpatient | ARI or ILI; ALRI | Antigen detection | Qualitative | Month |
| Straliotto, 2002 [[170]](https://paperpile.com/c/zPIKrB/OHWs) | Brazil | Porto Alegre | -51·1, -31 | 1990 - 1992 | outpatient; inpatient | ARI or ILI;ALRI | Antigen detection | Qualitative | Month |
| Sundell, 2016 [[171]](https://paperpile.com/c/zPIKrB/f3Cy) | Sweden | Gothenburg | 12, 57·7 | 2010 - 2013 | outpatient; inpatient | ARI or ILI | Nucleic acid detection | Qualitative | Month |
| Sutmoller, 1995 [[172]](https://paperpile.com/c/zPIKrB/f9vki) | Brazil | Rio de Janeiro | -43·2, -22·9 | 1987 - 1989 | outpatient; inpatient | ARI or ILI | Antigen detection | Qualitative | Month |
| Tabatabai, 2022 [[173]](https://paperpile.com/c/zPIKrB/A1UTM) | Germany | Heidelberg | 8·7, 49·4 | 2014 - 2017 | inpatient | ARI or ILI | Nucleic acid detection | Qualitative | Month |
| Tan, 2021 [[174]](https://paperpile.com/c/zPIKrB/vTj3) | Singapore | nationwide | 103·8, 1·4 | 2011 - 2016 | inpatient | ARI or ILI | Antigen detection | Qualitative | Month |
| Tang, 2010 [[175]](https://paperpile.com/c/zPIKrB/wB8zW) | Hong Kong | nationwide | 114·2, 22·3 | 2000 - 2007 | inpatient | ARI or ILI | Antigen detection | Qualitative | Month |
| Terletskaia-Ladwig, 2005 [[176]](https://paperpile.com/c/zPIKrB/KGT1J) | Germany | Stuttgart | 9·2, 48·8 | 1996 - 2004 | outpatient; inpatient | Clinical judgment | Antigen detection; Nucleic acid detection | Threshold-based | Week |
| Thomas, 1994 [[177]](https://paperpile.com/c/zPIKrB/vvvR) | Canada | British Columbia | -127·6, 53·7 | 1987 - 1992 | inpatient | ARI or ILI | Antigen detection | Qualitative | Month |
| Thongpan, 2020 [[178]](https://paperpile.com/c/zPIKrB/tlHj) | Thailand | nationwide | 101, 15·9 | 2012 - 2018 | Unknown | ARI or ILI | Nucleic acid detection | Qualitative | Month |
| Thwaites, 2020 [[179]](https://paperpile.com/c/zPIKrB/DTr5) | United Kingdom | Scoland | -4·2, 56·5 | 2000 - 2011 | inpatient | Unknown | Unknown | Qualitative | Month |
| Tsolia, 2003 [[180]](https://paperpile.com/c/zPIKrB/TMXA) | Greece | Athens | 23·7, 37·7 | 1997 - 2000 | inpatient | ALRI | Antigen detection | Qualitative | Month |
| Turner, 2012 [[181]](https://paperpile.com/c/zPIKrB/Y1v0) | Thailand | Maela camp | 98·4, 17·2 | 2007 - 2010 | outpatient | ALRI | Nucleic acid detection | Qualitative | Month |
| Ucakar, 2013 [[182]](https://paperpile.com/c/zPIKrB/dkxqZ) | Slovenia | nationwide | 14·9, 46·2 | 2006 - 2011 | outpatient; inpatient | ARI or ILI | Antigen detection; Nucleic acid detection | Threshold-based | Week |
| Valley-Omar, 2022* [[183]](https://paperpile.com/c/zPIKrB/1Q5Yl) | South Africa | KwaZulu-Natal province | 30·9, -28·5 | 2012 - 2015 | inpatient | SARI | Nucleic acid detection | Qualitative | Month |
| Valley-Omar, 2022* [[183]](https://paperpile.com/c/zPIKrB/1Q5Yl) | South Africa | North West Province | 25·3, -26·7 | 2012 - 2015 | inpatient | SARI | Nucleic acid detection | Qualitative | Month |
| van der Sande, 2004 [[184]](https://paperpile.com/c/zPIKrB/YMZz) | Gambia | Banjul | -16·6, 13·5 | 1993 - 2002 | inpatient | SARI | Antigen detection | Qualitative | Month |
| van Summeren, 2021 [[185]](https://paperpile.com/c/zPIKrB/HoDc) | France | France | 2·7, 47·1 | 2016 - 2021 | outpatient; inpatient | ARI or ILI; Clinical judgment | Virus detection; Antibody detection; Antigen detection; Nucleic acid detection | Qualitative | Week |
| van Summeren, 2021 [[185]](https://paperpile.com/c/zPIKrB/HoDc) | Iceland | Iceland | -21·1, 64·4 | 2016 - 2021 | outpatient; inpatient | ARI or ILI; Clinical judgment | Virus detection; Antibody detection; Antigen detection; Nucleic acid detection | Qualitative | Week |
| van Summeren, 2021 [[185]](https://paperpile.com/c/zPIKrB/HoDc) | The Netherlands | The Netherlands | 5·3, 52·1 | 2016 - 2021 | outpatient; inpatient | ARI or ILI; Clinical judgment | Virus detection; Antibody detection; Antigen detection; Nucleic acid detection | Qualitative | Week |
| Vandini, 2013 [[186]](https://paperpile.com/c/zPIKrB/4NIp) | Italy | Bologna | 11·3, 44·5 | 2007 - 2010 | outpatient | ARI or ILI | Antigen detection | Qualitative | Week |
| Verani, 2013 [[187]](https://paperpile.com/c/zPIKrB/el7n) | Guatemala | Santa Rosa, Guateala and Quetzaltenango | -90·4, 14·2 | 2007 - 2011 | inpatient | ARI or ILI | Nucleic acid detection | Qualitative | Month |
| Viegas, 2004 [[188]](https://paperpile.com/c/zPIKrB/OHDD) | Argentina | Buenos Aires city and Greater Buenos Aires | -58·4, -34·6 | 1998 - 2002 | inpatient | ALRI | Antigen detection | Qualitative | Month |
| Viguria, 2018 [[189]](https://paperpile.com/c/zPIKrB/vwxwK) | Spain | Navarra | -1·7, 42·7 | 2010 - 2015 | inpatient | Unknown | Antigen detection; Nucleic acid detection | Threshold-based | Week |
| Vila, 2022 [[190]](https://paperpile.com/c/zPIKrB/QTdsL) | Spain | Barcelona | 2·2, 41·4 | 2012 - 2020 | outpatient; inpatient | ALRI | Antigen detection; Nucleic acid detection | Qualitative | Month |
| Vos, 2019 [[191]](https://paperpile.com/c/zPIKrB/6BC5G) | The Netherlands | nationwide | 5·3, 52·1 | 2005 - 2017 | outpatient; inpatient | ARI or ILI | Virus detection; Antibody detection; Antigen detection; Nucleic acid detection | Coverage-based | Week |
| Vos, 2019 [[191]](https://paperpile.com/c/zPIKrB/6BC5G) | The Netherlands | nationwide | 5·3, 52·1 | 2005 - 2017 | outpatient; inpatient | ARI or ILI | Virus detection; Antibody detection; Antigen detection; Nucleic acid detection | Threshold-based | Week |
| Vos, 2019 [[191]](https://paperpile.com/c/zPIKrB/6BC5G) | The Netherlands | nationwide | 5·3, 52·1 | 2005 - 2017 | outpatient; inpatient | ARI or ILI | Virus detection; Antibody detection; Antigen detection; Nucleic acid detection | Coverage-based | Week |
| Vos, 2019 [[191]](https://paperpile.com/c/zPIKrB/6BC5G) | The Netherlands | nationwide | 5·3, 52·1 | 2005 - 2017 | outpatient; inpatient | ARI or ILI | Virus detection; Antibody detection; Antigen detection; Nucleic acid detection | Threshold-based | Week |
| Wagatsuma, 2021 [[192]](https://paperpile.com/c/zPIKrB/Yt6m7) | Japan | nationwide | 138·3, 36·2 | 2014 - 2017 | Unknown | Unknown | Unknown | Coverage-based | Month |
| Wahab, 2001 [[193]](https://paperpile.com/c/zPIKrB/g9zZ) | Qatar | Doha | 51·5, 25·3 | 1996 - 1998 | inpatient | Unknown | Antigen detection | Qualitative | Month |
| Wang, 2022 [[194]](https://paperpile.com/c/zPIKrB/an6hH) | USA | nationwide | -95·7, 37·1 | 2010 - 2022 | Unknown | Unknown | Unknown | Qualitative | Month |
| Weber, 1998 [[195]](https://paperpile.com/c/zPIKrB/4DDq) | Gambia | Western Region | -16·6, 13·2 | 1993 - 1996 | inpatient | ALRI | Antigen detection | Qualitative | Month |
| Weigl, 2000 [[196]](https://paperpile.com/c/zPIKrB/n9ew) | Germany | Kiel | 10·1, 54·3 | 1995 - 1999 | inpatient | ARI or ILI | Nucleic acid detection | Qualitative | Month |
| Weigl, 2002 [[197]](https://paperpile.com/c/zPIKrB/7fCuW) | Germany | Kiel | 10·1, 54·3 | 1994 - 2001 | inpatient | SARI | Antigen detection; Nucleic acid detection | Qualitative | Month |
| Weigl, 2002 [[197]](https://paperpile.com/c/zPIKrB/7fCuW) | Germany | Kiel | 10·1, 54·3 | 1994 - 2001 | inpatient | SARI | Antigen detection; Nucleic acid detection | Qualitative | Month |
| Weissenbacher, 1990 [[198]](https://paperpile.com/c/zPIKrB/xcs0) | Argentina | Buenos Aires | -58·4, -34·6 | 1984 - 1987 | outpatient; inpatient | ALRI | Antigen detection | Qualitative | Month |
| Wilfret, 2008 [[199]](https://paperpile.com/c/zPIKrB/pdOs) | USA | North Carolina | -79, 35·8 | 2003 - 2006 | outpatient; inpatient | Unknown | Antigen detection | Threshold-based | Month |
| Winter, 1996 [[200]](https://paperpile.com/c/zPIKrB/rzNg) | United Kingdom | Edinburgh | -3·2, 56 | 1985 - 1994 | inpatient | Unknown | Virus detection | Qualitative | Month |
| Wrotek, 2020 [[201]](https://paperpile.com/c/zPIKrB/u8hZN) | Poland | nationwide | 19·3, 51·7 | 2010 - 2017 | inpatient | Unknown | Unknown | Threshold-based | Week |
| Wrotek, 2020 [[201]](https://paperpile.com/c/zPIKrB/u8hZN) | Poland | Warsaw | 21, 52·2 | 2010 - 2017 | inpatient | Unknown | Unknown | Threshold-based | Month |
| Yamagami, 2019 [[202]](https://paperpile.com/c/zPIKrB/hfrAP) | Japan | Aichi prefecture | 137·3, 35 | 2012 - 2018 | Unknown | Unknown | Antigen detection; Nucleic acid detection | Coverage-based | Week |
| Yamagami, 2019 [[202]](https://paperpile.com/c/zPIKrB/hfrAP) | Japan | Akita prefecture | 140·3, 40·1 | 2012 - 2018 | Unknown | Unknown | Antigen detection; Nucleic acid detection | Coverage-based | Week |
| Yamagami, 2019 [[202]](https://paperpile.com/c/zPIKrB/hfrAP) | Japan | Aomori prefecture | 140·9, 40·8 | 2012 - 2018 | Unknown | Unknown | Antigen detection; Nucleic acid detection | Coverage-based | Week |
| Yamagami, 2019 [[202]](https://paperpile.com/c/zPIKrB/hfrAP) | Japan | Chiba prefecture | 140·2, 35·3 | 2012 - 2018 | Unknown | Unknown | Antigen detection; Nucleic acid detection | Coverage-based | Week |
| Yamagami, 2019 [[202]](https://paperpile.com/c/zPIKrB/hfrAP) | Japan | Ehime prefecture | 132·8, 33·6 | 2012 - 2018 | Unknown | Unknown | Antigen detection; Nucleic acid detection | Coverage-based | Week |
| Yamagami, 2019 [[202]](https://paperpile.com/c/zPIKrB/hfrAP) | Japan | Fukui prefecture | 136·2, 35·9 | 2012 - 2018 | Unknown | Unknown | Antigen detection; Nucleic acid detection | Coverage-based | Week |
| Yamagami, 2019 [[202]](https://paperpile.com/c/zPIKrB/hfrAP) | Japan | Fukuoka prefecture | 130·7, 33·6 | 2012 - 2018 | Unknown | Unknown | Antigen detection; Nucleic acid detection | Coverage-based | Week |
| Yamagami, 2019 [[202]](https://paperpile.com/c/zPIKrB/hfrAP) | Japan | Fukushima prefecture | 140·2, 37·4 | 2012 - 2018 | Unknown | Unknown | Antigen detection; Nucleic acid detection | Coverage-based | Week |
| Yamagami, 2019 [[202]](https://paperpile.com/c/zPIKrB/hfrAP) | Japan | Gifu prefecture | 137, 35·7 | 2012 - 2018 | Unknown | Unknown | Antigen detection; Nucleic acid detection | Coverage-based | Week |
| Yamagami, 2019 [[202]](https://paperpile.com/c/zPIKrB/hfrAP) | Japan | Gunma prefecture | 138·9, 36·6 | 2012 - 2018 | Unknown | Unknown | Antigen detection; Nucleic acid detection | Coverage-based | Week |
| Yamagami, 2019 [[202]](https://paperpile.com/c/zPIKrB/hfrAP) | Japan | Hiroshima prefecture | 133, 34·9 | 2012 - 2018 | Unknown | Unknown | Antigen detection; Nucleic acid detection | Coverage-based | Week |
| Yamagami, 2019 [[202]](https://paperpile.com/c/zPIKrB/hfrAP) | Japan | Hokkaido prefecture | 142·9, 43·2 | 2012 - 2018 | Unknown | Unknown | Antigen detection; Nucleic acid detection | Coverage-based | Week |
| Yamagami, 2019 [[202]](https://paperpile.com/c/zPIKrB/hfrAP) | Japan | Hyogo prefecture | 134·5, 34·9 | 2012 - 2018 | Unknown | Unknown | Antigen detection; Nucleic acid detection | Coverage-based | Week |
| Yamagami, 2019 [[202]](https://paperpile.com/c/zPIKrB/hfrAP) | Japan | Ibaraki prefecture | 140·2, 36·2 | 2012 - 2018 | Unknown | Unknown | Antigen detection; Nucleic acid detection | Coverage-based | Week |
| Yamagami, 2019 [[202]](https://paperpile.com/c/zPIKrB/hfrAP) | Japan | Ishikawa prefecture | 136·5, 36·3 | 2012 - 2018 | Unknown | Unknown | Antigen detection; Nucleic acid detection | Coverage-based | Week |
| Yamagami, 2019 [[202]](https://paperpile.com/c/zPIKrB/hfrAP) | Japan | Iwate prefecture | 141·3, 39·6 | 2012 - 2018 | Unknown | Unknown | Antigen detection; Nucleic acid detection | Coverage-based | Week |
| Yamagami, 2019 [[202]](https://paperpile.com/c/zPIKrB/hfrAP) | Japan | Kagawa prefecture | 134, 34·2 | 2012 - 2018 | Unknown | Unknown | Antigen detection; Nucleic acid detection | Coverage-based | Week |
| Yamagami, 2019 [[202]](https://paperpile.com/c/zPIKrB/hfrAP) | Japan | Kagoshima prefecture | 130·9, 31·4 | 2012 - 2018 | Unknown | Unknown | Antigen detection; Nucleic acid detection | Coverage-based | Week |
| Yamagami, 2019 [[202]](https://paperpile.com/c/zPIKrB/hfrAP) | Japan | Kanagawa prefecture | 139·3, 35·5 | 2012 - 2018 | Unknown | Unknown | Antigen detection; Nucleic acid detection | Coverage-based | Week |
| Yamagami, 2019 [[202]](https://paperpile.com/c/zPIKrB/hfrAP) | Japan | Kochi prefecture | 133·3, 33·5 | 2012 - 2018 | Unknown | Unknown | Antigen detection; Nucleic acid detection | Coverage-based | Week |
| Yamagami, 2019 [[202]](https://paperpile.com/c/zPIKrB/hfrAP) | Japan | Kumamoto prefecture | 130·8, 32·9 | 2012 - 2018 | Unknown | Unknown | Antigen detection; Nucleic acid detection | Coverage-based | Week |
| Yamagami, 2019 [[202]](https://paperpile.com/c/zPIKrB/hfrAP) | Japan | kyoto prefecture | 135·5, 35·2 | 2012 - 2018 | Unknown | Unknown | Antigen detection; Nucleic acid detection | Coverage-based | Week |
| Yamagami, 2019 [[202]](https://paperpile.com/c/zPIKrB/hfrAP) | Japan | Mie prefecture | 136, 33·8 | 2012 - 2018 | Unknown | Unknown | Antigen detection; Nucleic acid detection | Coverage-based | Week |
| Yamagami, 2019 [[202]](https://paperpile.com/c/zPIKrB/hfrAP) | Japan | Miyagi prefecture | 141·1, 38·6 | 2012 - 2018 | Unknown | Unknown | Antigen detection; Nucleic acid detection | Coverage-based | Week |
| Yamagami, 2019 [[202]](https://paperpile.com/c/zPIKrB/hfrAP) | Japan | Miyazaki prefecture | 131·4, 32·6 | 2012 - 2018 | Unknown | Unknown | Antigen detection; Nucleic acid detection | Coverage-based | Week |
| Yamagami, 2019 [[202]](https://paperpile.com/c/zPIKrB/hfrAP) | Japan | Nagano prefecture | 137·9, 36·2 | 2012 - 2018 | Unknown | Unknown | Antigen detection; Nucleic acid detection | Coverage-based | Week |
| Yamagami, 2019 [[202]](https://paperpile.com/c/zPIKrB/hfrAP) | Japan | Nagasaki prefecture | 129·7, 33·2 | 2012 - 2018 | Unknown | Unknown | Antigen detection; Nucleic acid detection | Coverage-based | Week |
| Yamagami, 2019 [[202]](https://paperpile.com/c/zPIKrB/hfrAP) | Japan | Nara prefecture | 135·8, 34·3 | 2012 - 2018 | Unknown | Unknown | Antigen detection; Nucleic acid detection | Coverage-based | Week |
| Yamagami, 2019 [[202]](https://paperpile.com/c/zPIKrB/hfrAP) | Japan | Niigata prefecture | 138·9, 37·5 | 2012 - 2018 | Unknown | Unknown | Antigen detection; Nucleic acid detection | Coverage-based | Week |
| Yamagami, 2019 [[202]](https://paperpile.com/c/zPIKrB/hfrAP) | Japan | Oita prefecture | 131·4, 33·2 | 2012 - 2018 | Unknown | Unknown | Antigen detection; Nucleic acid detection | Coverage-based | Week |
| Yamagami, 2019 [[202]](https://paperpile.com/c/zPIKrB/hfrAP) | Japan | Okayama prefecture | 133·6, 34·9 | 2012 - 2018 | Unknown | Unknown | Antigen detection; Nucleic acid detection | Coverage-based | Week |
| Yamagami, 2019 [[202]](https://paperpile.com/c/zPIKrB/hfrAP) | Japan | Okinawa prefecture | 127·7, 26·1 | 2012 - 2018 | Unknown | Unknown | Antigen detection; Nucleic acid detection | Coverage-based | Week |
| Yamagami, 2019 [[202]](https://paperpile.com/c/zPIKrB/hfrAP) | Japan | Osaka prefecture | 135·6, 34·6 | 2012 - 2018 | Unknown | Unknown | Antigen detection; Nucleic acid detection | Coverage-based | Week |
| Yamagami, 2019 [[202]](https://paperpile.com/c/zPIKrB/hfrAP) | Japan | Saga prefecture | 130·2, 33·3 | 2012 - 2018 | Unknown | Unknown | Antigen detection; Nucleic acid detection | Coverage-based | Week |
| Yamagami, 2019 [[202]](https://paperpile.com/c/zPIKrB/hfrAP) | Japan | Saitama prefecture | 139·4, 36 | 2012 - 2018 | Unknown | Unknown | Antigen detection; Nucleic acid detection | Coverage-based | Week |
| Yamagami, 2019 [[202]](https://paperpile.com/c/zPIKrB/hfrAP) | Japan | Shiga prefecture | 136·1, 35·3 | 2012 - 2018 | Unknown | Unknown | Antigen detection; Nucleic acid detection | Coverage-based | Week |
| Yamagami, 2019 [[202]](https://paperpile.com/c/zPIKrB/hfrAP) | Japan | Shimane prefecture | 132·6, 35·1 | 2012 - 2018 | Unknown | Unknown | Antigen detection; Nucleic acid detection | Coverage-based | Week |
| Yamagami, 2019 [[202]](https://paperpile.com/c/zPIKrB/hfrAP) | Japan | Shizuoka prefecture | 138·3, 35·1 | 2012 - 2018 | Unknown | Unknown | Antigen detection; Nucleic acid detection | Coverage-based | Week |
| Yamagami, 2019 [[202]](https://paperpile.com/c/zPIKrB/hfrAP) | Japan | Tochigi prefecture | 139·9, 36·7 | 2012 - 2018 | Unknown | Unknown | Antigen detection; Nucleic acid detection | Coverage-based | Week |
| Yamagami, 2019 [[202]](https://paperpile.com/c/zPIKrB/hfrAP) | Japan | Tokushima prefecture | 134·3, 33·9 | 2012 - 2018 | Unknown | Unknown | Antigen detection; Nucleic acid detection | Coverage-based | Week |
| Yamagami, 2019 [[202]](https://paperpile.com/c/zPIKrB/hfrAP) | Japan | Tokyo | 139·7, 35·7 | 2012 - 2018 | Unknown | Unknown | Antigen detection; Nucleic acid detection | Coverage-based | Week |
| Yamagami, 2019 [[202]](https://paperpile.com/c/zPIKrB/hfrAP) | Japan | Tottori prefecture | 133·4, 35·4 | 2012 - 2018 | Unknown | Unknown | Antigen detection; Nucleic acid detection | Coverage-based | Week |
| Yamagami, 2019 [[202]](https://paperpile.com/c/zPIKrB/hfrAP) | Japan | Toyama prefecture | 137·2, 36·7 | 2012 - 2018 | Unknown | Unknown | Antigen detection; Nucleic acid detection | Coverage-based | Week |
| Yamagami, 2019 [[202]](https://paperpile.com/c/zPIKrB/hfrAP) | Japan | Wakayama prefecture | 135·4, 33·9 | 2012 - 2018 | Unknown | Unknown | Antigen detection; Nucleic acid detection | Coverage-based | Week |
| Yamagami, 2019 [[202]](https://paperpile.com/c/zPIKrB/hfrAP) | Japan | Yamagata prefecture | 140·1, 38·5 | 2012 - 2018 | Unknown | Unknown | Antigen detection; Nucleic acid detection | Coverage-based | Week |
| Yamagami, 2019 [[202]](https://paperpile.com/c/zPIKrB/hfrAP) | Japan | Yamaguchi prefecture | 131·5, 34·3 | 2012 - 2018 | Unknown | Unknown | Antigen detection; Nucleic acid detection | Coverage-based | Week |
| Yamagami, 2019 [[202]](https://paperpile.com/c/zPIKrB/hfrAP) | Japan | Yamanashi prefecture | 138·6, 35·7 | 2012 - 2018 | Unknown | Unknown | Antigen detection; Nucleic acid detection | Coverage-based | Week |
| Yokosawa, 2006 [[203]](https://paperpile.com/c/zPIKrB/k2Xb) | Brazil | Uberlândia | -48·3, -18·9 | 2001 - 2004 | outpatient; inpatient | ARI or ILI | Antigen detection | Qualitative | Month |
| Yorita, 2007 [[204]](https://paperpile.com/c/zPIKrB/29BE) | USA | Hawaii | -155·6, 19·9 | 1997 - 2004 | inpatient | ALRI | Unknown | Qualitative | Month |
| Yu, 2019 [[205]](https://paperpile.com/c/zPIKrB/5XvIE) | China | Beijing | 116·4, 39·9 | 2007 - 2015 | inpatient | ALRI | Nucleic acid detection | Threshold-based | Week |
| Yu, 2019 [[205]](https://paperpile.com/c/zPIKrB/5XvIE) | China | Beijing | 116·4, 39·9 | 2007 - 2015 | inpatient | ALRI | Nucleic acid detection | Threshold-based | Week |
| Yu, 2019 [[205]](https://paperpile.com/c/zPIKrB/5XvIE) | China | Beijing | 116·4, 39·9 | 2007 - 2015 | inpatient | ALRI | Nucleic acid detection | Threshold-based | Week |
| Yusuf, 2007 [[206]](https://paperpile.com/c/zPIKrB/riwp) | Canada | Winnipeg | -97·1, 49·9 | 2002 - 2004 | Unknown | Unknown | Virus detection; Antigen detection | Qualitative | Month |
| Yusuf, 2007 [[206]](https://paperpile.com/c/zPIKrB/riwp) | Chile | Santiago | -70·7, -33·4 | 1999 - 2003 | Unknown | Unknown | Virus detection; Antigen detection | Qualitative | Month |
| Yusuf, 2007 [[206]](https://paperpile.com/c/zPIKrB/riwp) | USA | Buffalo | -78·9, 42·9 | 1995 - 2002 | Unknown | Unknown | Virus detection; Antigen detection | Qualitative | Month |
| Yusuf, 2007 [[206]](https://paperpile.com/c/zPIKrB/riwp) | USA | Houston | -95·4, 29·8 | 1999 - 2002 | Unknown | Unknown | Virus detection; Antigen detection | Qualitative | Month |
| Yusuf, 2007 [[206]](https://paperpile.com/c/zPIKrB/riwp) | USA | Miami | -80·2, 25·8 | 2000 - 2003 | Unknown | Unknown | Virus detection; Antigen detection | Qualitative | Month |
| Yusuf, 2007 [[206]](https://paperpile.com/c/zPIKrB/riwp) | USA | Tucson | -111, 32·2 | 1999 - 2003 | Unknown | Unknown | Virus detection; Antigen detection | Qualitative | Month |
| Zhang, 2010 [[207]](https://paperpile.com/c/zPIKrB/6P2a) | China | Chongqing | 106·6, 29·6 | 2006 - 2009 | inpatient | ARI or ILI | Nucleic acid detection | Qualitative | Month |
| Zhang, 2013 [[208]](https://paperpile.com/c/zPIKrB/K43M) | China | Suzhou | 120·6, 31·3 | 2001 - 2011 | inpatient | ARI or ILI | Antigen detection | Qualitative | Month |
| Zhao, 2014 [[209]](https://paperpile.com/c/zPIKrB/rGdI) | United Kingdom | nationwide | -1·6, 52·7 | 2009 - 2012 | Unknown | Unknown | Nucleic acid detection | Qualitative | Month |
| Zlateva, 2007 [[210]](https://paperpile.com/c/zPIKrB/vjMv) | Belgium | Leuven | 4·7, 50·9 | 1996 - 2006 | outpatient; inpatient | SARI | Virus detection; Antigen detection; Nucleic acid detection | Qualitative | Month |

Note: ARI, acute respiratory infection; ILI, influenza-like illness; SARI, severe acute respiratory infection; ALRI, acute lower respiratory infection

[**Supplementary Table**](https://docs.google.com/document/d/1I4dWqDo4EhvkiX1s2J-0H5fuGgVyt68VTcB-H1RzDxY/edit#stabl_3) **2.** Methods used to determine the timing (the start, peak and end) of RSV seasons in the included publications.

| **Method/Method Category** | **Number of studies (n=59)** | **References** |
| --- | --- | --- |
| Threshold-based method | 51 | [7,14,16,17,22,28,29,33,36–38,44,47,57,61,65,66,69,71,73, 75–77,80–83,93,99,106,110,111,124,125,127,135,138,142, 145,147,149,154,155,157,159,160,165,176,182,189,191,199,  201,205] |
| Coverage-based methods  (AAP, Search Index, MEM) | 8 | [2,3,31,77,109,191,192,202] |
| Model-based methods  (Change point model, Over-dispersed Poisson regression, Expectation-based Poisson scan statistics, Time series methods) | 6 | [4,5,107,129,139,146] |

[**Supplementary Table**](https://docs.google.com/document/d/1I4dWqDo4EhvkiX1s2J-0H5fuGgVyt68VTcB-H1RzDxY/edit#stabl_4) **3**. Characteristics of the threshold-based methods used to determine the season of RSV activity.

| **Method description** | | | **Number of studies (n=59)** | **References** |
| --- | --- | --- | --- | --- |
| **Prime indicator** | **Threshold** | **Other requirements** |  |  |
| Positive percentage | 10% | None | 6 | [[7,28,99,106,111,205]](https://paperpile.com/c/zPIKrB/T5MU6+UJyei+XkHEI+vDVJf+5XvIE+HZLo) |
|  |  | continuity^a^ ≥ 2 | 10 | [[33,44,81–83,110,135,142,149,154]](https://paperpile.com/c/zPIKrB/GMmUc+yqMda+VCs9C+80EEw+u2aTv+7vWjg+HrPoU+h4sVb+3NLPe+Ox5th) |
|  |  | continuity ≥ 2, positive samples ≥ 2 | 5 | [[65,73,80,138,165]](https://paperpile.com/c/zPIKrB/QzfFW+o3kmc+23GWJ+Raleq+VnTAx) |
|  |  | continuity ≥ 2, tests ≥ 11 | 2 | [[17,124]](https://paperpile.com/c/zPIKrB/aBsPn+gWGAj) |
|  |  | tests ≥ 10 | 2 | [[125,199]](https://paperpile.com/c/zPIKrB/br0sv+pdOs) |
|  |  | tests ≥ 20 | 3 | [[145,147,182]](https://paperpile.com/c/zPIKrB/6twje+ChHdE+dkxqZ) |
|  | 1% | None | 1 | [[61]](https://paperpile.com/c/zPIKrB/CpdqX) |
|  | 3% | continuity ≥ 2 | 3 | [[17,77,127]](https://paperpile.com/c/zPIKrB/gWGAj+1iYBm+LX59k) |
|  | 5% | None | 1 | [[77]](https://paperpile.com/c/zPIKrB/1iYBm) |
|  |  | continuity ≥ 2 | 1 | [[38]](https://paperpile.com/c/zPIKrB/42Pvu) |
|  | 7% | None | 1 | [[77]](https://paperpile.com/c/zPIKrB/1iYBm) |
|  | mean of the 5-week moving average | continuity >=3 | 1 | [[160]](https://paperpile.com/c/zPIKrB/i2Xbo) |
|  | threefold of the median | None | 1 | [[57]](https://paperpile.com/c/zPIKrB/vNcVL) |
|  | mean | continuity ≥ 2 | 1 | [[29]](https://paperpile.com/c/zPIKrB/StFDy) |
| RSV cases | 0 | continuity ≥ 3 | 1 | [[155]](https://paperpile.com/c/zPIKrB/lp0rv) |
|  | 2 (per week)  5 (per month) | None | 2 | [[157,176]](https://paperpile.com/c/zPIKrB/uo77Z+KGT1J) |
|  | 20 | None | 1 | [[191]](https://paperpile.com/c/zPIKrB/6BC5G) |
|  | 100 | None | 2 | [[69,76]](https://paperpile.com/c/zPIKrB/XvW5a+kKmQz) |
|  | 200 | None | 2 | [[69,71]](https://paperpile.com/c/zPIKrB/rMNIq+kKmQz) |
|  | 10% of peak | continuity ≥ 2 | 1 | [[82]](https://paperpile.com/c/zPIKrB/GMmUc) |
|  | 5% of the total number | continuity ≥ 2 | 1 | [[22]](https://paperpile.com/c/zPIKrB/Z8Prv) |
|  | 1·2% of the total cases of that year | None | 3 | [[36,77,191]](https://paperpile.com/c/zPIKrB/6BC5G+1iYBm+Alr49) |
|  | 60% of weekly average cases of that year | None | 2 | [[66,77]](https://paperpile.com/c/zPIKrB/YkTtU+1iYBm) |
|  | two times the average weekly number | None | 1 | [[22]](https://paperpile.com/c/zPIKrB/Z8Prv) |
|  | two times the average weekly number | cases ≥ 5 | 1 | [[47]](https://paperpile.com/c/zPIKrB/sdUN2) |
|  | 10 times the 4-week  moving average at week 29 | continuity ≥ 2 | 1 | [[127]](https://paperpile.com/c/zPIKrB/LX59k) |
|  | 1.24-fold mean of previous 5 months | None | 1 | [[14]](https://paperpile.com/c/zPIKrB/9fNn0) |
| Normalized increase | 10 | continuity ≥ 2 | 2 | [[127,159]](https://paperpile.com/c/zPIKrB/LX59k+l3Uth) |
| RSV-positive hospitalization rate | 10% | continuity ≥ 2, tests > 5 | 1 | [[37]](https://paperpile.com/c/zPIKrB/OpAug) |
|  | monthly average hospitalization rate | None | 1 | [[93]](https://paperpile.com/c/zPIKrB/I3rwW) |
| RSV-associated hospitalization | 2 | continuity ≥ 2 | 1 | [[189]](https://paperpile.com/c/zPIKrB/vwxwK) |
|  | 2% (week) or 8% (month) of annual hospitalization | None | 1 | [[201]](https://paperpile.com/c/zPIKrB/u8hZN) |
|  | the baseline (mean of June, July, and August during the period of study) plus 2 standard deviations (SD). | None | 1 | [[75]](https://paperpile.com/c/zPIKrB/nwDvF) |
| IRR or OR | 1  (statistically significant) | continuity ≥ 3 | 1 | [[16]](https://paperpile.com/c/zPIKrB/tl6A9) |

^a^ continuity means the minimum consecutive weeks/months meeting certain requirements were necessary to determine a season start.

**Supplementary Table 4.** Description of RSV seasons of the pattern “two-year cycle” reported by the included publications.

| **Site** | **Study** | **Years** | **Description** |
| --- | --- | --- | --- |
| Finland | Waris, 1991 [[211]](https://paperpile.com/c/zPIKrB/aDK44) | 1981-1990 | A 2-year cycle beginning in December or January: a minor peak in the spring and a major peak in the next winter. |
|  | Renko, 2019 [[129]](https://paperpile.com/c/zPIKrB/hD5ZO) | 1995-2006 | Every odd year a small spring epidemic was followed by a large epidemic peaking around December. |
| Vienna, Austria | Aberle, 2008 [[6]](https://paperpile.com/c/zPIKrB/xZQ74) | 2000 - 2007 | Early RSV seasons with peak activity in January and December were followed by late seasons with peak activity in February and March. |
| Stockholm, Sweden | Eriksson, 2002 [[63]](https://paperpile.com/c/zPIKrB/j8Al7) | 1987 - 1998 | There was a pattern of early large and late small epidemic seasons alternating biannually during the entire 12-y period. |
|  | Reyes, 1997 [[157]](https://paperpile.com/c/zPIKrB/uo77Z) | 1984-1994 | The seasonal pattern varied every other year, with late (peaks in weeks 13-18) epidemics followed by early (peaks in weeks 49-5) ones. The number of detected cases was significantly greater, around twice as many, during early than during late epidemics. |
| Kiel, Germany | Weigl, 2002 [[197]](https://paperpile.com/c/zPIKrB/7fCuW) | 1998 - 2001 | From 1997/98 onwards, a 2-year pattern with a late season starting between December and February followed by an early season starting in September to October was ob-served. Taking data from 1994 onwards into consideration the season regularly started late until 1998/99. |
|  | Weigl, 2007 [[212]](https://paperpile.com/c/zPIKrB/G5GoC) | 1996-2006 | In uneven ey, the RSV season started late at the end of December to January and was less severe than in even ey when the season started early at the end of September to October with a high incidence. |
| Stuttgart, Germany | Terletskaia-Ladwig, 2005 [[176]](https://paperpile.com/c/zPIKrB/KGT1J) | 1996-2001 | An early season with strong RSV activity (early-high phase) was followed by a weaker late season (late-low phase) in a regular biennial rhythm. |
| Germany | Reiche, 2009 [[155]](https://paperpile.com/c/zPIKrB/lp0rv) | 1998 - 2007 | A regular 2-year cyclic pattern was observed for two consecutive late and early seasons. |
|  | Obando-Pacheco, 2018 [[142]](https://paperpile.com/c/zPIKrB/h4sVb) | 2010-2017 | An early season starting in October–November and finishing in March–April and a late season starting in December and finishing in May, with both seasons having a similar duration. |
| Denmark | Jepsen, 2018 [[98]](https://paperpile.com/c/zPIKrB/EaElK) | 2010 - 2015 | Every other year the RSV season had an early start in weeks 46 to 48, and was rather mild in terms of hospitalisations. Every alternating year the RSV-season would start a few weeks later (week 50–52) and be characterized with a markedly higher RSV-hospitalization incidence. |
| Zagreb County, Croatia | [Mlinaric-Galinovic](https://pubmed.ncbi.nlm.nih.gov/?term=Mlinaric-Galinovic+G&cauthor_id=18226194), 2008 [[131]](https://paperpile.com/c/zPIKrB/otzRz) | 1994-2004 | RSV epidemics peaked in December/January of years 1994/95, 1996/97, 1998/99, 2000/01, 2002/ 03, and 2004/05 ("large seasons"), but in March/April of years 1996, 1998, 2000, 2002, and 2004 ("small seasons") |
| Switzerland | Duppenthaler,2003 [[61]](https://paperpile.com/c/zPIKrB/CpdqX) | 1997-2001 | The two minor epidemics were characterized by late onset, late peak , late end and low hospitalization rates. The major epidemics began early, peaked early, ended by week 14 and caused two to fourfold higher hospitalization rates. |
| Bismarck, USA | Irmen, 2000 [[97]](https://paperpile.com/c/zPIKrB/xcSo5) | 1987 - 1998 | The incidence of RSV tended to be higher and the month of peak RSV activity seemed to occur earlier (December through February) during the epidemic following a short interval (2- to 5-month), compared with the incidence and peak activity occurring later (March through May) following a long interval (7- to 9- month). The long interval and short interval were alternating. |
| Salt Lake County, USA | Leecaster, 2011 [[108]](https://paperpile.com/c/zPIKrB/nwSk) | 2001-2008 | The biennial variation in our seasonal epidemic data was seen in the early exponential growth rates (slope of the cumulative case curves[)](https://www.ncbi.nlm.nih.gov/pmc/articles/PMC3094225/figure/F1/) as well as total epidemic size. |
| Perth, Australia | Moore, 2009 [[133]](https://paperpile.com/c/zPIKrB/mAtYr) | 1997-2005 | The RSV identification rate showed consistent biennial peaks in even-numbered years. |

**Supplementary Table 5.** Descriptions of RSV seasons of the pattern “two peaks a year” reported by the included publications.

| **Site** | **Study** | **Years** | **Peak1** | **Peak2** |
| --- | --- | --- | --- | --- |
| Hong Kong | Mak, 2012 [[121]](https://paperpile.com/c/zPIKrB/J6R1R) | 2004 - 2011 | March - April | July - September |
|  | Tang, 2010 [[175]](https://paperpile.com/c/zPIKrB/wB8zW) | 2000 - 2007 | March - May | July - August |
|  | Chan, 2015 [[47]](https://paperpile.com/c/zPIKrB/sdUN2) | 1998 - 2012 | Week 10-15 | Week 29-38 |
| Shenzhen, China | He, 2014 [[84]](https://paperpile.com/c/zPIKrB/m7VoW) | 2007 - 2010 | March - May | November - December |
| Guangzhou, China | Liu, 2019 [[114]](https://paperpile.com/c/zPIKrB/rm9oD) | 2009 - 2016 | February - April | August - October |
| Dadaab, Kenya | Nyoka, 2017 [[140]](https://paperpile.com/c/zPIKrB/hdTg7) | 2007 - 2011 | October - January | May - July |
| Taiwan | Hsu, 2014 [[93]](https://paperpile.com/c/zPIKrB/I3rwW) | 2000 - 2010 | April | September |
|  | Chi, 2011 [[50]](https://paperpile.com/c/zPIKrB/osdms) | 2004-2007 | Spring | Autumn |
| Miami | Yusuf, 2007 [[206]](https://paperpile.com/c/zPIKrB/riwp) | 2000-2003 | Week 38-42 | January and February |
| Bardados | Li, 2019 [[3]](https://paperpile.com/c/zPIKrB/BQ74y) | 2010-2017 | February - March | July - October |
| Salvador, Brazil | Li, 2019 [[3]](https://paperpile.com/c/zPIKrB/BQ74y) | 2009-2013 | March - May | October - December |

**Supplementary Table 6.** Descriptions of RSV of “unclear pattern” reported by the included publications.

| **Site** | **Study** | **Years** | **Description** |
| --- | --- | --- | --- |
| Rio de Janeiro, Brazil | Sutmoller, 1995 [[172]](https://paperpile.com/c/zPIKrB/f9vki) | 1987-1989 | During the 3-year period, infection with RSV was clearly seasonal: increases in the late fall and winter were observed, except in 1989, when two peaks oc- curred-one in the first quarter (January-March), and one in the third quarter (July-September) |
| Nova Scotia, Canada | AI-Assam, 2009 [[8]](https://paperpile.com/c/zPIKrB/tfn1D) | 2005 - 2008 | The onset and peak periods of RSV activity varied over time, but the duration of each year’s outbreak was similar, last-ing five to six months. In 2005-2006, the RSV season started in February, peaked in April and ended in July. In 2006-2007, the onset was in December, with a peak in February and ending in late May. During 2007-2008 there were two peak periods (February/March) and (May/June) with the epidemic ending in late June. |
| Hong Kong | Liu, 2019[[114]](https://paperpile.com/c/zPIKrB/rm9oD) | 2006-2008 | There was no definite seasonality,but incidence was lowest between October and January. |
| Dhaka, Bangladesh | Stockman, 2013[[168]](https://paperpile.com/c/zPIKrB/9Yq61) | 2004 - 2008 | Annual data suggests RSV activity occurred during defined periods lasting approximately three months with no clear seasonal pattern. During the 4 study years there was one peak in January. 2006, July 2006 and October 2007. There were at least 2 months with low RSV detections between peaks. |
| Kimberley, Australia | Hogan, 2016 [[89]](https://paperpile.com/c/zPIKrB/VsIIl) | 2000-2013 | There is a less identifiable seasonal pattern in the Pilbara region in the state’s north, and no seasonal peak evident in the Kimberley region. |
| Pilbara, Australia |  |  |  |
| Nairobi, Kenya | Rose, 2020 [[160]](https://paperpile.com/c/zPIKrB/i2Xbo) | 2006-2018 | RSV did not have a clear pattern of circulation in Nairobi, and we could not define sea- son onset, offset, or peak for that region. |
| Mexico | Obando-Pacheco, 2018 [[142]](https://paperpile.com/c/zPIKrB/h4sVb) | 2012-2015 | A 2-season year is followed by a milder year, where the outbreak starts in spring and activity is maintained almost all year round with no clear peaks. |

[**Supplementary Table**](https://docs.google.com/document/d/1PIbnf9Bu1RZLooXKvEq9IuGhpqv_UyyLBUtGsyH_5B0/edit#stabl_2) **7.** Correlation analysis on the association between duration of RSV season and latitude, climatic zone, and daily average mean temperature and the absolute humidity.

| **Variable** | **Hemisphere** | **Climatic**  **zone**^b^ | **All estimates** | | **Estimates**  **from qualitative methods** | | **Estimates**  **from quantitative methods** | |
| --- | --- | --- | --- | --- | --- | --- | --- | --- |
|  |  |  | **Correlation coefficient** | **p-value** | **Correlation coefficient** | **p-value** | **Correlation coefficient** | **p-value** |
| **Latitude** ^a^ | **Northern**  **Hemisphere** | **All** | -0·38 (-0·49, -0·25) | <0·001 | -0·08 (-0·33, 0·19) | 0·60 | -0·47 (-0·59, -0·33) | <0·001 |
|  |  | **Temperate** | -0·31 (-0·44, -0·17) | <0·001 | -0·06 (-0·35, 0·24) | 0·81 | -0·39 (-0·52, -0·23) | <0·001 |
|  | **Southern**  **Hemisphere** | **All** | -0·46 (-0·67, -0·18) | 0·0020 | -0·48 (-0·27, 0·87) | 0·87 | -0·45 (-0·69, -0·13) | <0·0083 |
|  |  | **Temperate** | -0·39 (-0·68, -0·01) | 0·056 | -0·48 (-0·54, 0·93) | 0·93 | 0·037 (-0·43, 0·48) | 0·88 |
|  | **Both** | **All** | -0·36 (-0·46, -0·24) | <0·001 | -0·15 (-0·38, 0·09) | 0·22 | -0·41 (-0·53, -0·28) | <0·001 |
|  |  | **Temperate** | -0·29 (-0·41, -0·16) | <0·001 | -0·09 (-0·36, 0·19) | 0·51 | -0·35 (-0·48, -0·20) | <0·001 |
| **Daily average mean temperature** | **Northern**  **Hemisphere** | **All** | 0·33 (0·20, 0·45) | <0·001 | 0·008 (-0·26, 0·27) | 0·95 | 0·42 (0·27, 0·54) | <0·001 |
|  |  | **Temperate** | 0·29 (0·14, 0·42) | <0·001 | -0·036 (-0·33, 0·26) | 0·82 | 0·36 (0·20, 0·50) | <0·001 |
|  | **Southern**  **Hemisphere** | **All** | 0·49 (0·22, 0·69) | 0·001 | 0·53 (-0·21, 0·88) | 0·1 | 0·48 (0·17, 0·71) | 0·0046 |
|  |  | **Temperate** | 0·40 (0·005, 0·69) | 0·048 | 0·63 (-0·37, 0·95) | 0·18 | 0·071 (-0·40, 0·51) | 0·77 |
|  | **Both** | **All** | 0·33 (0·22, 0·44) | <0·001 | 0·07 (-0·177, 0·308) | 0·58 | 0·39 (0·26, 0·51) | <0·001 |
|  |  | **Temperate** | 0·28 (0·14, 0·40) | <0·001 | -0·005 (-0·28, 0·27) | 0·97 | 0·34 (0·19, 0·47) | <0·001 |
| **Daily average mean absolute humidity** | **Northern**  **Hemisphere** | **All** | 0·31 (0·18, 0·43) | <0·001 | 0·068 (-0·20, 0·33) | 0·62 | 0·39 (0·24, 0·52) | <0·001 |
|  |  | **Temperate** | 0·29 (0·14, 0·42) | <0·001 | 0·014 (-0·28, 0·31) | 0·93 | 0·35 (0·19, 0·50) | <0·001 |
|  | **Southern**  **Hemisphere** | **All** | 0·55 (0·30, 0·73) | <0·001 | 0·53 (-0·21, 0·88) | 0·15 | 0·53 (0·23, 0·74) | 0·0016 |
|  |  | **Temperate** | 0·20 (-0·21, 0·55) | 0·34 | 0·48 (-0·55, 0·93) | 0·34 | -0·55 (-0·80, -0·12) | 0·016 |
|  | **Both** | **All** | 0·34 (0·22, 0·45) | <0·001 | 0·16 (-0·092, 0·39) | 0·22 | 0·39 (0·26, 0·51) | <0·001 |
|  |  | **Temperate** | 0·272 (0·14, 0·40) | <0·001 | 0·085 (-0·20, 0·35) | 0·55 | 0·32 (0·16, 0·46) | <0·001 |
| **Climatic zone**^c^ | **Northern**  **Hemisphere** | **All** | 0·29 | <0·001 | -0·034 | 0·77 | 0·36 | <0·001 |
|  |  | **Temperate** | 0·21 | <0·001 | -0·13 | 0·33 | 0·28 | <0·001 |
|  | **Southern**  **Hemisphere** | **All** | 0·29 | 0·018 | 0·18 | 0·59 | 0·29 | 0·042 |
|  |  | **Temperate** | 0·075 | 0·66 | NA | NA | -0·043 | 0·83 |
|  | **Both** | **All** | 0·28 | <0·001 | 0·024 | 0·82 | 0·33 | <0·001 |
|  |  | **Temperate** | 0·19 | 0·0014 | -0·094 | 0·46 | 0·24 | <0·001 |
| **Analytic methods**^d^ | **Northern**  **Hemisphere** | **All** | 0·24 | <0·001 | NA | NA | 0·34 | <0·001 |
|  |  | **Temperate** | 0·31 | <0·001 | NA | NA | 0·43 | <0·001 |
|  | **Southern**  **Hemisphere** | **All** | 0·26 | 0·12 | NA | NA | 0·25 | 0·090 |
|  |  | **Temperate** | 0·32 | 0·15 | NA | NA | 0·42 | 0·035 |
|  | **Both** | **All** | 0·25 | <0·001 | NA | NA | 0·32 | <0·001 |
|  |  | **Temperate** | 0·31 | <0·001 | NA | NA | 0·42 | <0·001 |

^a^ The absolute value of the latitude coordinate was used in the correlation analysis.

^b^ In the column of Climatic zone, Temperate includes the estimates from regions where the latitude > 23·5 or <-23·5.

^c^ Climate zone includes tropics (-23·5 ° – 23·5 °), subtropics, (-35 ° – -23·5 °, 23·5 ° – 35 °), and temperate zone (> 35 °, <-35 °) Kendall's tau statistic was used to evaluate the correlations.

^d^ Analytic methods used for estimating RSV seasonality were categorized into four groups, including qualitative methods, and three groups of quantitative methods, i.e., threshold-based methods, coverage-based methods, and model-based methods (details see Supplementary material). Kendall's tau statistic was used to evaluate the correlations.

[**Supplementary Table**](https://docs.google.com/document/d/1PIbnf9Bu1RZLooXKvEq9IuGhpqv_UyyLBUtGsyH_5B0/edit#stabl_2) **8**. Associations identified in the linear regression analysis between durations of RSV seasons and study characteristics from all investigations included in the review.

| **Variable** | **Change in the duration of RSV season（months）per unit change or compared to the reference group** | **95% confidence interval** | **p-value** | **Adjusted R squared** |
| --- | --- | --- | --- | --- |
| **Climatic zone** |  |  |  |  |
| Temperate zone  (<-35° or >35°) | 0 | Referent |  |  |
| Subtropical region  (-35°- -23·5° or 23·5°- 35°) | 0·69 | 0·21, 1·18 |  |  |
| Tropical region (-23·5°– 23·5°) | 1·15 | 0·42, 1·88 |  |  |
| **Analysis Method** |  |  |  |  |
| Coverage-based method | 0 | Referent |  |  |
| Qualitative method | 1·02 | 0·52, 1·51 | <0·001 |  |
| Threshold-based method | 1·41 | 0·96, 1·86 | <0·001 |  |
| Model-based method | 1·80 | -1·10, 4·70 | 0·20 |  |
| **Meteorological Factor** |  |  |  |  |
| Daily average mean absolute humidity | 0·011 | -0·0028, 0·026 | 0·19 |  |

^a^ The absolute value of the latitude coordinate was used in the regression analysis.

**Supplementary Table 9.** Associations identified in the linear regression analysis with the alternatrive model (using latitude instead of climatic zone) between durations of RSV seasons estimated from quantitative approaches and characteristics of the studies.

| **Variable** | **Change in the duration of RSV season (months) per unit change or compared to the reference group** | **95% confidence Interval** | **p-value** | **Adjusted R^2^** |
| --- | --- | --- | --- | --- |
| **Geocode** |  |  |  | 0·31 |
| Latitude^a^ | -0·041 | -0·064, -0·017 | <0·001 |  |
| **Analysis method** |  |  |  |  |
| Coverage-based method | 0 | Referent |  |  |
| Threshold-based method | 1·35 | 0·90, 1·81 | <0·001 |  |
| Model-based method | 1·68 | -1·33, 4·69 | 0·27 |  |
| **Meteorological Factor** |  |  |  |  |
| Daily average mean absolute humidity | 0·016 | -0·0012, 0·034 | 0·067 |  |

^a^ The absolute value of the latitude coordinate was used in the regression analysis.

[**Supplementary Table**](https://docs.google.com/document/d/1PIbnf9Bu1RZLooXKvEq9IuGhpqv_UyyLBUtGsyH_5B0/edit#stabl_2) **10**. Associations identified in the linear regression analysis with alternative model (using latitude instead of climatic zone) between durations of RSV seasons and study characteristics from all investigations included in the review.

| **Variable** | **Change in the duration of RSV season（months）per unit change or compared to the reference group** | **95% confidence interval** | **p-value** | **Adjusted R squared** |
| --- | --- | --- | --- | --- |
| **Geocode** |  |  |  | 0·24 |
| Latitude | -0·030 | -0·049, -0·011 | 0·0018 |  |
| **Analysis Method** |  |  |  |  |
| Coverage-based method | 0 | Referent |  |  |
| Qualitative method | 1·02 | 0·54, 1·51 | <0·001 |  |
| Threshold-based method | 1·35 | 0·91, 1·79 | <0·001 |  |
| Model-based method | 1·61 | -1·29, 4·51 | 0·28 |  |
| **Meteorological Factor** |  |  |  |  |
| Daily average mean absolute humidity | 0·012 | -0·0019, 0·026 | 0·089 |  |

^a^ The absolute value of the latitude coordinate was used in the regression analysis.

**Supplementary Table 11.** Associations identified in the linear regression analysis with the alternatrive model (using daily average mean temperature instead of climatic zone) between durations of RSV seasons estimated from quantitative approaches and characteristics of the studies.

| **Variable** | **Change in the duration of RSV season (months) per unit change or compared to the reference group** | **95% confidence Interval** | **p-value** | **Adjusted R^2^** |
| --- | --- | --- | --- | --- |
| **Analysis method** |  |  |  | 0.28 |
| Coverage-based method | 0 | Referent |  |  |
| Threshold-based method | 1·30 | 0·83, 1·76 | <0·001 |  |
| Model-based method | 1·37 | -1·69, 4·43 | 0·38 |  |
| **Meteorological Factor** |  |  |  |  |
| Daily average mean absolute humidity | 0·022 | 0.0028, 0.041 | 0·025 |  |
| Daily average mean temperature | 0.058 | 0.0077, 0.11 | 0.024 |  |

^a^ The absolute value of the latitude coordinate was used in the regression analysis.

[**Supplementary Table**](https://docs.google.com/document/d/1PIbnf9Bu1RZLooXKvEq9IuGhpqv_UyyLBUtGsyH_5B0/edit#stabl_2) **12**. Associations identified in the linear regression analysis with alternative model (using daily average mean temperature instead of climatic zone) between durations of RSV seasons and study characteristics from all investigations included in the review.

| **Variable** | **Change in the duration of RSV season（months）per unit change or compared to the reference group** | **95% confidence interval** | **p-value** | **Adjusted R squared** |
| --- | --- | --- | --- | --- |
| **Analysis Method** |  |  |  | 0.23 |
| Coverage-based method | 0 | Referent |  |  |
| Qualitative method | 0·99 | 0·50, 1·49 | <0·001 |  |
| Threshold-based method | 1·31 | 0·87, 1·75 | <0·001 |  |
| Model-based method | 1·38 | -1·55, 4·31 | 0·36 |  |
| **Meteorological Factor** |  |  |  |  |
| Daily average mean absolute humidity | 0·018 | 0·0034, 0·032 | 0·015 |  |
| Daily average mean temperature | 0.042 | 0·0031, 0·080 | 0.034 |  |

^a^ The absolute value of the latitude coordinate was used in the regression analysis.
